# Multiorgan Outcomes Following COVID-19 Vaccine vs Infection: 30M Analysis

**DOI:** 10.64898/2026.01.30.25343064

**Authors:** Eman A. Toraih, Dara Bruce, Mohammad H. Hussein, Hani Aiash, Stephen J. Thomas

## Abstract

**Background:** Cardiovascular and cerebrovascular risks of SARS-CoV-2 infection and mRNA vaccination remain incompletely defined and lacking comparative outcomes such as sex-specific vulnerabilities.

**Methods:** Using the TriNetX Research Network (December 2020–December 2024), we identified four mutually exclusive cohorts: uninfected/unvaccinated (naïve), infected/unvaccinated, vaccinated-only, and infected/vaccinated (hybrid immunity). We compared 50 prespecified cardiovascular, cerebrovascular, and mortality outcomes across four pairwise cohort comparisons, with analyses stratified by sex and time of event windows (0–3, 3– 6, 6–9, and >9months). Different vaccine dosing strategies were analyzed.

**Results:** Among 30.3 million individuals, infection was associated with a 4.5-fold increased mortality in males and 4.0-fold in females (*p*<0.001) as well as marked increases in myocarditis, myocardial infarction, and pulmonary embolism. Inflammatory cardiac complications occurred four times more often after infection than vaccination. Vaccination alone conferred a 76% reduction in major adverse cardiovascular events (MACE) in males and 69% in females, with no detectable cardiovascular toxicity. Post-infection vaccination provided an additional 36–38% MACE reduction, though males with hybrid immunity had a late increased risk of pericarditis. Completing the two-dose vaccine series maximally reduced mortality (by 77%) and myocarditis (by 62%) versus single dosing; further doses gave minimal additional benefit but sustained the benefit of the primary vaccination series. Females had higher infection-linked myocarditis risk despite lower mortality.

**Conclusions:** SARS-CoV-2 infection confers substantially greater and sustained cardiovascular and cerebrovascular risk than mRNA vaccination, confirming a highly favorable benefit-risk profile for vaccination. These findings support extended cardiovascular surveillance after infection and targeted, risk-based vaccination strategies.

## 1. INTRODUCTION

The COVID-19 pandemic infected more than 770 million individuals, resulted in over 7 million deaths worldwide, and remains a global health threat^1^. Beyond its acute respiratory manifestations, SARS-CoV-2 infection increases the risk of cardiovascular morbidity, with survivors facing heightened risk of major adverse cardiovascular events (MACE), including myocardial injury, arrhythmias, thrombosis, and stroke—sequelae recognized as major contributors to both short- and long-term mortality ^2-5^. These events reflect convergent pathophysiologic mechanisms: direct viral invasion of cardiomyocytes via ACE2 receptors ^6^, systemic cytokine-driven inflammatory vascular injury ^7^, and the induction of a persistent hypercoagulable state ^8^. The introduction of mRNA vaccines in December 2020 altered the pandemic’s trajectory, demonstrating >90% efficacy against severe COVID-19 ^9,10^ in the months immediately following vaccination. However, rare cardiovascular adverse events following vaccination—particularly myocarditis in young males—have complicated risk-benefit assessments and contributed to vaccine hesitancy and rethinking vaccination strategies ^11-13^.

This has ignited critical debates, necessitating a precise and data-driven reassessment of the absolute and relative cardiovascular risks associated with the primary modes of immunity: natural infection, vaccination alone, and the increasingly common state of hybrid immunity (infection followed by vaccination). Despite the urgency of this clinical question, the existing literature is constrained by several key limitations. Prior studies have frequently focused on infection or vaccination in isolation, employed short follow-up periods, utilized heterogeneous outcome definitions, and rarely included robust, large-scale cohorts with hybrid immunity ^13-16^. Crucially, the temporal evolution of risk and the influence of sex-specific factors or specific vaccine dosing strategies on long-term cardiovascular outcomes remain inadequately characterized. These limitations have constrained the ability to provide definitive, globally applicable guidance for vaccination policy and clinical care.

To provide the necessary high-resolution evidence, we conducted the largest real-world, sex-stratified comparative analysis to date, leveraging a federated health network encompassing 30 million individuals. Our primary objective was to systematically quantify the incidence of cardiovascular, cerebrovascular, thrombotic, metabolic, and mortality outcomes across all clinically relevant exposure groups—unvaccinated infection, vaccination alone, and hybrid immunity—compared with infection- and vaccine-naïve controls. Temporal trajectories of risk were delineated over four post-exposure intervals (0–3, 3–6, 6–9, and >9 months). Secondary analyses examined the impact of vaccine dose strategies—including primary, monovalent, and bivalent series—on incremental risk reduction. Our study outcomes can inform vaccination policy, risk communication, and long-term patient care in the post-pandemic era.

## 2. METHODS

### 2.1 Study design and data source

We conducted a retrospective cohort study using the TriNetX research network, a federated health platform encompassing de-identified electronic medical records from 108 participating healthcare organizations, collectively representing over 150 million patients. The network provides longitudinal data, including demographics, diagnoses, procedures, medications, laboratory values, and clinical observations. Ethical approval was not required as only de-identified data were analyzed.

### 2.2 Study population and cohort definitions

Eligible individuals had documented biological sex, known race, and a minimum of two healthcare encounters between December 2020 and December 2024. COVID-19 infection was defined by positive PCR/antigen testing or ICD-10 codes (U07·1, U07·2). Vaccination status was ascertained from electronic health records, state immunization registries, and pharmacy databases. We excluded patients with clinical trial enrollment; chromosomal abnormalities; major congenital heart disease; baseline known cardiovascular or cerebrovascular disease; active/recent malignancy (<12 months); recent substance use disorders (<3 months); body mass index ≥40 kg/m^2^; Eastern Cooperative Oncology Group (ECOG) performance status ≥2; recent live vaccine receipt (<3 months); influenza infection or vaccination (<12 months); chronic hepatitis B or C; HIV; tuberculosis; or prior non-Pfizer COVID-19 vaccination (**Table S1**).

Patients were stratified into four mutually exclusive cohorts: (1) Naive—no documented infection or vaccination; (2) Infected/unvaccinated—documented infection without subsequent vaccination; (3) Vaccinated only—BNT162b2 (Pfizer-BioNTech) vaccination without prior or subsequent documented infection; (4) Infected/vaccinated (hybrid immunity)—documented infection followed by vaccination ≥14 days post-recovery. The index date was defined as the first positive test (Group 2), first vaccine dose (Groups 3 and 4), or a randomly assigned matched date (Group 1).

### 2.3 Outcome definitions

Fifty predefined clinical outcomes were evaluated in eight categories: cardiovascular events, cardiac arrhythmias, inflammatory heart disease, heart failure and cardiomyopathy, thrombotic events, cerebrovascular events, lipid abnormalities, cardiac biomarkers, healthcare utilization, and all-cause mortality. Only new-onset outcomes occurring after the index date were considered; those with prevalent endpoints were excluded from time-specific analyses. Detailed outcome definitions and codes are presented in **Table S2**. Risk analyses were conducted for four prespecified periods following index events: 0–3 months (acute), 3–6 months (subacute), 6–9 months (intermediate), and >9 months (persistent effects).

### 2.4 Statistical analysis

The primary analyses consisted of four pairwise comparisons: (1) infected, unvaccinated versus naive controls; (2) vaccinated-only versus controls; (3) infected, vaccinated versus infected, unvaccinated; and (4) vaccinated-only versus infected, unvaccinated. All analyses were stratified by biological sex. Propensity score matching was not undertaken due to the large cohort size (>30 million individuals). Simultaneous stratification by age and sex was precluded by small numbers of outcomes per subgroup, which prevented reliable age-stratified analyses. Secondary analyses evaluated the impact of vaccine dosing regimens, specifically comparing outcomes among recipients of a complete two-dose primary series versus a single dose, and among those receiving one versus multiple (monovalent or bivalent booster) doses.

Cohort sizes varied slightly between groups, as each pairwise analysis was conducted independently within the federated TriNetX research network using identical inclusion and exclusion criteria. These minor discrepancies arose from differences in query timing and data synchronization native to the federated platform. To ensure consistency, the minimum cohort size across all four-time intervals was reported in the workflow. For outcomes with fewer than ten events in a cohort, TriNetX privacy policy reports a value of “10” instead of the true count, which may inflate event rates and relative risks for these rare outcomes; zero events are accurately represented as “0”. Results in these instances should be interpreted with caution. Two-sided chi-square tests were used for categorical variables, and results are presented as event counts, percentages, relative risks (RR), and 95% confidence intervals (CI). Statistical significance was defined as p<0.05. Data were accessed and analyzed between July 1–30, 2025. Analyses used integrated TriNetX tools; visualizations were produced with RStudio (version 2025.05.1).

## 3. RESULTS

### 3.1 Study population

Between December 2020 and December 2024, 30,323,720 patients met inclusion criteria (**Figure 1**). Sex-specific group sizes and mean follow-up durations (22-26.5 months) for each pairwise comparison are summarized in **Tables S3-S10**.

**Figure 1.**
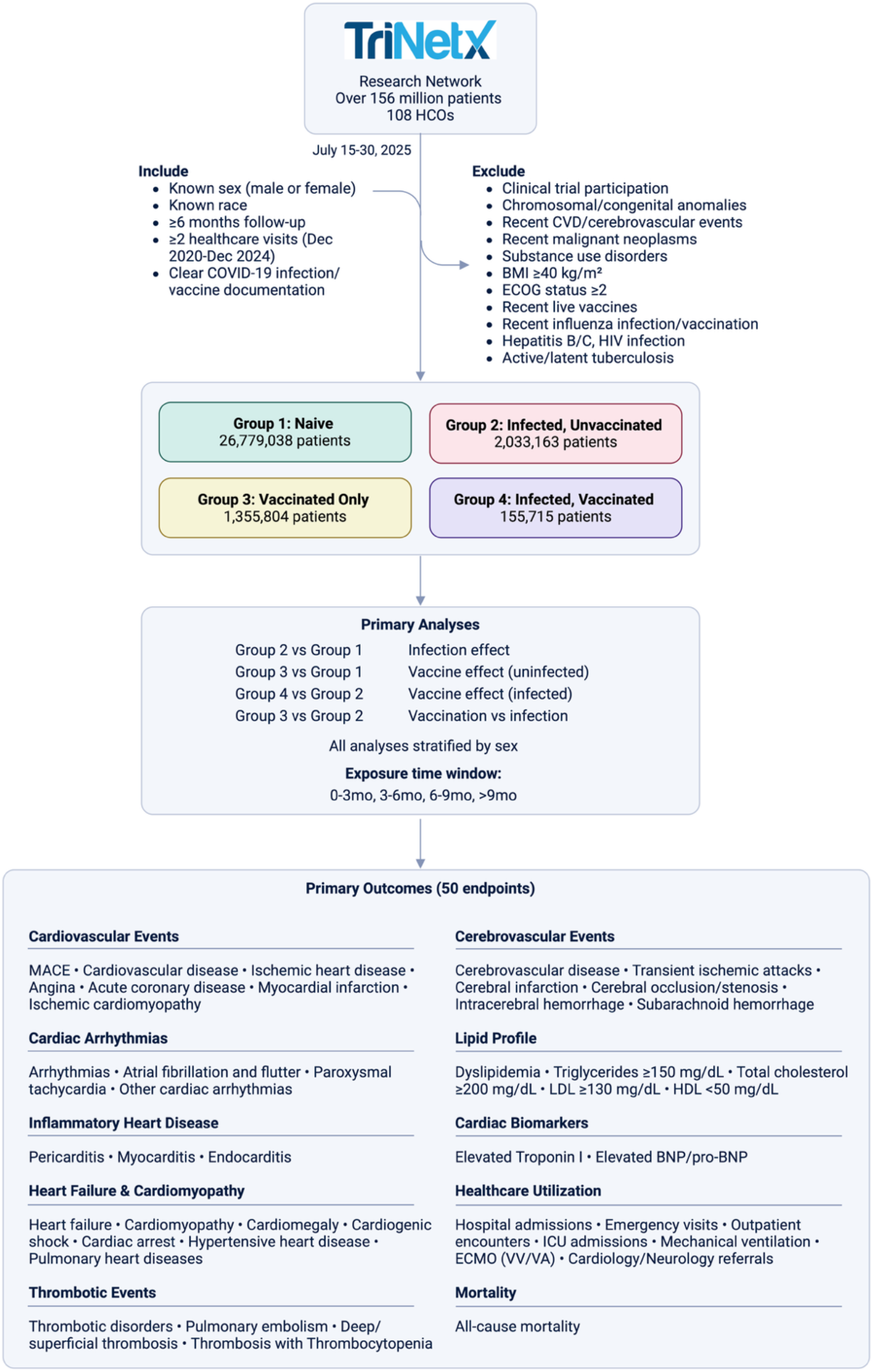
Study flowchart, cohort selection, and outcomes. Created using BioRender (biorender.com).

### 3.2 Infection effect (Group 2 vs Group 1)

COVID-19 infection in 2 million unvaccinated people led to pronounced acute and subacute increases in cardiovascular and cerebrovascular risk compared to 27 million control subjects. Myocarditis risk rose 4.44-fold in males and 5.59-fold in females; myocardial infarction nearly doubled; and acute coronary disease and stroke risks tripled. Pulmonary embolism increased fourfold in males and threefold in females. All-cause mortality was substantially higher: 2.7% in infected males vs 0.6% controls; 1.5% in infected females vs 0.4% controls. Most risks attenuated after six months, though myocarditis, mortality, and some adverse outcomes remained elevated long term (**Figure 2** and **Table S3-S4**).

**Figure 2.**
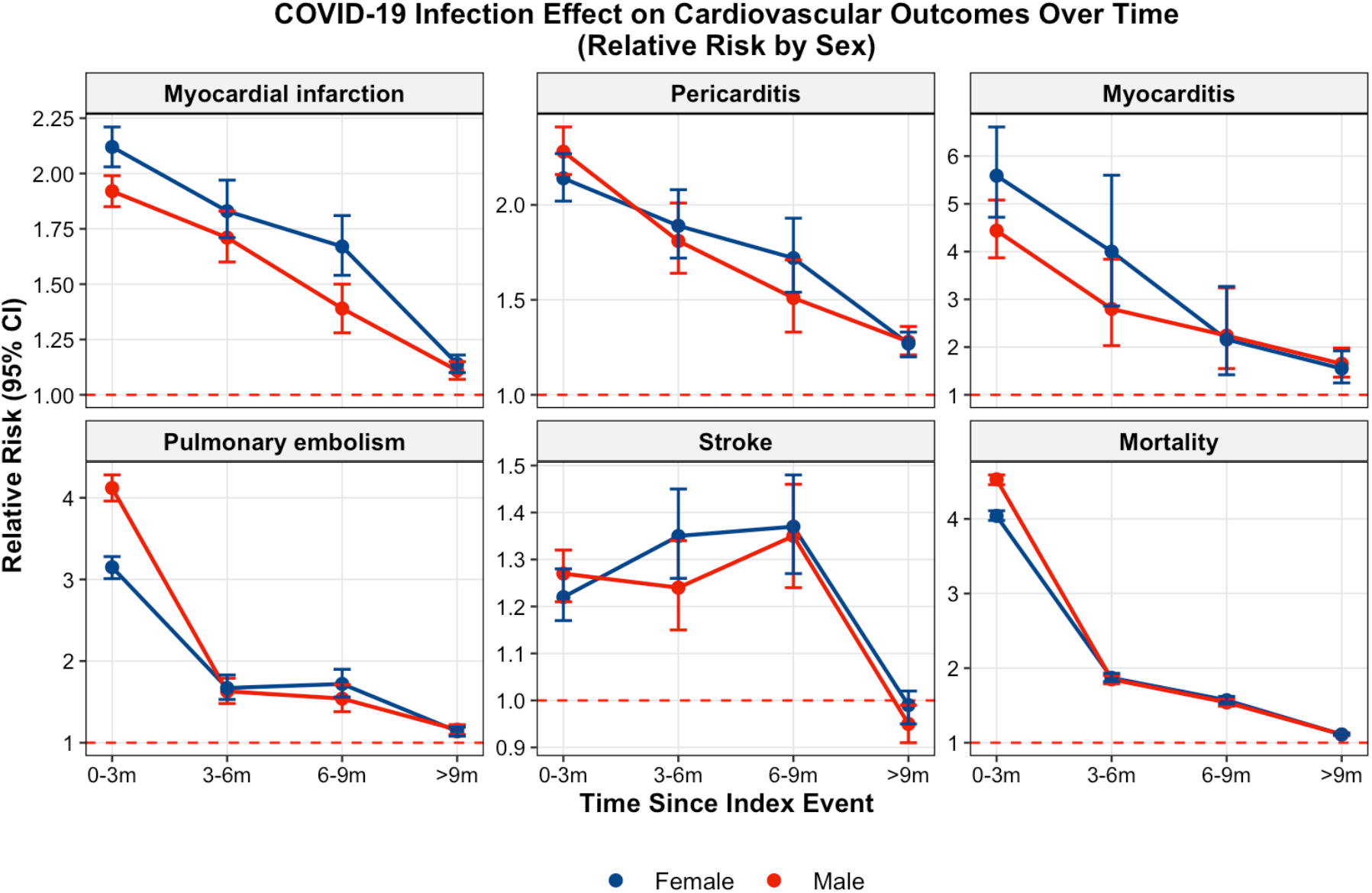
Sex-stratified risk of outcomes: Infection vs Naive Controls. Relative risk (RR) with 95% confidence intervals comparing infected, unvaccinated individuals (Group 2) with infection- and vaccine-naive controls (Group 1) across four post-infection intervals: 0–3 months (acute phase), 3–6 months (subacute phase), 6–9 months, and >9 months. The red dashed line indicates no effect (RR=1). Blue circles and lines represent females; red circles and lines represent males. Error bars denote 95% confidence intervals.

### 3.3 Vaccine effect in uninfected (Group 3 vs Group 1)

In the 1.36 million vaccinated-only group, MACE was 76% lower in males and 65% lower in females compared to controls, with sustained reductions in myocardial infarction, stroke, arrhythmias, and mortality. Benefits persisted for 3–6 months and returned to baseline after nine months, with no excess cardiovascular or inflammatory risk (**Figure 3** and **Table S5-S6**).

**Figure 3.**
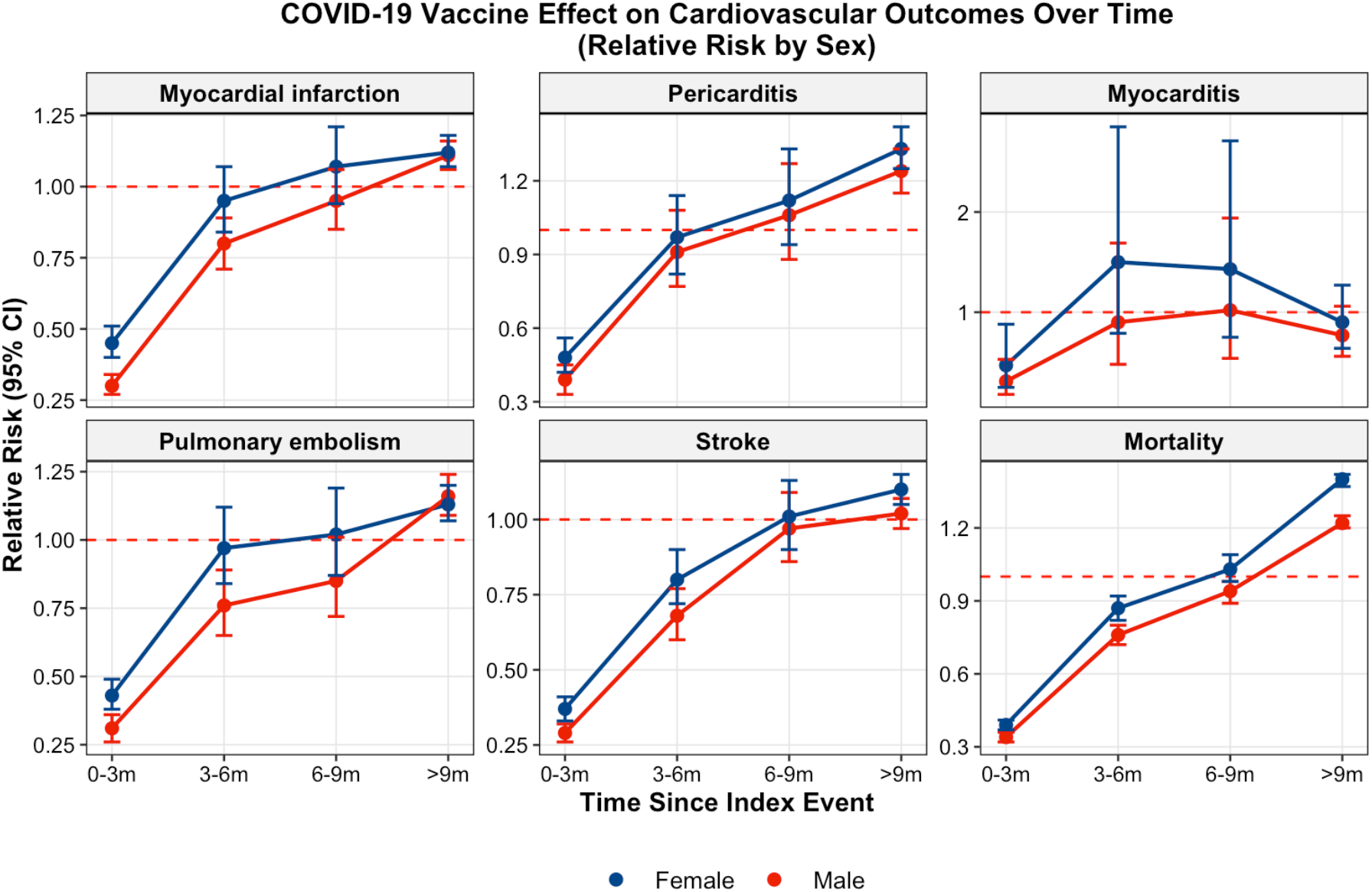
Sex-stratified risk of outcomes: Vaccination vs Naive Controls. Relative risk (RR) with 95% confidence intervals comparing vaccinated-only individuals (Group 3) with infection- and vaccine-naive controls (Group 1) across four post-vaccination intervals: 0–3, 3–6, 6–9, and >9 months. The red dashed line indicates no effect (RR=1). Blue circles and lines represent females; red circles and lines represent males. Error bars denote 95% confidence intervals.

### 3.4 Vaccine effect in infected (Group 4 vs Group 2)

Among 155,715 with hybrid immunity, post-infection vaccination reduced MACE by 36% in males and 38% in females versus infected-only patients. Most acute phase benefits waned by 6–9 months; however, in females, the reduction in cerebrovascular risk persisted beyond that of cardiac outcomes. Event counts for some endpoints were low, warranting cautious interpretation (**Figure 4** and **Table S7-S8)**.

**Figure 4.**
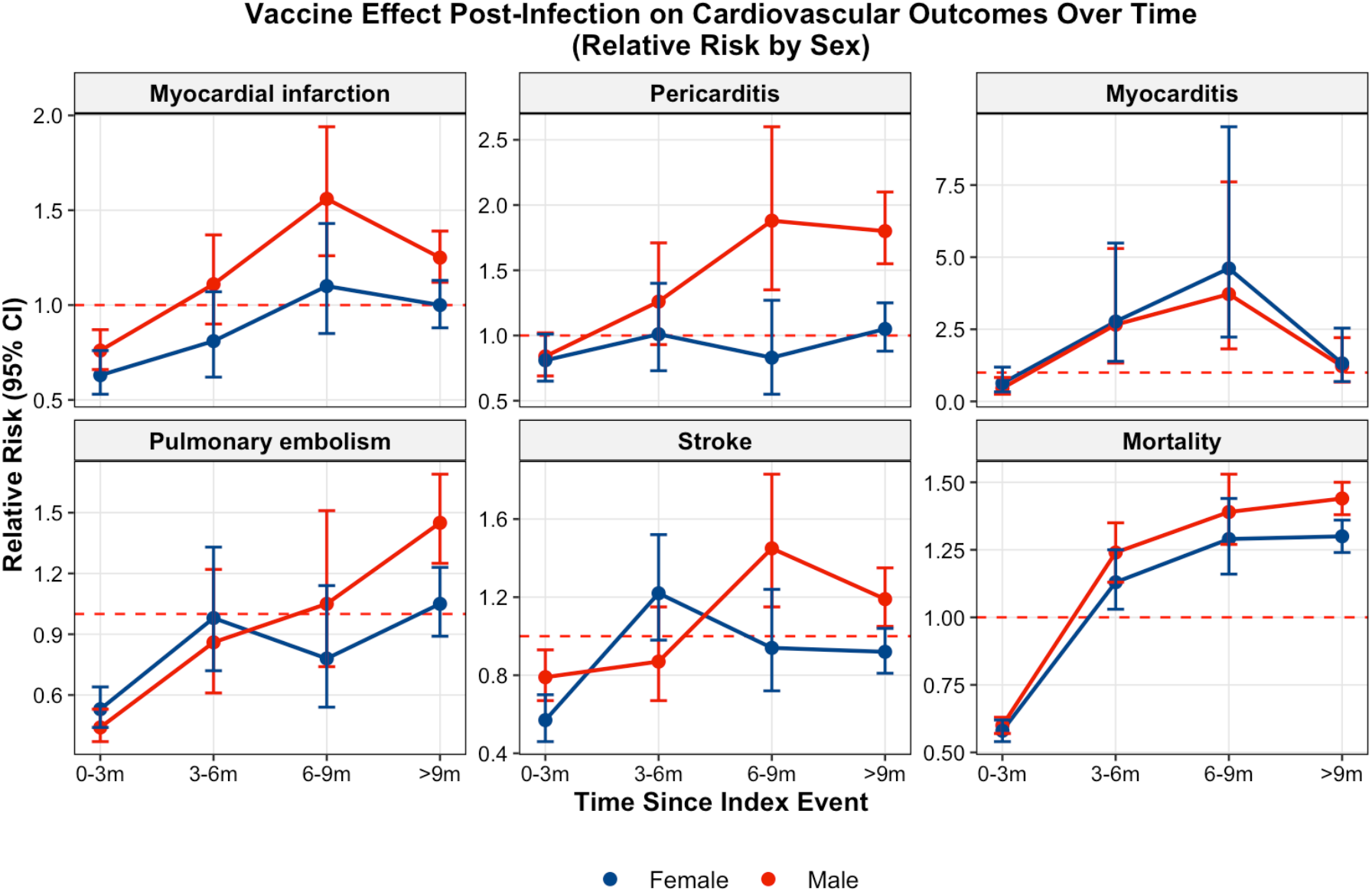
Sex-stratified risk of outcomes: Hybrid Immunity vs Infection Alone. Relative risk (RR) with 95% confidence intervals comparing individuals with hybrid immunity (infected then vaccinated, Group 4) with infected-only individuals (Group 2) across four post-vaccination intervals: 0–3, 3–6, 6–9, and >9 months. The red dashed line indicates no effect (RR=1). Blue circles and lines represent females; red circles and lines represent males. Error bars denote 95% confidence intervals.

### 3.5 Vaccination versus infection (Group 3 vs Group 2)

Directly comparing 1.4 million vaccinated-only and 2 million infected-only subjects, vaccination conferred dramatically superior safety: myocarditis risk reduced by over 90%, pulmonary embolism by up to 90%, and mortality by around 90%. Benefits persisted for nine months in females but waned slightly in males (**Figure 5** and **Table S9-S10**).

**Figure 5.**
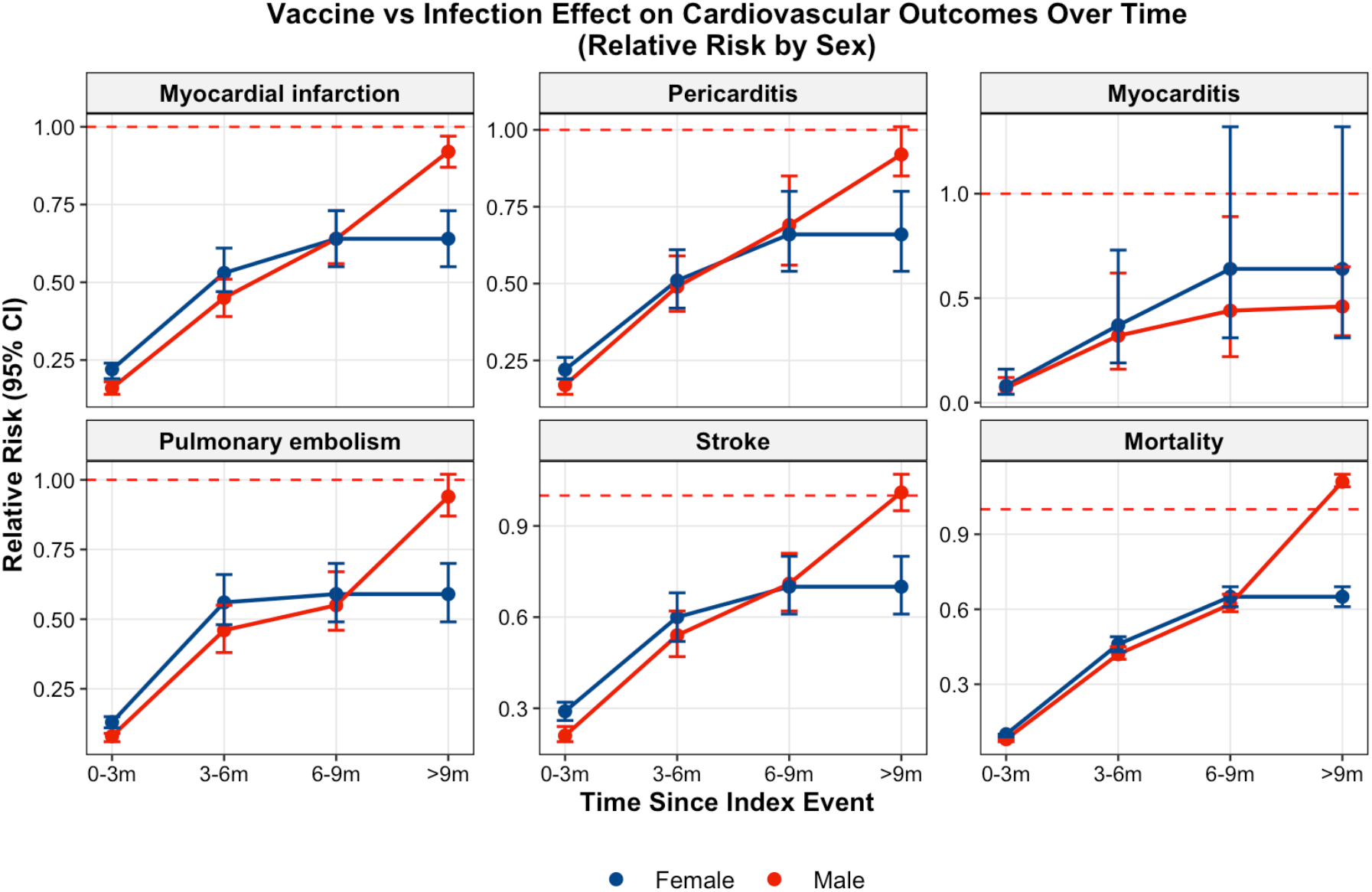
Sex-stratified risk of outcomes: Vaccination vs Infection. Relative risk (RR) with 95% confidence intervals comparing vaccinated-only individuals (Group 3) with infected-only individuals (Group 2) across four post-exposure intervals: 0–3, 3–6, 6–9, and >9 months. The red dashed line indicates no effect (RR=1). Blue circles and lines represent females; red circles and lines represent males. Error bars denote 95% confidence intervals.

### 3.6 Metabolic, biomarker, and healthcare utilization outcomes

COVID-19 infection was associated with marked metabolic derangement: hypertriglyceridemia risk increased by 33% in males and 54% in females, alongside a 23% increase in low HDL in females. Cardiac stress biomarkers such as troponin I and BNP were elevated 2.6–3.1 times over controls. Infected and unvaccinated individuals had substantially higher intensive care utilization, with mechanical ventilation required 3.5–3.9 times more often and ECMO-VV usage 13.6 times higher than in controls. In contrast, vaccination led to 60–69% reductions in dyslipidemia and 67–79% lower cardiac biomarker levels and was associated with more than 80% fewer ICU admissions (**Table S3-S10**).

### 3.7 Sex-stratified cardiovascular and cardiovascular risk patterns

Following infection, females had higher risks for infarction (RR 2.12 vs 1.92 in males), myocarditis (5.59 vs 4.44), arrhythmias (1.63 vs 1.45), and lipid derangement (1.54 vs 1.33); while males suffered more thrombotic events (4.12 vs 3.15) and mortality (4.53 vs 4.04). Vaccination provided stronger short-term benefit for males, but females maintained more durable cerebrovascular and mortality reductions. In hybrid immunity, males exhibited late pericarditis (RR 1.88 vs 0.83) while females retained cerebrovascular protection >9 months (stroke RR 0.59 vs 0.68) (**Table S3-S10**).

### 3.8 Vaccine dosing effects

Two-dose primary BNT162b2 series reduced mortality by 77%, myocarditis by 62%, pericarditis by 58%, myocardial infarction by 53%, and stroke by 48% versus a single dose. Additional monovalent doses diminished mortality by 27%, hospitalization by 42%; bivalent doses added a further 23% reduction in severe disease, yet incremental vascular risk reductions plateaued after the primary series (**Table S11**).

## 4. DISCUSSION

In this large, multinational federated cohort of over 30 million individuals, SARS-CoV-2 infection was associated with striking and sustained increases in cardiovascular and cerebrovascular risk—spanning myocarditis, infarction, thromboembolism, arrhythmias, and stroke—while mRNA vaccination conferred substantial protection without evidence of cardiovascular toxicity. The data identify infection as the primary vascular hazard, while vaccination offers significant benefit. Hybrid immunity provided further short-term protection, with late sex-specific patterns.

Natural SARS-CoV-2 infection was associated with three-to five-fold increases in cardiovascular and cerebrovascular events—including myocarditis, myocardial infarction, pulmonary embolism, arrhythmia, and stroke—during the initial three months, with risks remaining elevated through nine months and all-cause mortality in males exceeding 4.5-fold. These findings are supported by mechanistic evidence from autopsy and registry studies linking acute infection to persistent endothelial injury, chronic immune activation, and thrombo-inflammatory dysregulation ^17,18^, reframing COVID-19 as a systemic vascular disorder that extends beyond the acute respiratory phase ^19^. Notably, the cardiovascular risk attributed to infection was greater than the risk conferred by major chronic conditions such as diabetes (RR=1.8) or hypertension (RR=1.6) ^20^. Our cohort’s excess mortality and adverse event rates are at least as high as those reported in large U.S. studies (mortality RR: 3.9–4.5) ^21,22^ and surpass recent European estimates (approximately 2.8-fold) ^23^, likely reflecting differences in variant waves (Delta and Omicron versus ancestral strains), comorbidity burden (42% obesity prevalence in U.S. versus 28% in Europe), longer follow-up duration than prior reports, and evolving clinical management strategies. Taken together, these findings reinforce COVID-19 infection as a dominant, global vascular threat necessitating sustained post-infection cardiovascular surveillance to address ongoing inflammation, endothelial dysfunction, and immune activation ^24,25^.

In contrast, BNT162b2 vaccination in previously uninfected individuals led to 65–76% reductions in MACE and 61–66% lower mortality, with parallel reductions in myocardial infarction, inflammatory heart disease, and ischemic stroke. The greatest benefits were observed within three months and remained significant through six months. Rare vaccination-associated myocarditis was observed far less frequently than post-infection cases (a 4.2-fold lower risk in males, 2.3-fold lower in females), fundamentally recalibrating the benefit-risk ratio and reaffirming vaccination as a key preventive intervention ^26^.

Mechanistically, infection triggers direct viral invasion of cardiomyocytes via ACE2 receptors ^6^, provoking cytokine-mediated inflammation with interleukin-6 levels 10–100-fold higher than those after vaccination ^7^ and promoting cardiac-reactive autoantibodies ^27^. Vaccination, by contrast, elicits controlled immunological responses with limited systemic inflammation, no evidence of sustained endothelial injury, and strong formation of immune memory even after breakthrough infection—mitigating the cascade that drives cardiovascular complications ^28^.

Although spike protein has been detected in rare myocarditis cases following vaccination, the occurrence of myocarditis across different vaccine platforms—including non-mRNA vaccines— suggests that spike protein presence may serve as a biomarker of inflammatory processes rather than representing a direct causal mechanism ^29^. This cardiovascular protection persists even during breakthrough infections, because vaccine-primed memory responses prevent the cytokine storm and hyperinflammation that drive infection-associated cardiovascular events.

Dose–response analysis demonstrated that completing the initial two-dose series achieved near-maximal cardiovascular protection (mortality reduction by 77%, myocarditis by 62%, myocardial infarction by 53% vs. single dose), whereas subsequent additional doses resulted in minimal further gain, but sustained risk reductions associated with the primary vaccination series. This plateau effect diverges from the antibody-centric response but parallels influenza vaccination, suggesting cardiovascular benefit is primarily linked to moderating inflammation rather than preventing all infection ^30-32^. Meta-analytic evidence across multiple viruses, including influenza and SARS-CoV-2, further supports the concept that vaccination-driven reduction in vascular risk depends on both individual susceptibility and immune priming dynamics, rather than simple dose escalation ^33^. Thus, strategies for ongoing COVID-19 vaccination may increasingly resemble annual influenza immunization ^34^, with additional doses recommended for older adults, those with comorbidities, and other high-risk groups.

Hybrid immunity—vaccination after infection—yielded further MACE and stroke reductions, confirming additive benefit via immune and endothelial stabilization ^35,36^. However, the observed delayed pericarditis risk in males beyond six months may reflect residual confounding, as individuals receiving vaccination after infection were often those with more severe initial illness or underlying comorbidities, or may be attributable to unique immune reconstitution phenomena. These findings highlight the need for extended clinical surveillance in this subgroup.

Sex-specific findings are especially relevant: females displayed higher relative rates of MACE, arrhythmia, dyslipidemia, and stroke post-infection, while males exhibited higher mortality and thromboembolic risk. Vaccination produced greater initial benefit for males and more durable effects for females, consistent with distinct hormonal and immunological factors—such as estrogen-mediated suppression of systemic inflammation ^37,38^ and testosterone-driven pro-inflammatory bias ^39^. The delayed pericarditis seen in males with hybrid immunity may reflect cumulative inflammatory burden and tissue-resident memory T-cell responses ^40,41^.

Our data reinforce COVID-19 vaccination as a primary tool for cardiovascular risk reduction. For patients with established cardiovascular disease or risk factors, additional vaccine doses should be prioritized informed by the known kinetics of waning immunity, variant evolution and the need for vaccine reformulation, and trends in SARS-CoV-2 transmission and disease—while surveillance for late cardiac complications, particularly pericarditis in males with prior infection, should be extended. Personalized, sex-aware risk communication and follow-up protocols are warranted.

Strengths of our study include scale, temporal/sex stratification, and comprehensive outcomes in real-world settings. Limitations include absence of propensity score matching, rare-event imprecision via privacy rules, restriction to BNT162b2, and inability to stratify by age within sex. Observational design precludes definitive causality, but dose-response patterns support inference.

Future work should clarify sex-dependent immune and endothelial mechanisms, variant- and vaccine-specific risks, and the optimal timing between infection and vaccination to minimize recurrence. The mechanisms underlying myocarditis across vaccine types, and the delayed pericarditis seen in males with hybrid immunity, require targeted investigation to clarify immunological causes and improve risk prediction. Longitudinal studies, including imaging and biomarkers, are needed to refine prognosis and determine if subclinical vascular injury leads to long-term atherosclerosis or waning vaccine protection.

## Conclusions

SARS-CoV-2 infection is associated with profound cardiovascular risk, whereas mRNA vaccination offers strong and durable risk reduction. These results support COVID-19 vaccination as part of cardiovascular preventive care and inform vaccination policy and recommendations.

## Author contributions

Conceptualization: E.A.T, H.A, S.T; Methodology: E.A.T, D.B; Software: E.A.T, D.B; Investigation: E.A.T, D.B, M.H.H; Formal Analysis: E.A.T, D.B; Validation: all authors; Writing – Original Draft: E.A.T, M.H.H; Writing – Review & Editing: All authors; Supervision: H.A, S.T; Project Administration: H.A, S.T.

## Data availability

Data access is available through the TriNetX research network under appropriate data use agreements and institutional approvals.

## Acknowledgments

We acknowledge the healthcare organizations contributing data to the TriNetX research network and the patients whose de-identified data made this research possible.

## Artificial Intelligence Disclosure

During manuscript preparation, large language models were utilized as editorial assistance tools to reduce word count in compliance with journal guidelines. The following AI technologies were employed: Claude Sonnet 4.5 (Anthropic, Inc.), ChatGPT-5 (OpenAI), and Perplexity Pro (auto-selected optimal model). These tools were used exclusively for text condensation, rephrasing, and searching recent relevant articles. All quantitative data, including statistical values were manually cross-referenced with original study databases to prevent hallucinations or errors after summarization of the original draft. The authors critically reviewed, edited, and validated all AI-assisted content. No AI tools were used for data analysis, interpretation, study design, or generation of original scientific content. The authors assume complete responsibility for the accuracy, integrity, and originality of all submitted material.

