## Supplementary Tables for "Multiorgan Outcomes Following COVID-19 Vaccine vs Infection: 30M Analysis": SupplementaryTables.pdf


| Tables | Title | Page |
|---|---|---|
| Table S1 | Diagnostic, laboratory, and procedural codes used to define study cohorts. | 2-18 |
| Table S2 | Diagnostic and procedural code definitions for all 50 outcomes. | 19-22 |
| Table S3 | Outcomes in male infected, unvaccinated individuals compared to naive controls (Group 2 vs Group 1) across four post-exposure time windows. | 23-26 |
| Table S4 | Outcomes in female infected, unvaccinated individuals compared to naive controls (Group 2 vs Group 1) across four post-exposure time windows. | 27-30 |
| Table S5 | Outcomes in male in vaccinated individuals without prior infection compared to naive controls (Group 3 vs Group 1) across four post-exposure time windows. | 31-34 |
| Table S6 | Outcomes in female vaccinated individuals without prior infection compared to naive controls (Group 3 vs Group 1) across four post-exposure time windows. | 35-38 |
| Table S7 | Outcomes in male in infected, vaccinated individuals compared to infected, unvaccinated individuals (Group 4 vs Group 2) across four post-exposure time windows. | 39-42 |
| Table S8 | Outcomes in female in infected, vaccinated individuals compared to infected, unvaccinated individuals (Group 4 vs Group 2) across four post-exposure time windows. | 43-46 |
| Table S9 | Outcomes in male vaccinated individuals without infection compared to infected, unvaccinated individuals (Group 3 vs Group 2) across four post-exposure time windows. | 47-50 |
| Table S10 | Outcomes in female vaccinated individuals without infection compared to infected, unvaccinated individuals (Group 3 vs Group 2) across four post-exposure time windows. | 51-54 |
| Table S11 | Dose-dependent protection against cardiovascular events during the acute phase following Pfizer-BioNTech vaccination. | 55-58 |

**Table S1.** Diagnostic, laboratory, and procedural codes used to define study cohorts.

| Study Group | Criterion Type | System | Code | Description |
|---|---|---|---|---|
| **All study cohorts** | **Inclusion** | | | |
| | Demographics | HL7 V3.0 | F/M | Female gender or Male gender |
| | | HL7 V3.0 | 1002-5 | American Indian or Alaska Native |
| | | HL7 V3.0 | 2028-9 | Asian |
| | | HL7 V3.0 | 2054-5 | Black or African American |
| | | HL7 V3.0 | 2076-8 | Native Hawaiian or Other Pacific Islander |
| | | HL7 V3.0 | 2106-3 | White |
| | Healthcare utilization | Visit count | ≥2 visits | At least 2 healthcare visits, Dec 1, 2020- Dec 31, 2024 |
| | **Exclusion** | | | |
| | Clinical trials | LNC | 82786-5 | Identifier Clinical trial protocol |
| | | ICD-10-CM | Z00.6 | Encounter for examination for normal comparison and control in clinical research program |
| | | LNC | 35522-2 | Clinical trial protocol Quality control and quality assurance section |
| | | HCPCS | S9996 | Meals for clinical trial participant and one caregiver/companion |
| | | HCPCS | S9988 | Services provided as part of a phase i clinical trial |
| | | HCPCS | S9990 | Services provided as part of a phase ii clinical trial |
| | | HCPCS | S9991 | Services provided as part of a phase iii clinical trial |
| | | HCPCS | G9537 | Imaging needed as part of a clinical trial; or other clinician ordered the study |
| | Recent cardio/cerebral conditions last 12 months | ICD-10-CM | I20 | Angina pectoris |
| | | ICD-10-CM | I21 | Acute myocardial infarction |
| | | ICD-10-CM | I22 | Subsequent ST elevation (STEMI) and non-ST elevation (NSTEMI) myocardial infarction |
| | | ICD-10-CM | I24.9 | Acute ischemic heart disease, unspecified |
| | | ICD-10-CM | I25 | Chronic ischemic heart disease |
| | | ICD-10-CM | I26-I28 | Pulmonary heart disease and diseases of pulmonary circulation |
| | | ICD-10-CM | I26-I28 | Pulmonary embolism |
| | | ICD-10-CM | I30 | Acute pericarditis |
| | | ICD-10-CM | I31 | Other diseases of pericardium |
| | | ICD-10-CM | I32 | Pericarditis in diseases classified elsewhere |
| | | ICD-10-CM | I40 | Acute myocarditis |
| | | ICD-10-CM | I41 | Myocarditis in diseases classified elsewhere |
| | | ICD-10-CM | I42 | Cardiomyopathy |
| | | ICD-10-CM | I47 | Paroxysmal tachycardia |
| | | ICD-10-CM | I48 | Atrial fibrillation and flutter |
| | | ICD-10-CM | I49 | Other cardiac arrhythmias |
| | | ICD-10-CM | I50 | Heart failure |
| | | ICD-10-CM | I51.4 | Myocarditis, unspecified |
| | | ICD-10-CM | I51.7 | Cardiomegaly |
| | | ICD-10-CM | I60-I69 | Cerebrovascular diseases |
| | | ICD-10-CM | I61 | Nontraumatic intracerebral hemorrhage |
| | | ICD-10-CM | I63 | Cerebral infarction |
| | | ICD-10-CM | R00.0 | Tachycardia, unspecified |
| | | ICD-10-CM | R00.1 | Bradycardia, unspecified |
| | Congenital Heart Disease | ICD-10-CM | Q20 | Congenital malformations of cardiac chambers and connections |

| | | | | |
|---|---|---|---|---|
| | | ICD-10-CM | Q21 | Congenital malformations of cardiac septa |
| | | ICD-10-CM | Q22 | Congenital malformations of pulmonary and tricuspid valves |
| | | ICD-10-CM | Q23 | Congenital malformations of aortic and mitral valves |
| | | ICD-10-CM | Q24 | Other congenital malformations of heart |
| | | ICD-10-CM | Q25 | Congenital malformations of great arteries |
| | | ICD-10-CM | Q26 | Congenital malformations of great veins |
| | Neoplasm last 12 months | ICD-10-CM | C00-C14 | Malignant neoplasms of lip, oral cavity and pharynx |
| | | ICD-10-CM | C15-C26 | Malignant neoplasms of digestive organs |
| | | ICD-10-CM | C30-C39 | Malignant neoplasms of respiratory and intrathoracic organs |
| | | ICD-10-CM | C40-C41 | Malignant neoplasms of bone and articular cartilage |
| | | ICD-10-CM | C43-C44 | Melanoma and other malignant neoplasms of skin |
| | | ICD-10-CM | C45-C49 | Malignant neoplasms of mesothelial and soft tissue |
| | | ICD-10-CM | C50-C50 | Malignant neoplasms of breast (C50) |
| | | ICD-10-CM | C51-C58 | Malignant neoplasms of female genital organs |
| | | ICD-10-CM | C60-C63 | Malignant neoplasms of male genital organs |
| | | ICD-10-CM | C64-C68 | Malignant neoplasms of urinary tract |
| | | ICD-10-CM | C69-C72 | Malignant neoplasms of eye, brain and other parts of central nervous system |
| | | ICD-10-CM | C73-C75 | Malignant neoplasms of thyroid and other endocrine glands |
| | | ICD-10-CM | C76-C80 | Malignant neoplasms of ill-defined, other secondary and unspecified sites |
| | | ICD-10-CM | C7A-C7A | Malignant neuroendocrine tumors (C7A) |
| | | ICD-10-CM | C7B-C7B | Secondary neuroendocrine tumors (C7B) |
| | | ICD-10-CM | C81-C96 | Malignant neoplasms of lymphoid, hematopoietic and related tissue |
| | Recent substance use disorders last 3 months | ICD-10-CM | F10 | Alcohol related disorders |
| | | ICD-10-CM | F11 | Opioid related disorders |
| | | ICD-10-CM | F12 | Cannabis related disorders |
| | | ICD-10-CM | F13 | Sedative, hypnotic, or anxiolytic related disorders |
| | | ICD-10-CM | F14 | Cocaine related disorders |
| | | ICD-10-CM | F15 | Other stimulant related disorders |
| | | ICD-10-CM | F16 | Hallucinogen related disorders |
| | | ICD-10-CM | F19 | Other psychoactive substance related disorders |
| | BMI outside desired range | TNX | 9083 | BMI (at least 40.00 kg/m2 (most recent occurrence)) |
| | | LNC | 3915 | Body Mass Index (at least 40.00 kg/m2 (most recent occurrence)) |
| | Pregnancy (past 9 months) | ICD-10-CM | Z33.1 | Pregnant state, incidental |
| | | ICD-10-CM | O00-O9A | Pregnancy, childbirth and the puerperium (all codes) |
| | ECOG performance status score outside desired range | TNX | 2002 | ECOG Performance Status (at least 2.00 {score} (most recent occurrence)) |
| | | LNC | 89247-1 | ECOG Performance Status score (at least 2.00 {score} (most recent occurrence)) |
| | | CPT | 90476 | Adenovirus vaccine, type 4, live, for oral use |

| | | | | |
|---|---|---|---|---|
| | Live vaccine within last 3 month | CPT | 90477 | Adenovirus vaccine, type 7, live, for oral use |
| | | RxNorm | 221050 | BCG, live, Tice strain |
| | | CPT | 90585 | Bacillus Calmette-Guerin vaccine (BCG) for tuberculosis, live, for percutaneous use |
| | | CPT | 90586 | Bacillus Calmette-Guerin vaccine (BCG) for bladder cancer, live, for intravesical use |
| | | CPT | 90625 | Cholera vaccine, live, adult dosage, 1 dose schedule, for oral use |
| | | RxNorm | 798286 | human-bovine reassortant rotavirus strain G1 vaccine |
| | | RxNorm | 798288 | human-bovine reassortant rotavirus strain G2 vaccine |
| | | RxNorm | 798290 | human-bovine reassortant rotavirus strain G3 vaccine |
| | | RxNorm | 798292 | human-bovine reassortant rotavirus strain G4 vaccine |
| | | RxNorm | 798294 | human-bovine reassortant rotavirus strain P1A[8] vaccine |
| | | CPT | 90660 | Influenza virus vaccine, trivalent, live (LAIV3), for intranasal use |
| | | CPT | 90664 | Influenza virus vaccine, live (LAIV), pandemic formulation, for intranasal use |
| | | CPT | 90672 | Influenza virus vaccine, quadrivalent, live (LAIV4), for intranasal use |
| | | CPT | 90673 | Influenza virus vaccine, trivalent (RIV3), derived from recombinant DNA, hemagglutinin (HA) protein only, preservative and antibiotic free, for intramuscular use |
| | | RxNorm | 1005911 | influenza virus vaccine, live attenuated, A-Perth-16-2009 (H3N2) strain |
| | | RxNorm | 1005909 | Influenza Virus Vaccine, Live Attenuated, A-California-7-2009 (H1N1) strain (deprecated 2016) |
| | | CPT | 90705 | Measles virus vaccine, live, for subcutaneous use (deprecated 2018) |
| | | CPT | 90707 | Measles, mumps and rubella virus vaccine (MMR), live, for subcutaneous use |
| | | CPT | 90710 | Measles, mumps, rubella, and varicella vaccine (MMRV), live, for subcutaneous use |
| | | CPT | 90708 | Measles and rubella virus vaccine, live, for subcutaneous use (deprecated 2018) |
| | | RxNorm | 804179 | measles virus vaccine live, Enders' attenuated Edmonston strain |
| | | RxNorm | 2603499 | measles virus vaccine live, attenuated Schwarz strain |
| | | SNOMED | 5.72481E+14 | Administration of live attenuated measles vaccine |
| | | CPT | 90704 | Mumps virus vaccine, live, for subcutaneous use (deprecated 2018) |
| | | RxNorm | 798372 | mumps virus vaccine live, Jeryl Lynn strain |
| | | CPT | 90712 | Poliovirus vaccine, (any type[s]) (OPV), live, for oral use (deprecated 2018) |
| | | RxNorm | OMOP1004329 | Poliomyelitis Virus Type 2 (Sabin Strain (P712, Ch, 2ab)) Live Attenuated Oral Vaccine |
| | | RxNorm | OMOP1004287 | Poliomyelitis Virus Type 3 (Sabin Strain (Leon 12a1b)) Live Attenuated Oral Vaccine |

| | | | | |
|---|---|---|---|---|
| | | CPT | 90680 | Rotavirus vaccine, pentavalent (RV5), 3 dose schedule, live, for oral use |
| | | CPT | 90681 | Rotavirus vaccine, human, attenuated (RV1), 2 dose schedule, live, for oral use |
| | | RxNorm | 805573 | rotavirus vaccine, live attenuated, G1P[8] human 89-12 strain |
| | | CPT | 90706 | Rubella virus vaccine, live, for subcutaneous use (deprecated 2018) |
| | | RxNorm | 762817 | rubella virus vaccine live (Wistar RA 27-3 strain) |
| | | RxNorm | 762595 | Salmonella typhi Ty21a live antigen |
| | | CPT | 90690 | Typhoid vaccine, live, oral |
| | | RxNorm | 807219 | typhoid Vi polysaccharide vaccine, S typhi Ty2 strain |
| | | CPT | 90716 | Varicella virus vaccine (VAR), live, for subcutaneous use |
| | | RxNorm | 1292422 | varicella-zoster virus vaccine live (Oka-Merck) strain |
| | | CPT | 90717 | Yellow fever vaccine, live, for subcutaneous use |
| | | RxNorm | 804187 | yellow fever virus strain 17D-204 live antigen |
| | | CPT | 90736 | Zoster (shingles) vaccine (HZV), live, for subcutaneous injection |
| | Influenza flu last 12 months | LNC | 34487-9 | Influenza virus A RNA [Presence] in Specimen by NAA with probe detection (labResult: Positive) |
| | | LNC | 38381-0 | Influenza virus A cDNA [Presence] in Specimen by NAA with probe detection (labResult: Positive) |
| | | LNC | 38382-8 | Influenza virus B [Presence] in Specimen by Organism specific culture (labResult: Positive) |
| | | LNC | 48310-7 | Influenza virus A [Presence] in Specimen by Organism specific culture (labResult: Positive) |
| | | LNC | 49523-4 | Influenza virus A H3 RNA [Presence] in Isolate by NAA with probe detection (labResult: Positive) |
| | | LNC | 49531-7 | Influenza virus A RNA [Presence] in Isolate by NAA with probe detection (labResult: Positive) |
| | | LNC | 49535-8 | Influenza virus B RNA [Presence] in Isolate by NAA with probe detection (labResult: Positive) |
| | | LNC | 49520-0 | Influenza virus A H1 RNA [Presence] in Isolate by NAA with probe detection (labResult: Positive) |
| | | LNC | 76078-5 | Influenza virus A RNA [Presence] in Nasopharynx by NAA with probe detection (labResult: Positive) |
| | | LNC | 80588-7 | Influenza virus A M gene [Presence] in Nasopharynx by NAA with probe detection (labResult: Positive) |
| | | LNC | 80590-3 | Influenza virus A H3 HA gene [Presence] in Nasopharynx by NAA with probe detection (labResult: Positive) |
| | | LNC | 82166-0 | Influenza virus A RNA [Presence] in Nasopharynx by NAA with non-probe detection (labResult: Positive) |
| | | LNC | 49521-8 | Influenza virus A H1 RNA [Presence] in Specimen by NAA with probe detection (labResult: Positive) |

| | | | | |
|---|---|---|---|---|
| | | LNC | 49524-2 | Influenza virus A H3 RNA [Presence] in Specimen by NAA with probe detection (labResult: Positive) |
| | | LNC | 76079-3 | Influenza virus B RNA [Presence] in Bronchoalveolar lavage by NAA with probe detection (labResult: Positive) |
| | | LNC | 76080-1 | Influenza virus B RNA [Presence] in Nasopharynx by NAA with probe detection (labResult: Positive) |
| | | LNC | 77026-3 | Influenza virus A H1 RNA [Presence] in Nasopharynx by NAA with probe detection (labResult: Positive) |
| | | LNC | 77027-1 | Influenza virus A H3 RNA [Presence] in Nasopharynx by NAA with probe detection (labResult: Positive) |
| | | LNC | 82167-8 | Influenza virus A H1 RNA [Presence] in Nasopharynx by NAA with non-probe detection (labResult: Positive) |
| | | LNC | 82169-4 | Influenza virus A H3 RNA [Presence] in Nasopharynx by NAA with non-probe detection (labResult: Positive) |
| | | LNC | 82170-2 | Influenza virus B RNA [Presence] in Nasopharynx by NAA with non-probe detection (labResult: Positive) |
| | | LNC | 92142-9 | Influenza virus A RNA [Presence] in Respiratory system specimen by NAA with probe detection (labResult: Positive) |
| | | LNC | 40982-1 | Influenza virus B RNA [Presence] in Specimen by NAA with probe detection (labResult: Positive) |
| | | LNC | 62462-7 | Influenza virus A+B RNA [Presence] in Specimen by NAA with probe detection (labResult: Positive) |
| | HCV diagnosis + viral labs last 12 months | LNC | 5012-0 | Hepatitis C virus RNA [Presence] in Specimen by NAA with probe detection (labResult: Positive) |
| | | LNC | 5010-4 | Hepatitis C virus RNA [Presence] in Blood by NAA with probe detection (labResult: Positive) |
| | | LNC | 48576-3 | Hepatitis C virus RNA [Presence] in Specimen by Probe with signal amplification (labResult: Positive) |
| | | LNC | 11259-9 | Hepatitis C virus RNA [Presence] in Serum or Plasma by NAA with probe detection (labResult: Positive) |
| | | LNC | 49379-1 | Hepatitis C virus RNA [Units/volume] (viral load) in Tissue by NAA with probe detection (at least 15.00 [IU]/mL (most recent occurrence)) |
| | | LNC | 49376-7 | Hepatitis C virus RNA [Units/volume] (viral load) in Specimen by NAA with probe detection (at least 15.00 [IU]/mL (most recent occurrence)) |
| | | LNC | 49380-9 | Hepatitis C virus RNA [#/volume] (viral load) in Specimen by NAA with probe detection (at least 50.00 {copies}/mL (most recent occurrence)) |
| | | LNC | 10676-5 | Hepatitis C virus RNA [Units/volume] (viral load) in Serum or Plasma by Probe with |

| | | |
|---|---|---|
| | | amplification (at least 15.00 [IU]/mL (most recent occurrence)) |
| LNC | 49375-9 | Hepatitis C virus RNA [Log #/volume] (viral load) in Tissue by NAA with probe detection (at least 1.10 {Log_copies}/mL (most recent occurrence)) |
| LNC | 50023-1 | Hepatitis C virus RNA panel (viral load) in Serum or Plasma by NAA with probe detection (at least 15.00 units (most recent occurrence)) |
| LNC | 20571-6 | Hepatitis C virus RNA [#/volume] (viral load) in Serum or Plasma by Probe with signal amplification |
| LNC | 29609-5 | Hepatitis C virus RNA [Units/volume] (viral load) in Serum or Plasma by Probe with signal amplification (at least 15.00 [IU]/mL (most recent occurrence)) |
| LNC | 20416-4 | Hepatitis C virus RNA [#/volume] (viral load) in Serum or Plasma by NAA with probe detection (at least 50.00 {copies}/mL (most recent occurrence)) |
| LNC | 11011-4 | Hepatitis C virus RNA [Units/volume] (viral load) in Serum or Plasma by NAA with probe detection (at least 15.00 [IU]/mL (most recent occurrence)) |
| LNC | 49605-9 | Hepatitis C virus RNA [log units/volume] (viral load) in Specimen by NAA with probe detection (at least 1.10 {Log_IU}/mL (most recent occurrence)) |
| LNC | 38180-6 | Hepatitis C virus RNA [log units/volume] (viral load) in Serum or Plasma by NAA with probe detection (at least 1.10 {Log_IU}/mL (most recent occurrence)) |
| LNC | 47252-2 | Hepatitis C virus RNA [Log #/volume] (viral load) in Serum or Plasma by NAA with probe detection (at least 1.10 {log_copies}/mL (most recent occurrence)) |
| LNC | 42617-1 | Hepatitis C virus RNA [log units/volume] (viral load) in Serum or Plasma by Probe with signal amplification (at least 1.10 {Log_IU}/mL (most recent occurrence)) |
| LNC | 49372-6 | Hepatitis C virus RNA [Log #/volume] (viral load) in Specimen by NAA with probe detection (at least 1.10 {Log_copies}/mL (most recent occurrence)) |
| LNC | 49758-6 | Hepatitis C virus RNA [Units/volume] (viral load) in Serum or Plasma by Probe and target amplification method detection limit = 5 iU/mL (at least 15.00 [IU]/mL (most recent occurrence)) |
| LNC | 34704-7 | Hepatitis C virus RNA [Units/volume] (viral load) in Serum or Plasma by Probe and target amplification method detection limit = 50 iU/mL (at least 15.00 [IU]/mL (most recent occurrence)) |
| CPT | 87902 | Infectious agent genotype analysis by nucleic acid (DNA or RNA); Hepatitis C virus |
| CPT | 81596 | Infectious disease, chronic hepatitis C virus (HCV) infection, six biochemical assays (ALT, |

| | | | | |
|---|---|---|---|---|
| | | | | A2-macroglobulin, apolipoprotein A-1, total bilirubin, GGT, and haptoglobin) utilizing serum, prognostic algorithm reported as scores for fibrosis and necroinflammatory activity in liver |
| | | CPT | 86804 | Hepatitis C antibody; confirmatory test (eg, immunoblot) |
| | | CPT | 86803 | Hepatitis C antibody |
| | | CPT | 1014274 | Hepatitis C antibody |
| | | SNOMED | 395197008 | Hepatitis C viral load |
| | | SNOMED | 64411004 | Hepatitis C antibody measurement |
| | | SNOMED | 397662006 | Hepatitis C virus genotype determination |
| | | RXNORM | OMOP5179608 | HEPATITIS C IMMUNE GLOBULIN (HUMAN) |
| | | ICD-10-CM | B17.10 | Acute hepatitis C without hepatic coma |
| | | ICD-10-CM | B17.11 | Acute hepatitis C with hepatic coma |
| | | ICD-10-CM | B17.1 | Acute hepatitis C |
| | | ICD-10-CM | B18.2 | Chronic viral hepatitis C |
| | | ICD-10-CM | B19.20 | Unspecified viral hepatitis C without hepatic coma |
| | | ICD-10-CM | B19.21 | Unspecified viral hepatitis C with hepatic coma |
| | Hepatitis B last 12 months | ICD-10-CM | B16 | Acute hepatitis B |
| | | ICD-10-CM | B16.1 | Acute hepatitis B with delta-agent without hepatic coma |
| | | ICD-10-CM | B19.1 | Unspecified viral hepatitis B |
| | | ICD-10-CM | B19.10 | Unspecified viral hepatitis B without hepatic coma |
| | | ICD-10-CM | B19.11 | Unspecified viral hepatitis B with hepatic coma |
| | | ICD-10-CM | Z22.51 | Carrier of viral hepatitis B (deprecated 2018) |
| | | ICD-10-CM | B18.1 | Chronic viral hepatitis B without delta-agent |
| | | TNX | 9059 | Hepatitis B virus surface Ab [Units/volume] in Serum (at least 0.00 m[IU]/mL (most recent occurrence)) |
| | | LNC | 5195-3 | Hepatitis B virus surface Ag [Presence] in Serum (labResult: Positive) |
| | HIV diagnosis or viral labs last 12 months | ICD-10-CM | B20 | Human immunodeficiency virus [HIV] disease |
| | | ICD-10-CM | B20-B20 | Human immunodeficiency virus [HIV] disease (B20) |
| | | ICD-10-CM | Z21 | Asymptomatic human immunodeficiency virus [HIV] infection status |
| | | ICD-10-CM | O98.71 | Human immunodeficiency virus [HIV] disease complicating pregnancy |
| | | ICD-10-CM | O98.72 | Human immunodeficiency virus [HIV] disease complicating childbirth |
| | | ICD-10-CM | B97.35 | Human immunodeficiency virus, type 2 [HIV 2] as the cause of diseases classified elsewhere |
| | | BIOM | 3918\|1 | Human Immunodeficiency Virus (HIV)/Acquired Immunodeficiency Syndrome (AIDS) |
| | | BIOM | 3859\|1 | Associated with HIV/AIDS HIV positive |
| | | BIOM | 3859 | HIV Status |
| | | SNOMED | 315124004 | HIV viral load |
| | | SNOMED | 446987006 | HIV 1 genotyping |
| | | LNC | 59052-1 | HIV 1+Hepatitis C virus RNA+Hepatitis B virus DNA [Presence] in Serum or Plasma by NAA with probe detection (labResult: Positive) |
| | | LNC | 48558-1 | HIV genotype [Susceptibility] |

| | | |
|---|---|---|
| LNC | 45182-3 | HIV phenotype [Susceptibility] |
| LNC | 48346-1 | HIV 1+O+2 Ab [Units/volume] in Serum or Plasma |
| LNC | 49573-9 | HIV genotype [Susceptibility] in Isolate by Genotype method Narrative |
| LNC | 22358-6 | HIV 2 Ab [Units/volume] in Serum |
| LNC | 83101-6 | HIV 1+2 Ab and HIV1 p24 Ag panel- Serum or Plasma by Immunoassay |
| LNC | 85380-4 | HIV immunoassay testing algorithm interpretation in Serum, Plasma or Blood |
| LNC | 73658-7 | HIV 1 subtype in Specimen by NAA with probe detection |
| LNC | 29539-4 | HIV 1 RNA [Log #/volume] (viral load) in Plasma by Probe with signal amplification (at least 1.30 {log_copies}/mL (most recent occurrence)) |
| LNC | 21008-8 | HIV 1 RNA [#/volume] (viral load) in Serum or Plasma by Probe (at least 20.00 {copies}/mL (most recent occurrence)) |
| LNC | 23876-6 | HIV 1 RNA [Units/volume] (viral load) in Plasma by Probe with signal amplification (at least 20.00 [IU]/mL (most recent occurrence)) |
| LNC | 10351-5 | HIV 1 RNA [Units/volume] (viral load) in Serum or Plasma by Probe with amplification (at least 20.00 [IU]/mL (most recent occurrence)) |
| LNC | 59419-2 | HIV 1 RNA [#/volume] (viral load) in Plasma by Probe with signal amplification (at least 20.00 {copies}/mL (most recent occurrence)) |
| LNC | 29541-0 | HIV 1 RNA [Log #/volume] (viral load) in Serum or Plasma by NAA with probe detection (at least 1.30 {log_copies}/mL (most recent occurrence)) |
| LNC | 41498-7 | HIV 1 RNA [#/volume] (viral load) in Cerebral spinal fluid by NAA with probe detection (at least 20.00 {copies}/mL (most recent occurrence)) |
| LNC | 41497-9 | HIV 1 RNA [Log #/volume] (viral load) in Cerebral spinal fluid by NAA with probe detection (at least 1.30 {Log_copies}/mL (most recent occurrence)) |
| LNC | 70241-5 | HIV 1 RNA [#/volume] (viral load) in Plasma by Probe and target amplification method detection limit = 20 copies/mL (at least 20.00 {copies}/mL (most recent occurrence)) |
| LNC | 51780-5 | HIV 1 RNA [Log #/volume] (viral load) in Serum or Plasma by Probe and target amplification method detection limit = 0.5 log copies/mL (at least 1.30 {Log_copies}/mL (most recent occurrence)) |
| LNC | 25835-0 | HIV 1 RNA [Presence] in Serum or Plasma by NAA with probe detection (labResult: Positive) |
| LNC | 5018-7 | HIV 1 RNA [Presence] in Specimen by NAA with probe detection (labResult: Positive) |
| LNC | 47359-5 | HIV 1 RNA [Presence] in Serum or Plasma from Donor by Probe with amplification (labResult: Positive) |

| | | | | |
|---|---|---|---|---|
| | | LNC | 42917-5 | HIV 1 RNA [Presence] in Cerebral spinal fluid by NAA with probe detection (labResult: Positive) |
| | | LNC | 41513-3 | HIV 1 RNA [#/volume] (viral load) in Serum or Plasma by Probe with amplification detection limit = 400 copies/mL (at least 20.00 {copies}/mL (most recent occurrence)) |
| | | LNC | 41514-1 | HIV 1 RNA [Log #/volume] (viral load) in Serum or Plasma by Probe with amplification detection limit = 2.6 log copies/mL (at least 1.30 {Log_copies}/mL (most recent occurrence)) |
| | | LNC | 41515-8 | HIV 1 RNA [#/volume] (viral load) in Serum or Plasma by Probe with amplification detection limit = 75 copies/mL (at least 20.00 {copies}/mL (most recent occurrence)) |
| | | LNC | 41516-6 | HIV 1 RNA [Log #/volume] (viral load) in Serum or Plasma by Probe with amplification detection limit = 1.9 log copies/mL (at least 1.30 {Log_copies}/mL (most recent occurrence)) |
| | | LNC | 48510-2 | HIV 1 RNA [Log #/volume] (viral load) in Serum or Plasma by Probe and target amplification method detection limit = 1.7 log copies/mL (at least 1.30 {Log_copies}/mL (most recent occurrence)) |
| | | LNC | 48511-0 | HIV 1 RNA [#/volume] (viral load) in Serum or Plasma by Probe and target amplification method detection limit = 50 copies/mL (at least 20.00 {copies}/mL (most recent occurrence)) |
| | | LNC | 48552-4 | HIV 1 RNA [Log #/volume] (viral load) in Serum or Plasma by Probe and target amplification method detection limit = 2.6 log copies/mL (at least 1.30 {Log_copies}/mL (most recent occurrence)) |
| | | LNC | 21333-0 | HIV 1 RNA [#/volume] in Serum (at least 20.00 {copies}/mL (most recent occurrence)) |
| | | LNC | 62469-2 | HIV 1 RNA [Units/volume] (viral load) in Serum or Plasma by NAA with probe detection (at least 20.00 [IU]/mL (most recent occurrence)) |
| | | LNC | 49890-7 | HIV 1 RNA [Log #/volume] (viral load) in Specimen by NAA with probe detection (at least 1.30 {Log_copies}/mL (most recent occurrence)) |
| | | LNC | 25836-8 | HIV 1 RNA [#/volume] (viral load) in Specimen by NAA with probe detection (at least 20.00 {copies}/mL (most recent occurrence)) |
| | | LNC | 20447-9 | HIV 1 RNA [#/volume] (viral load) in Serum or Plasma by NAA with probe detection (at least 20.00 {copies}/mL (most recent occurrence)) |
| | TB within last 12 months | ICD-10-CM | A15-A19 | Tuberculosis |
| | Influenza vaccinate last 12 months | CPT | 90653 | Influenza vaccine, inactivated (IIV), subunit, adjuvanted, for intramuscular use |
| | | CPT | 90654 | Influenza virus vaccine, trivalent (IIV3), split virus, preservative-free, for intradermal use |
| | | CPT | 90656 | Influenza virus vaccine, trivalent (IIV3), split virus, preservative free, 0.5 mL dosage, for intramuscular use |

| | | |
|---|---|---|
| CPT | 90655 | Influenza virus vaccine, trivalent (IIV3), split virus, preservative free, 0.25 mL dosage, for intramuscular use |
| CPT | 90657 | Influenza virus vaccine, trivalent (IIV3), split virus, 0.25 mL dosage, for intramuscular use |
| CPT | 90658 | Influenza virus vaccine, trivalent (IIV3), split virus, 0.5 mL dosage, for intramuscular use |
| CPT | 90660 | Influenza virus vaccine, trivalent, live (LAIV3), for intranasal use |
| CPT | 90661 | Influenza virus vaccine, trivalent (ccIIV3), derived from cell cultures, subunit, preservative and antibiotic free, 0.5 mL dosage, for intramuscular use |
| CPT | 90662 | Influenza virus vaccine (IIV), split virus, preservative free, enhanced immunogenicity via increased antigen content, for intramuscular use |
| CPT | 90664 | Influenza virus vaccine, live (LAIV), pandemic formulation, for intranasal use |
| CPT | 90666 | Influenza virus vaccine (IIV), pandemic formulation, split virus, preservative free, for intramuscular use |
| CPT | 90667 | Influenza virus vaccine (IIV), pandemic formulation, split virus, adjuvanted, for intramuscular use |
| CPT | 90668 | Influenza virus vaccine (IIV), pandemic formulation, split virus, for intramuscular use |
| CPT | 90672 | Influenza virus vaccine, quadrivalent, live (LAIV4), for intranasal use |
| CPT | 90673 | Influenza virus vaccine, trivalent (RIV3), derived from recombinant DNA, hemagglutinin (HA) protein only, preservative and antibiotic free, for intramuscular use |
| CPT | 90674 | Influenza virus vaccine, quadrivalent (ccIIV4), derived from cell cultures, subunit, preservative and antibiotic free, 0.5 mL dosage, for intramuscular use |
| CPT | 90682 | Influenza virus vaccine, quadrivalent (RIV4), derived from recombinant DNA, hemagglutinin (HA) protein only, preservative and antibiotic free, for intramuscular use |
| CPT | 90686 | Influenza virus vaccine, quadrivalent (IIV4), split virus, preservative free, 0.5 mL dosage, for intramuscular use |
| CPT | 90687 | Influenza virus vaccine, quadrivalent (IIV4), split virus, 0.25 mL dosage, for intramuscular use |
| CPT | 90688 | Influenza virus vaccine, quadrivalent (IIV4), split virus, 0.5 mL dosage, for intramuscular use |
| CPT | 90689 | Influenza virus vaccine, quadrivalent (IIV4), inactivated, adjuvanted, preservative free, 0.25 mL dosage, for intramuscular use |
| CPT | 90694 | Influenza virus vaccine, quadrivalent (aIIV4), inactivated, adjuvanted, preservative free, 0.5 mL dosage, for intramuscular use |
| CPT | 90756 | Influenza virus vaccine, quadrivalent (ccIIV4), derived from cell cultures, subunit, antibiotic free, 0.5 mL dosage, for intramuscular use |

| | | | | |
|---|---|---|---|---|
| | | SNOMED | 86198006 | Administration of influenza vaccine |
| | | ICD-10-PCS | 3E02340 | Introduction of Influenza Vaccine into Muscle, Percutaneous Approach |
| | Ever received non-Pfizer COVID vaccinate | | | |
| | *AstraZeneca (viral vector)* | CPT | 91302 | Severe acute respiratory syndrome coronavirus 2 (SARS-CoV-2) (coronavirus disease [COVID-19]) vaccine, DNA, spike protein, chimpanzee adenovirus Oxford 1 (ChAdOx1) vector, preservative free, 5x1010 viral particles/0.5 mL dosage, for intramuscular use (deprecated 2024) |
| | | CPT | 0021A | Immunization administration by intramuscular injection of severe acute respiratory syndrome coronavirus 2 (SARS-CoV-2) (coronavirus disease [COVID-19]) vaccine, DNA, spike protein, chimpanzee adenovirus Oxford 1 (ChAdOx1) vector, preservative free, 5x1010 viral particles/0.5 mL dosage; first dose (deprecated 2024) |
| | *Janssen (Johnson & Johnson) viral vector vaccines* | CPT | 91303 | Severe acute respiratory syndrome coronavirus 2 (SARS-CoV-2) (coronavirus disease [COVID-19]) vaccine, DNA, spike protein, adenovirus type 26 (Ad26) vector, preservative free, 5x1010 viral particles/0.5 mL dosage, for intramuscular use (deprecated 2024) |
| | | CPT | 0031A | Immunization administration by intramuscular injection of severe acute respiratory syndrome coronavirus 2 (SARS-CoV-2) (coronavirus disease [COVID-19]) vaccine, DNA, spike protein, adenovirus type 26 (Ad26) vector, preservative free, 5x1010 viral particles/0.5 mL dosage; single dose (deprecated 2024) |
| | | CPT | 0034A | Immunization administration by intramuscular injection of severe acute respiratory syndrome coronavirus 2 (SARS-CoV-2) (coronavirus disease [COVID-19]) vaccine, DNA, spike protein, adenovirus type 26 (Ad26) vector, preservative free, 5x1010 viral particles/0.5 mL dosage; booster dose (deprecated 2024) |
| | *Moderna mRNA vaccines* | CPT | 91301 | Severe acute respiratory syndrome coronavirus 2 (SARS-CoV-2) (coronavirus disease [COVID-19]) vaccine, mRNA-LNP, spike protein, preservative free, 100 mcg/0.5 mL dosage, for intramuscular use (deprecated 2024) |
| | | CPT | 0011A | Immunization administration by intramuscular injection of severe acute respiratory syndrome coronavirus 2 (SARS-CoV-2) (coronavirus disease [COVID-19]) vaccine, mRNA-LNP, spike protein, preservative free, 100 mcg/0.5 mL dosage; first dose (deprecated 2024) |
| | | CPT | 0012A | Immunization administration by intramuscular injection of severe acute respiratory syndrome coronavirus 2 (SARS-CoV-2) (coronavirus disease [COVID-19]) vaccine, mRNA-LNP, spike protein, preservative free, 100 mcg/0.5 mL dosage; second dose (deprecated 2024) |

| | | | | |
|---|---|---|---|---|
| | | CPT | 0013A | Immunization administration by intramuscular injection of severe acute respiratory syndrome coronavirus 2 (SARS-CoV-2) (coronavirus disease [COVID-19]) vaccine, mRNA-LNP, spike protein, preservative free, 100 mcg/0.5 mL dosage; third dose (deprecated 2024) |
| | | CPT | 0094A | Immunization administration by intramuscular injection of severe acute respiratory syndrome coronavirus 2 (SARS-CoV-2) (coronavirus disease [COVID-19]) vaccine, mRNA-LNP, spike protein, preservative free, 50 mcg/0.5 mL dosage; booster dose, when administered to individuals 18 years and over (deprecated 2024) |
| | | CPT | 0134A | Immunization administration by intramuscular injection of severe acute respiratory syndrome coronavirus 2 (SARS-CoV-2) (coronavirus disease [COVID-19]) vaccine, mRNA-LNP, spike protein, bivalent, preservative free, 50 mcg/0.5 mL dosage, booster dose (deprecated 2024) |
| | *Novavax (Protein Subunit)* | CPT | 91304 | Severe acute respiratory syndrome coronavirus 2 (SARS-CoV-2) (coronavirus disease [COVID-19]) vaccine, recombinant spike protein nanoparticle, saponin-based adjuvant, 5 mcg/0.5 mL dosage, for intramuscular use |
| | *Other (Adjuvanted)* | CPT | 0042A | Immunization administration by intramuscular injection of severe acute respiratory syndrome coronavirus 2 (SARS-CoV-2) (coronavirus disease [COVID-19]) vaccine, recombinant spike protein nanoparticle, saponin-based adjuvant, preservative free, 5 mcg/0.5 mL dosage; second dose (deprecated 2024) |
| **Group 1: Naive (Uninfected/ Unvaccinated)** | **Exclusion** | | | |
| | COVID-19 infection | Various | See Group 2 | No documented COVID-19 infection (all codes from Group 2) |
| | COVID-19 vaccination | Various | See Group 3 | No documented COVID-19 vaccination (all codes from Group 3) |
| **Group 2: Infected, Unvaccinated** | **Inclusion** | | | |
| | COVID-19 diagnosis | ICD-10-CM | U07.1 | COVID-19 |
| | | ICD-10-CM | U07.2 | COVID-19, virus not identified (WHO) |
| | | ICD-10-CM | B97.2 | Coronavirus as the cause of diseases classified elsewhere |
| | | ICD-10-CM | B34.2 | Coronavirus infection, unspecified |
| | | ICD-10-CM | J12.81 | Pneumonia due to SARS-associated coronavirus |
| | | ICD-10-CM | J12.82 | Pneumonia due to coronavirus disease 2019 |
| | COVID-19 laboratory | LOINC | 94309-2 | SARS-CoV-2 RNA [Presence] in Specimen by NAA with probe detection (Positive) |
| | | LOINC | 94500-6 | SARS-CoV-2 RNA [Presence] in Respiratory system specimen by NAA with probe detection (Positive) |
| | | LOINC | 94308-4 | SARS-CoV-2 N gene [Presence] in Specimen by NAA using CDC primer-probe set N2 (Positive) |
| | | LOINC | 94533-7 | SARS-CoV-2 N gene [Presence] in Respiratory system specimen by NAA with probe detection (Positive) |

| | | | | |
|---|---|---|---|---|
| | | LOINC | 94534-5 | SARS-CoV-2 RdRp gene [Presence] in Respiratory system specimen by NAA with probe detection (Positive) |
| | | LOINC | 94559-2 | SARS-CoV-2 ORF1ab region [Presence] in Respiratory system specimen by NAA with probe detection (Positive) |
| | | LOINC | 94745-7 | SARS-CoV-2 RNA [Cycle Threshold #] in Respiratory system specimen by NAA with probe detection |
| | | LOINC | 94746-5 | SARS-CoV-2 RNA [Cycle Threshold #] in Specimen by NAA with probe detection |
| | | TriNetX | 9088 | SARS coronavirus 2 and related RNA [Presence] (Positive) |
| | | TriNetX | 9089 | SARS coronavirus 2 IgG IgM Ab [Presence] in Serum or Plasma (Positive) |
| | **Exclusion** | | | |
| | COVID-19 vaccination | Various | See Group 3 | No documented COVID-19 vaccination (all codes from Group 3) |
| **Group 3: Vaccinated Only (No Prior Infection)** | **Inclusion** | | | |
| | Pfizer-BioNTech mRNA vaccines | | | |
| | *First or second dose of monovalent vaccine* | CPT | 91309 | Severe acute respiratory syndrome coronavirus 2 (SARS-CoV-2) (coronavirus disease [COVID-19]) vaccine, mRNA-LNP, spike protein, preservative free, 50 mcg/0.5 mL dosage, for intramuscular use (deprecated 2024) |
| | | CPT | 91308 | Severe acute respiratory syndrome coronavirus 2 (SARS-CoV-2) (coronavirus disease [COVID-19]) vaccine, mRNA-LNP, spike protein, preservative free, 3 mcg/0.2 mL dosage, diluent reconstituted, tris-sucrose formulation, for intramuscular use (deprecated 2024) |
| | | CPT | 91305 | Severe acute respiratory syndrome coronavirus 2 (SARS-CoV-2) (coronavirus disease [COVID-19]) vaccine, mRNA-LNP, spike protein, preservative free, 30 mcg/0.3 mL dosage, tris-sucrose formulation, for intramuscular use (deprecated 2024) |
| | | CPT | 91306 | Severe acute respiratory syndrome coronavirus 2 (SARS-CoV-2) (coronavirus disease [COVID-19]) vaccine, mRNA-LNP, spike protein, preservative free, 50 mcg/0.25 mL dosage, for intramuscular use (deprecated 2024) |
| | | CPT | 91307 | Severe acute respiratory syndrome coronavirus 2 (SARS-CoV-2) (coronavirus disease [COVID-19]) vaccine, mRNA-LNP, spike protein, preservative free, 10 mcg/0.2 mL dosage, diluent reconstituted, tris-sucrose formulation, for intramuscular use (deprecated 2024) |
| | | CPT | 91300 | Severe acute respiratory syndrome coronavirus 2 (SARS-CoV-2) (coronavirus disease [COVID-19]) vaccine, mRNA-LNP, spike protein, preservative free, 30 mcg/0.3 mL dosage, diluent reconstituted, for intramuscular use (deprecated 2024) |
| | | CPT | 0001A | Immunization administration by intramuscular injection of severe acute respiratory syndrome |

| | | |
|---|---|---|
| | | coronavirus 2 (SARS-CoV-2) (coronavirus disease [COVID-19]) vaccine, mRNA-LNP, spike protein, preservative free, 30 mcg/0.3 mL dosage, diluent reconstituted; first dose (deprecated 2024) |
| CPT | 0002A | Immunization administration by intramuscular injection of severe acute respiratory syndrome coronavirus 2 (SARS-CoV-2) (coronavirus disease [COVID-19]) vaccine, mRNA-LNP, spike protein, preservative free, 30 mcg/0.3 mL dosage, diluent reconstituted; second dose (deprecated 2024) |
| CPT | 0111A | Immunization administration by intramuscular injection of severe acute respiratory syndrome coronavirus 2 (SARS-CoV-2) (coronavirus disease [COVID-19]) vaccine, mRNA-LNP, spike protein, preservative free, 25 mcg/0.25 mL dosage; first dose (deprecated 2024) |
| CPT | 0112A | Immunization administration by intramuscular injection of severe acute respiratory syndrome coronavirus 2 (SARS-CoV-2) (coronavirus disease [COVID-19]) vaccine, mRNA-LNP, spike protein, preservative free, 25 mcg/0.25 mL dosage; second dose (deprecated 2024) |
| CPT | 0081A | Immunization administration by intramuscular injection of severe acute respiratory syndrome coronavirus 2 (SARS-CoV-2) (coronavirus disease [COVID-19]) vaccine, mRNA-LNP, spike protein, preservative free, 3 mcg/0.2 mL dosage, diluent reconstituted, tris-sucrose formulation; first dose (deprecated 2024) |
| CPT | 0082A | Immunization administration by intramuscular injection of severe acute respiratory syndrome coronavirus 2 (SARS-CoV-2) (coronavirus disease [COVID-19]) vaccine, mRNA-LNP, spike protein, preservative free, 3 mcg/0.2 mL dosage, diluent reconstituted, tris-sucrose formulation; second dose (deprecated 2024) |
| CPT | 0071A | Immunization administration by intramuscular injection of severe acute respiratory syndrome coronavirus 2 (SARS-CoV-2) (coronavirus disease [COVID-19]) vaccine, mRNA-LNP, spike protein, preservative free, 10 mcg/0.2 mL dosage, diluent reconstituted, tris-sucrose formulation; first dose (deprecated 2024) |
| CPT | 0072A | Immunization administration by intramuscular injection of severe acute respiratory syndrome coronavirus 2 (SARS-CoV-2) (coronavirus disease [COVID-19]) vaccine, mRNA-LNP, spike protein, preservative free, 10 mcg/0.2 mL dosage, diluent reconstituted, tris-sucrose formulation; second dose (deprecated 2024) |
| CPT | 0051A | Immunization administration by intramuscular injection of severe acute respiratory syndrome coronavirus 2 (SARS-CoV-2) (coronavirus disease [COVID-19]) vaccine, mRNA-LNP, spike protein, preservative free, 30 mcg/0.3 mL |

| | | | |
|---|---|---|---|
| | | | dosage, tris-sucrose formulation; first dose (deprecated 2024) |
| | CPT | 0052A | Immunization administration by intramuscular injection of severe acute respiratory syndrome coronavirus 2 (SARS-CoV-2) (coronavirus disease [COVID-19]) vaccine, mRNA-LNP, spike protein, preservative free, 30 mcg/0.3 mL dosage, tris-sucrose formulation; second dose (deprecated 2024) |
| *Booster dose of monovalent vaccine* | CPT | 0053A | Immunization administration by intramuscular injection of severe acute respiratory syndrome coronavirus 2 (SARS-CoV-2) (coronavirus disease [COVID-19]) vaccine, mRNA-LNP, spike protein, preservative free, 30 mcg/0.3 mL dosage, tris-sucrose formulation; third dose (deprecated 2024) |
| | CPT | 0054A | Immunization administration by intramuscular injection of severe acute respiratory syndrome coronavirus 2 (SARS-CoV-2) (coronavirus disease [COVID-19]) vaccine, mRNA-LNP, spike protein, preservative free, 30 mcg/0.3 mL dosage, tris-sucrose formulation; booster dose (deprecated 2024) |
| | CPT | 0073A | Immunization administration by intramuscular injection of severe acute respiratory syndrome coronavirus 2 (SARS-CoV-2) (coronavirus disease [COVID-19]) vaccine, mRNA-LNP, spike protein, preservative free, 10 mcg/0.2 mL dosage, diluent reconstituted, tris-sucrose formulation; third dose (deprecated 2024) |
| | CPT | 0074A | Immunization administration by intramuscular injection of severe acute respiratory syndrome coronavirus 2 (SARS-CoV-2) (coronavirus disease [COVID-19]) vaccine, mRNA-LNP, spike protein, preservative free, 10 mcg/0.2 mL dosage, diluent reconstituted, tris-sucrose formulation; booster dose (deprecated 2024) |
| | CPT | 0094A | Immunization administration by intramuscular injection of severe acute respiratory syndrome coronavirus 2 (SARS-CoV-2) (coronavirus disease [COVID-19]) vaccine, mRNA-LNP, spike protein, preservative free, 50 mcg/0.5 mL dosage; booster dose, when administered to individuals 18 years and over (deprecated 2024) |
| | CPT | 0003A | Immunization administration by intramuscular injection of severe acute respiratory syndrome coronavirus 2 (SARS-CoV-2) (coronavirus disease [COVID-19]) vaccine, mRNA-LNP, spike protein, preservative free, 30 mcg/0.3 mL dosage, diluent reconstituted; third dose (deprecated 2024) |
| | CPT | 0004A | Immunization administration by intramuscular injection of severe acute respiratory syndrome coronavirus 2 (SARS-CoV-2) (coronavirus disease [COVID-19]) vaccine, mRNA-LNP, spike protein, preservative free, 30 mcg/0.3 mL dosage, diluent reconstituted; booster dose (deprecated 2024) |

| | | | |
|---|---|---|---|
| *Bivalent vaccine* | CPT | 0173A | Immunization administration by intramuscular injection of severe acute respiratory syndrome coronavirus 2 (SARS-CoV-2) (coronavirus disease [COVID-19]) vaccine, mRNA-LNP, bivalent spike protein, preservative free, 3 mcg/0.2 mL dosage, diluent reconstituted, tris-sucrose formulation, third dose |
| | CPT | 91314 | Severe acute respiratory syndrome coronavirus 2 (SARS-CoV-2) (coronavirus disease [COVID-19]) vaccine, mRNA-LNP, spike protein, bivalent, preservative free, 25 mcg/0.25 mL dosage, for intramuscular use (deprecated 2024) |
| | CPT | 91311 | Severe acute respiratory syndrome coronavirus 2 (SARS-CoV-2) (coronavirus disease [COVID-19]) vaccine, mRNA-LNP, spike protein, preservative free, 25 mcg/0.25 mL dosage, for intramuscular use (deprecated 2024) |
| | CPT | 91312 | Severe acute respiratory syndrome coronavirus 2 (SARS-CoV-2) (coronavirus disease [COVID-19]) vaccine, mRNA-LNP, bivalent spike protein, preservative free, 30 mcg/0.3 mL dosage, tris-sucrose formulation, for intramuscular use (deprecated 2024) |
| | CPT | 0164A | Immunization administration by intramuscular injection of severe acute respiratory syndrome coronavirus 2 (SARS-CoV-2) (coronavirus disease [COVID-19]) vaccine, mRNA-LNP, spike protein, bivalent, preservative free, 10 mcg/0.2 mL dosage, booster dose |
| | CPT | 0154A | Immunization administration by intramuscular injection of severe acute respiratory syndrome coronavirus 2 (SARS-CoV-2) (coronavirus disease [COVID-19]) vaccine, mRNA-LNP, bivalent spike protein, preservative free, 10 mcg/0.2 mL dosage, diluent reconstituted, tris-sucrose formulation, booster dose (deprecated 2024) |
| | CPT | 0144A | Immunization administration by intramuscular injection of severe acute respiratory syndrome coronavirus 2 (SARS-CoV-2) (coronavirus disease [COVID-19]) vaccine, mRNA-LNP, spike protein, bivalent, preservative free, 25 mcg/0.25 mL dosage, booster dose (deprecated 2024) |
| | CPT | 0124A | Immunization administration by intramuscular injection of severe acute respiratory syndrome coronavirus 2 (SARS-CoV-2) (coronavirus disease [COVID-19]) vaccine, mRNA-LNP, bivalent spike protein, preservative free, 30 mcg/0.3 mL dosage, tris-sucrose formulation, booster dose (deprecated 2024) |
| | CPT | 91315 | Severe acute respiratory syndrome coronavirus 2 (SARS-CoV-2) (coronavirus disease [COVID-19]) vaccine, mRNA-LNP, bivalent spike protein, preservative free, 10 mcg/0.2 mL dosage, diluent reconstituted, tris-sucrose formulation, for intramuscular use (deprecated 2024) |

| | | | | |
|---|---|---|---|---|
| | **Exclusion** | | | |
| | COVID-19 infection | Various | See Group 2 | No documented COVID-19 infection (all codes from Group 2) |
| **Group 4: Infected, Vaccinated** | **Inclusion** | | | |
| | COVID-19 infection | Various | See Group 2 | All COVID-19 diagnostic and laboratory codes from Group 2 |
| | COVID-19 vaccination | Various | See Group 3 | All COVID-19 vaccine codes from Group 3 |

All outcomes were defined using validated ICD-10-CM diagnosis codes, SNOMED CT concepts, CPT procedure codes, and laboratory values.

**Table S2.** Diagnostic and procedural code definitions for all 50 outcomes.

| Outcome | Term | System | Code | Description |
|---|---|---|---|---|
| Cardiovascular Events | Major adverse cardiovascular events | ICD-10-CM | I21 | Acute myocardial infarction |
| | | ICD-10-CM | I22 | Subsequent STEMI and NSTEMI |
| | | ICD-10-CM | I24 | Other acute ischemic heart diseases |
| | | ICD-10-CM | I25.5 | Ischemic cardiomyopathy |
| | | ICD-10-CM | I20 | Angina pectoris |
| | | ICD-10-CM | I20-I25 | Ischemic heart diseases |
| | | ICD-10-CM | I50 | Heart failure |
| | | ICD-10-CM | I42 | Cardiomyopathy |
| | | ICD-10-CM | I46 | Cardiac arrest |
| | | ICD-10-CM | R56.0 | Febrile convulsions |
| | | ICD-10-CM | R57.0 | Cardiogenic shock |
| | | ICD-10-CM | I63 | Cerebral infarction |
| | | ICD-10-CM | I65 | Occlusion/stenosis of precerebral arteries |
| | | ICD-10-CM | I66 | Occlusion/stenosis of cerebral arteries |
| | | ICD-10-CM | G45 | Transient cerebral ischemic attacks |
| | | ICD-10-CM | I60 | Nontraumatic subarachnoid hemorrhage |
| | | ICD-10-CM | I61 | Nontraumatic intracerebral hemorrhage |
| | Cardiovascular disease | ICD-10-CM | I20-I25 | Ischemic heart diseases |
| | | ICD-10-CM | I48 | Atrial fibrillation and flutter |
| | | ICD-10-CM | I47 | Paroxysmal tachycardia |
| | | ICD-10-CM | I49 | Other cardiac arrhythmias |
| | | ICD-10-CM | I50 | Heart failure |
| | | ICD-10-CM | I42 | Cardiomyopathy |
| | | ICD-10-CM | I46 | Cardiac arrest |
| | | ICD-10-CM | R57.0 | Cardiogenic shock |
| | | ICD-10-CM | I30 | Acute pericarditis |
| | | ICD-10-CM | I31 | Other diseases of pericardium |
| | | ICD-10-CM | I32 | Pericarditis in diseases classified elsewhere |
| | | ICD-10-CM | I40 | Acute myocarditis |
| | | ICD-10-CM | I41 | Myocarditis in diseases classified elsewhere |
| | | ICD-10-CM | I51.4 | Myocarditis, unspecified |
| | | ICD-10-CM | I11 | Hypertensive heart disease |
| | | ICD-10-CM | I26 | Pulmonary embolism |
| | | ICD-10-CM | I82 | Other venous embolism and thrombosis |
| | | ICD-10-CM | D75.84 | Platelet-activating anti-PF4 disorders |
| | Ischemic heart disease | ICD-10-CM | I20-I25 | Ischemic heart diseases |
| | Angina | ICD-10-CM | I20 | Angina pectoris |
| | Acute coronary syndrome | ICD-10-CM | I24 | Other acute ischemic heart diseases |
| | Myocardial infarction | ICD-10-CM | I21 | Acute myocardial infarction |
| | | ICD-10-CM | I22 | Subsequent STEMI and NSTEMI |
| | Ischemic cardiomyopathy | ICD-10-CM | I25.5 | Ischemic cardiomyopathy |
| | Other cardiac diseases | ICD-10-CM | I51 | Complications and ill-defined heart disease |
| Cardiac Arrhythmias | Arrhythmias (any) | ICD-10-CM | I48 | Atrial fibrillation and flutter |
| | | ICD-10-CM | I47 | Paroxysmal tachycardia |
| | | ICD-10-CM | I49 | Other cardiac arrhythmias |
| | | ICD-10-CM | R00 | Abnormalities of heart beat |
| | Atrial fibrillation and flutter | ICD-10-CM | I48 | Atrial fibrillation and flutter |

| | | | | |
|---|---|---|---|---|
| | Paroxysmal tachycardia | ICD-10-CM | I47 | Paroxysmal tachycardia |
| | Other cardiac arrhythmias | ICD-10-CM | I49 | Other cardiac arrhythmias |
| Inflammatory Heart Disease | Inflammatory heart disease | ICD-10-CM | I30 | Acute pericarditis |
| | | ICD-10-CM | I31 | Other diseases of pericardium |
| | | ICD-10-CM | I32 | Pericarditis in diseases classified elsewhere |
| | | ICD-10-CM | I40 | Acute myocarditis |
| | | ICD-10-CM | I41 | Myocarditis in diseases classified elsewhere |
| | | ICD-10-CM | I51.4 | Myocarditis, unspecified |
| | | ICD-10-CM | I38 | Endocarditis, valve unspecified |
| | | ICD-10-CM | I39 | Endocarditis in diseases classified elsewhere |
| | Pericarditis | ICD-10-CM | I30 | Acute pericarditis |
| | | ICD-10-CM | I31 | Other diseases of pericardium |
| | | ICD-10-CM | I32 | Pericarditis in diseases classified elsewhere |
| | Myocarditis | ICD-10-CM | I40 | Acute myocarditis |
| | | ICD-10-CM | I41 | Myocarditis in diseases classified elsewhere |
| | | ICD-10-CM | I51.4 | Myocarditis, unspecified |
| | Endocarditis | ICD-10-CM | I38 | Endocarditis, valve unspecified |
| | | ICD-10-CM | I39 | Endocarditis in diseases classified elsewhere |
| Heart Failure and Cardiomyopathy | Hypertensive heart disease | ICD-10-CM | I11 | Hypertensive heart disease |
| | Pulmonary heart diseases | ICD-10-CM | I26 | Pulmonary embolism |
| | | ICD-10-CM | I27 | Other pulmonary heart diseases |
| | | ICD-10-CM | I28 | Other diseases of pulmonary vessels |
| | Heart failure | ICD-10-CM | I50 | Heart failure |
| | Cardiomyopathy | ICD-10-CM | I42 | Cardiomyopathy |
| | Cardiomegaly | ICD-10-CM | I51.7 | Cardiomegaly |
| | Cardiogenic shock | ICD-10-CM | R57.0 | Cardiogenic shock |
| | Cardiac arrest | ICD-10-CM | I46 | Cardiac arrest |
| Thrombotic Events | Thrombotic disorders | ICD-10-CM | I26 | Pulmonary embolism |
| | | ICD-10-CM | I82 | Other venous embolism and thrombosis |
| | | ICD-10-CM | D75.84 | Platelet-activating anti-PF4 disorders |
| | Pulmonary embolism | ICD-10-CM | I26 | Pulmonary embolism |
| | Deep vein thrombosis | ICD-10-CM | I82 | Other venous embolism and thrombosis |
| | Thrombosis with thrombocytopenia | ICD-10-CM | D75.84 | Platelet-activating anti-PF4 disorders |
| Cerebrovascular Events | Cerebrovascular disease | ICD-10-CM | I60-I69 | Cerebrovascular diseases |
| | | ICD-10-CM | G45 | Transient cerebral ischemic attacks |
| | Transient ischemic attacks | ICD-10-CM | G45 | Transient cerebral ischemic attacks |
| | Cerebral infarction | ICD-10-CM | I63 | Cerebral infarction |
| | Cerebral occlusion/stenosis | ICD-10-CM | I65 | Occlusion/stenosis of precerebral arteries |
| | | ICD-10-CM | I66 | Occlusion/stenosis of cerebral arteries |
| | Intracerebral hemorrhage | ICD-10-CM | I61 | Nontraumatic intracerebral hemorrhage |
| | Subarachnoid hemorrhage | ICD-10-CM | I60 | Nontraumatic subarachnoid hemorrhage |
| Lipid Profile | Dyslipidemia | ICD-10-CM | E78 | Disorders of lipoprotein metabolism |
| | Triglycerides ≥150 mg/dL | LOINC | 2571-8 | Triglyceride [Mass/volume] in Serum or Plasma |
| | | LOINC | 3043-7 | Triglyceride [Mass/volume] in Blood |

| | | | | |
|---|---|---|---|---|
| | | LOINC | 12951-0 | Triglyceride [Mass/volume] in Capillary blood |
| | Total cholesterol ≥200 mg/dL | LOINC | 2093-3 | Cholesterol [Mass/volume] in Serum or Plasma |
| | | LOINC | 35200-5 | Cholesterol [Mass/volume] in Serum or Plasma by VAP |
| | LDL cholesterol ≥130 mg/dL | LOINC | 2089-1 | Cholesterol in LDL [Mass/volume] in Serum or Plasma |
| | | LOINC | 13457-7 | Cholesterol in LDL [Mass/volume] in Serum or Plasma by calculation |
| | | LOINC | 18262-6 | Cholesterol in LDL [Mass/volume] in Serum or Plasma by Direct assay |
| | HDL cholesterol <50 mg/dL | LOINC | 2085-9 | Cholesterol in HDL [Mass/volume] in Serum or Plasma |
| Cardiac Biomarkers | Elevated Troponin I | LOINC | 10839-9 | Troponin I.cardiac [Mass/volume] in Serum or Plasma |
| | | LOINC | 42757-5 | Troponin I.cardiac [Mass/volume] in Blood |
| | | LOINC | 89579-7 | Troponin I.cardiac [Mass/volume] in Serum or Plasma by High sensitivity method |
| | | LOINC | 6598-7 | Troponin I.cardiac [Mass/volume] in Serum or Plasma by Detection limit <= 0.01 ng/mL |
| | Elevated Natriuretic peptide B/prohormone | LOINC | 42637-9 | Natriuretic peptide B [Mass/volume] in Blood |
| | | LOINC | 30934-4 | Natriuretic peptide B [Mass/volume] in Serum or Plasma |
| | | LOINC | 35257-5 | Natriuretic peptide B [Mass or Moles/volume] in Serum or Plasma |
| | | LOINC | 33762-6 | NT-proBNP [Mass/volume] in Serum or Plasma |
| | | LOINC | 83107-3 | NT-proBNP [Mass/volume] in Serum or Plasma by Immunoassay |
| | | LOINC | 33763-4 | NT-proBNP [Moles/volume] in Serum or Plasma |
| Healthcare Utilization | Hospital admissions | Visit Type | Inpatient | Inpatient hospital admission |
| | Emergency department visits | Visit Type | Emergency | Emergency department encounter |
| | Outpatient encounters | Visit Type | Outpatient | Outpatient clinic visit |
| | Intensive care unit admission | SNOMED | 305351004 | Admission to intensive care unit |
| | | SNOMED | 309904001 | Intensive care unit |
| | | SNOMED | 309905000 | Admission to critical care unit |
| | Mechanical ventilation | ICD-10-PCS | 5A09 | Assistance with Respiratory Ventilation |
| | | ICD-10-PCS | 5A19 | Performance of Respiratory Ventilation |
| | | CPT | 94002 | Ventilation assist and management, initiation of pressure or volume preset ventilators |
| | | CPT | 94003 | Ventilation assist and management, hospital inpatient/observation, subsequent days |
| | | CPT | 94004 | Ventilation assist and management, nursing facility, per day |
| | | CPT | 94005 | Home ventilator management care plan oversight |
| | Extracorporeal membrane oxygenation | CPT | 33946-33989 | Extracorporeal membrane oxygenation (ECMO) procedures |
| | | ICD-10-PCS | 5A15 | Extracorporeal oxygenation |
| | Cardiology referrals | SNOMED | 183856001 | Referral to cardiology service |
| | | SNOMED | 183857005 | Referral to cardiologist |

| | | | | |
|---|---|---|---|---|
| | Neurology referrals | SNOMED | 183866006 | Referral to neurology service |
| | Referral to NEJM | Custom | N/A | Study-specific referral query |
| Mortality | All-cause mortality | Demographics | Deceased | Deceased status indicator |
| | | ICD-10-CM | R99 | Ill-defined and unknown cause of mortality |

International Classification of Diseases, Tenth Revision, Clinical Modification (ICD-10-CM), Systematized Nomenclature of Medicine Clinical Terms (SNOMED CT), Logical Observation Identifiers Names and Codes (LOINC), and Current Procedural Terminology (CPT) codes used to define study outcomes.

**Table S3.** Outcomes in male infected, unvaccinated individuals compared to naive controls (Group 2 vs Group 1) across four post-exposure time windows.

| Time | Outcome | Group 2 (n=848,176) | Group 1 (n=11,954,643) | *p*-value | RR (95%CI) |
|---|---|---|---|---|---|
| | **Follow-up (months)** | **22 ± 16.2** | **22.7 ± 17.7** | | |
| 0-3 mo | Major adverse cardiovascular events | 9663 (1.1%) | 140795 (1.2%) | 0.001 | 0.97 (0.95-0.99) |
| | Cardiovascular disease | 14202 (1.8%) | 180094 (1.5%) | <0.001 | 1.14 (1.12-1.16) |
| | Ischemic heart disease | 6710 (0.8%) | 92874 (0.8%) | 0.795 | 1 (0.97-1.02) |
| | Myocardial angina | 610 (0.1%) | 9759 (0.1%) | <0.001 | 0.83 (0.76-0.9) |
| | Acute coronary disease | 1222 (0.1%) | 5504 (0%) | <0.001 | 2.94 (2.77-3.13) |
| | Myocardial infarction | 3121 (0.3%) | 21770 (0.2%) | <0.001 | 1.92 (1.85-1.99) |
| | Ischemic cardiomyopathy | 926 (0.1%) | 9681 (0.1%) | <0.001 | 1.27 (1.18-1.35) |
| | Arrythmias | 15915 (2%) | 160962 (1.4%) | <0.001 | 1.45 (1.42-1.47) |
| | Atrial fibrillation and flutter | 4822 (0.5%) | 50869 (0.4%) | <0.001 | 1.28 (1.25-1.32) |
| | Paroxysmal tachycardia | 2557 (0.3%) | 18336 (0.1%) | <0.001 | 1.86 (1.78-1.94) |
| | Other cardiac arrythmias | 6379 (0.7%) | 53063 (0.4%) | <0.001 | 1.64 (1.59-1.68) |
| | Inflammatory heart disease | 2327 (0.2%) | 13209 (0.1%) | <0.001 | 2.34 (2.24-2.44) |
| | Pericarditis | 1495 (0.2%) | 8668 (0.1%) | <0.001 | 2.28 (2.16-2.41) |
| | Myocarditis | 279 (0%) | 829 (0%) | <0.001 | 4.44 (3.87-5.08) |
| | Endocarditis | 752 (0.1%) | 4448 (0%) | <0.001 | 2.23 (2.07-2.41) |
| | Hypertensive heart disease | 12479 (1.8%) | 290424 (2.7%) | <0.001 | 0.66 (0.65-0.67) |
| | Pulmonary heart diseases | 5033 (0.5%) | 23437 (0.2%) | <0.001 | 2.88 (2.79-2.96) |
| | Heart Failure | 4924 (0.5%) | 46519 (0.4%) | <0.001 | 1.43 (1.39-1.48) |
| | Cardiomyopathy | 2362 (0.2%) | 24400 (0.2%) | <0.001 | 1.29 (1.24-1.35) |
| | Cardiomegaly | 5574 (0.6%) | 34686 (0.3%) | <0.001 | 2.16 (2.1-2.22) |
| | Cardiogenic shock | 752 (0.1%) | 4453 (0%) | <0.001 | 2.23 (2.06-2.41) |
| | Cardiac arrest | 1832 (0.2%) | 9581 (0.1%) | <0.001 | 2.53 (2.4-2.66) |
| | Thrombotic disorders | 6092 (0.6%) | 28124 (0.2%) | <0.001 | 2.91 (2.83-2.99) |
| | Pulmonary embolism | 3384 (0.4%) | 10927 (0.1%) | <0.001 | 4.12 (3.96-4.28) |
| | Deep and superficial thrombosis | 3921 (0.4%) | 21826 (0.2%) | <0.001 | 2.4 (2.32-2.48) |
| | Thrombosis with Thrombocytopenia | 0 (0%) | 10 (0%) | 0.384 | NA |
| | Elevated Troponin I | 4359 (0.5%) | 22633 (0.2%) | <0.001 | 2.57 (2.49-2.66) |
| | Elevated Natriuretic peptide B | 10193 (1.1%) | 44289 (0.4%) | <0.001 | 3.14 (3.07-3.21) |
| | Dyslipidemia | 21348 (3.5%) | 431563 (4.2%) | <0.001 | 0.83 (0.82-0.84) |
| | Triglyceride ≥150 mg/dl | 12159 (1.5%) | 131864 (1.1%) | <0.001 | 1.33 (1.31-1.36) |
| | Total cholesterol ≥200 mg/dl | 6476 (0.8%) | 126853 (1.1%) | <0.001 | 0.73 (0.71-0.75) |
| | LDL-cholesterol ≥130 mg/dl | 5758 (0.7%) | 109241 (0.9%) | <0.001 | 0.74 (0.72-0.76) |
| | HDL-cholesterol <50 mg/dl | 18373 (2.5%) | 276818 (2.5%) | <0.001 | 1.03 (1.02-1.05) |
| | Cerebrovascular disease | 4289 (0.5%) | 55073 (0.4%) | 0.001 | 1.05 (1.02-1.09) |
| | Transient ischemic attacks | 771 (0.1%) | 8266 (0.1%) | <0.001 | 1.24 (1.15-1.33) |
| | Cerebral infarction | 2146 (0.2%) | 22595 (0.2%) | <0.001 | 1.27 (1.21-1.32) |
| | Cerebral occlusion without infarction | 1543 (0.2%) | 19444 (0.2%) | 0.041 | 1.06 (1-1.11) |
| | Intracerebral hemorrhage | 585 (0.1%) | 6089 (0%) | <0.001 | 1.27 (1.16-1.38) |
| | Subarachnoid hemorrhage | 329 (0%) | 4263 (0%) | 0.762 | 1.02 (0.91-1.14) |
| | Hospital admission | 19728 (2.9%) | 378635 (3.6%) | <0.001 | 0.79 (0.78-0.8) |
| | Emergency visits | 18523 (4.5%) | 300448 (3.7%) | <0.001 | 1.22 (1.2-1.24) |
| | Outpatient encounters | 23366 (4.8%) | 912680 (10.9%) | <0.001 | 0.44 (0.44-0.45) |
| | Intensive care unit admission | 1744 (0.2%) | 37232 (0.3%) | <0.001 | 0.63 (0.6-0.66) |
| | Mechanical ventilation | 10711 (1.1%) | 36860 (0.3%) | <0.001 | 3.88 (3.8-3.97) |
| | Referral to Cardio services | 10 (0%) | 22 (0%) | <0.001 | 5.99 (2.84-12.6) |
| | Referral to Neuro services | 26 (0%) | 247 (0%) | 0.111 | 1.39 (0.93- 2.08) |
| | ECMO | 289 (0%) | 645 (0%) | <0.001 | 5.9 (5.14-6.78) |
| | ECMO-VV | 240 (0%) | 233 (0%) | <0.001 | 13.57 (11.3-16.2) |
| | ECMO-VA | 98 (0%) | 450 (0%) | <0.001 | 2.87 (2.31-3.57) |
| | Mortality | 25841 (2.7%) | 75197 (0.6%) | <0.001 | 4.53 (4.46-4.59) |
| 3-6mo | Major adverse cardiovascular events | 3424 (0.4%) | 47122 (0.4%) | 0.184 | 1.02 (0.99-1.06) |

| | | | | | |
|---|---|---|---|---|---|
| | Cardiovascular disease | 4189 (0.5%) | 59611 (0.5%) | 0.24 | 1.02 (0.99-1.05) |
| | Ischemic heart disease | 2392 (0.3%) | 31336 (0.3%) | 0.015 | 1.05 (1.01-1.1) |
| | Myocardial angina | 337 (0%) | 3335 (0%) | <0.001 | 1.34 (1.2-1.5) |
| | Acute coronary disease | 386 (0%) | 2340 (0%) | <0.001 | 2.19 (1.97-2.44) |
| | Myocardial infarction | 961 (0.1%) | 7522 (0.1%) | <0.001 | 1.71 (1.6-1.83) |
| | Ischemic cardiomyopathy | 323 (0%) | 3455 (0%) | <0.001 | 1.24 (1.1-1.39) |
| | Arrythmias | 5337 (0.7%) | 60040 (0.5%) | <0.001 | 1.31 (1.27-1.34) |
| | Atrial fibrillation and flutter | 1375 (0.1%) | 17589 (0.1%) | 0.042 | 1.06 (1-1.12) |
| | Paroxysmal tachycardia | 825 (0.1%) | 7167 (0.1%) | <0.001 | 1.53 (1.43-1.65) |
| | Other cardiac arrythmias | 1995 (0.2%) | 19012 (0.2%) | <0.001 | 1.43 (1.37-1.5) |
| | Inflammatory heart disease | 632 (0.1%) | 4564 (0%) | <0.001 | 1.84 (1.69-2) |
| | Pericarditis | 418 (0%) | 3055 (0%) | <0.001 | 1.81 (1.64-2.01) |
| | Myocarditis | 46 (0%) | 217 (0%) | <0.001 | 2.8 (2.03-3.84) |
| | Endocarditis | 224 (0%) | 1520 (0%) | <0.001 | 1.95 (1.69-2.24) |
| | Hypertensive heart disease | 5242 (0.8%) | 97351 (0.9%) | <0.001 | 0.82 (0.8-0.84) |
| | Pulmonary heart diseases | 1097 (0.1%) | 8846 (0.1%) | <0.001 | 1.67 (1.57-1.77) |
| | Heart Failure | 1593 (0.2%) | 16899 (0.1%) | <0.001 | 1.28 (1.22-1.35) |
| | Cardiomyopathy | 750 (0.1%) | 8490 (0.1%) | <0.001 | 1.18 (1.09-1.27) |
| | Cardiomegaly | 1329 (0.1%) | 10479 (0.1%) | <0.001 | 1.71 (1.61-1.81) |
| | Cardiogenic shock | 236 (0%) | 1529 (0%) | <0.001 | 2.04 (1.78-2.34) |
| | Cardiac arrest | 557 (0.1%) | 3547 (0%) | <0.001 | 2.08 (1.9-2.27) |
| | Thrombotic disorders | 1121 (0.1%) | 10454 (0.1%) | <0.001 | 1.45 (1.36-1.54) |
| | Pulmonary embolism | 489 (0.1%) | 4010 (0%) | <0.001 | 1.63 (1.48-1.79) |
| | Deep and superficial thrombosis | 891 (0.1%) | 8091 (0.1%) | <0.001 | 1.47 (1.38-1.58) |
| | Thrombosis with Thrombocytopenia | 0 (0%) | 10 (0%) | 0.384 | NA |
| | Elevated Troponin I | 1134 (0.1%) | 7063 (0.1%) | <0.001 | 2.15 (2.02-2.29) |
| | Elevated Natriuretic peptide B | 1627 (0.2%) | 15043 (0.1%) | <0.001 | 1.49 (1.41-1.56) |
| | Dyslipidemia | 8318 (1.4%) | 126938 (1.3%) | <0.001 | 1.1 (1.07-1.12) |
| | Triglyceride ≥150 mg/dl | 4393 (0.5%) | 41471 (0.4%) | <0.001 | 1.54 (1.49-1.58) |
| | Total cholesterol ≥200 mg/dl | 4080 (0.5%) | 37603 (0.3%) | <0.001 | 1.55 (1.5-1.6) |
| | LDL-cholesterol ≥130 mg/dl | 3591 (0.4%) | 31978 (0.3%) | <0.001 | 1.58 (1.53-1.64) |
| | HDL-cholesterol <50 mg/dl | 7006 (1%) | 77765 (0.7%) | <0.001 | 1.4 (1.37-1.44) |
| | Cerebrovascular disease | 1711 (0.2%) | 19007 (0.2%) | <0.001 | 1.22 (1.16-1.28) |
| | Transient ischemic attacks | 414 (0%) | 3294 (0%) | <0.001 | 1.67 (1.51-1.85) |
| | Cerebral infarction | 732 (0.1%) | 7877 (0.1%) | <0.001 | 1.24 (1.15-1.34) |
| | Cerebral occlusion without infarction | 603 (0.1%) | 6604 (0.1%) | <0.001 | 1.22 (1.12-1.32) |
| | Intracerebral hemorrhage | 177 (0%) | 1625 (0%) | <0.001 | 1.44 (1.23-1.68) |
| | Subarachnoid hemorrhage | 109 (0%) | 1045 (0%) | 0.001 | 1.38 (1.13-1.68) |
| | Hospital admission | 5600 (0.8%) | 104777 (1%) | <0.001 | 0.81 (0.78-0.83) |
| | Emergency visits | 8228 (2.1%) | 143424 (1.8%) | <0.001 | 1.15 (1.12-1.17) |
| | Outpatient encounters | 9303 (2%) | 227398 (3.1%) | <0.001 | 0.66 (0.65-0.68) |
| | Intensive care unit admission | 516 (0.1%) | 14079 (0.1%) | <0.001 | 0.49 (0.45-0.54) |
| | Mechanical ventilation | 1084 (0.1%) | 9667 (0.1%) | <0.001 | 1.51 (1.42-1.61) |
| | Referral to Cardio services | 0 (0%) | 10 (0%) | 0.384 | NA |
| | Referral to Neuro services | 13 (0%) | 154 (0%) | 0.713 | 1.11 (0.63-1.96) |
| | ECMO | 11 (0%) | 121 (0%) | 0.566 | 1.2 (0.65-2.22) |
| | ECMO-VV | 10 (0%) | 42 (0%) | 0.001 | 3.14 (1.57-6.25) |
| | ECMO-VA | 10 (0%) | 88 (0%) | 0.224 | 1.5 (0.78-2.88) |
| | Mortality | 5285 (0.5%) | 37694 (0.3%) | <0.001 | 1.85 (1.79-1.9) |
| 6-9mo | Major adverse cardiovascular events | 2743 (0.3%) | 36661 (0.3%) | 0.008 | 1.05 (1.01-1.1) |
| | Cardiovascular disease | 3398 (0.4%) | 45670 (0.4%) | <0.001 | 1.08 (1.04-1.12) |
| | Ischemic heart disease | 1863 (0.2%) | 24258 (0.2%) | 0.016 | 1.06 (1.01-1.11) |
| | Myocardial angina | 293 (0%) | 2671 (0%) | <0.001 | 1.45 (1.29-1.64) |
| | Acute coronary disease | 290 (0%) | 1981 (0%) | <0.001 | 1.94 (1.72-2.2) |
| | Myocardial infarction | 678 (0.1%) | 6546 (0.1%) | <0.001 | 1.39 (1.28-1.5) |
| | Ischemic cardiomyopathy | 245 (0%) | 2460 (0%) | <0.001 | 1.32 (1.16-1.5) |

| | | | | | |
|---|---|---|---|---|---|
| | Arrythmias | 4539 (0.6%) | 48207 (0.4%) | <0.001 | 1.39 (1.35-1.43) |
| | Atrial fibrillation and flutter | 1063 (0.1%) | 13527 (0.1%) | 0.051 | 1.06 (1-1.13) |
| | Paroxysmal tachycardia | 650 (0.1%) | 5395 (0%) | <0.001 | 1.61 (1.48-1.74) |
| | Other cardiac arrythmias | 1632 (0.2%) | 14865 (0.1%) | <0.001 | 1.5 (1.43-1.58) |
| | Inflammatory heart disease | 416 (0%) | 3579 (0%) | <0.001 | 1.55 (1.4-1.71) |
| | Pericarditis | 271 (0%) | 2380 (0%) | <0.001 | 1.51 (1.33-1.71) |
| | Myocarditis | 33 (0%) | 194 (0%) | <0.001 | 2.24 (1.55-3.24) |
| | Endocarditis | 153 (0%) | 1199 (0%) | <0.001 | 1.69 (1.43-2) |
| | Hypertensive heart disease | 4286 (0.6%) | 73291 (0.7%) | <0.001 | 0.89 (0.86-0.92) |
| | Pulmonary heart diseases | 764 (0.1%) | 6783 (0.1%) | <0.001 | 1.51 (1.41-1.63) |
| | Heart Failure | 1204 (0.1%) | 12729 (0.1%) | <0.001 | 1.28 (1.21-1.36) |
| | Cardiomyopathy | 581 (0.1%) | 6285 (0%) | <0.001 | 1.23 (1.13-1.34) |
| | Cardiomegaly | 965 (0.1%) | 8470 (0.1%) | <0.001 | 1.53 (1.44-1.64) |
| | Cardiogenic shock | 179 (0%) | 1231 (0%) | <0.001 | 1.92 (1.64-2.25) |
| | Cardiac arrest | 345 (0%) | 3007 (0%) | <0.001 | 1.52 (1.36-1.7) |
| | Thrombotic disorders | 864 (0.1%) | 8057 (0.1%) | <0.001 | 1.45 (1.35-1.55) |
| | Pulmonary embolism | 360 (0%) | 3123 (0%) | <0.001 | 1.54 (1.38-1.71) |
| | Deep and superficial thrombosis | 688 (0.1%) | 6306 (0%) | <0.001 | 1.46 (1.35-1.58) |
| | Thrombosis with Thrombocytopenia | 0 (0%) | 0 (0%) | NA | NA |
| | Elevated Troponin I | 946 (0.1%) | 6284 (0%) | <0.001 | 2.02 (1.88-2.16) |
| | Elevated Natriuretic peptide B | 1256 (0.1%) | 12212 (0.1%) | <0.001 | 1.41 (1.33-1.5) |
| | Dyslipidemia | 6953 (1.2%) | 97409 (1%) | <0.001 | 1.2 (1.17-1.22) |
| | Triglyceride ≥150 mg/dl | 3803 (0.5%) | 35202 (0.3%) | <0.001 | 1.57 (1.52-1.62) |
| | Total cholesterol ≥200 mg/dl | 3642 (0.4%) | 31943 (0.3%) | <0.001 | 1.63 (1.57-1.68) |
| | LDL-cholesterol ≥130 mg/dl | 3108 (0.4%) | 27497 (0.2%) | <0.001 | 1.59 (1.54-1.65) |
| | HDL-cholesterol <50 mg/dl | 5965 (0.9%) | 63027 (0.6%) | <0.001 | 1.48 (1.44-1.52) |
| | Cerebrovascular disease | 1426 (0.2%) | 15183 (0.1%) | <0.001 | 1.27 (1.21-1.34) |
| | Transient ischemic attacks | 365 (0%) | 2799 (0%) | <0.001 | 1.73 (1.55-1.93) |
| | Cerebral infarction | 625 (0.1%) | 6186 (0%) | <0.001 | 1.35 (1.24-1.46) |
| | Cerebral occlusion without infarction | 537 (0.1%) | 5365 (0%) | <0.001 | 1.33 (1.22-1.46) |
| | Intracerebral hemorrhage | 149 (0%) | 1268 (0%) | <0.001 | 1.55 (1.31-1.84) |
| | Subarachnoid hemorrhage | 101 (0%) | 916 (0%) | <0.001 | 1.45 (1.18-1.79) |
| | Hospital admission | 4858 (0.7%) | 71652 (0.7%) | 0.202 | 1.02 (0.99-1.05) |
| | Emergency visits | 7192 (1.8%) | 121312 (1.6%) | <0.001 | 1.19 (1.16-1.22) |
| | Outpatient encounters | 7169 (1.6%) | 146973 (2%) | <0.001 | 0.78 (0.76-0.8) |
| | Intensive care unit admission | 419 (0%) | 10499 (0.1%) | <0.001 | 0.54 (0.49-0.59) |
| | Mechanical ventilation | 830 (0.1%) | 7376 (0.1%) | <0.001 | 1.52 (1.41-1.63) |
| | Referral to Cardio services | 0 (0%) | 10 (0%) | 0.384 | NA |
| | Referral to Neuro services | 10 (0%) | 141 (0%) | 0.835 | 0.93 (0.49-1.77) |
| | ECMO | 10 (0%) | 98 (0%) | 0.371 | 1.35 (0.7-2.58) |
| | ECMO-VV | 10 (0%) | 36 (0%) | <0.001 | 3.66 (1.82-7.38) |
| | ECMO-VA | 10 (0%) | 66 (0%) | 0.038 | 2 (1.03-3.88) |
| | Mortality | 3884 (0.4%) | 33172 (0.3%) | <0.001 | 1.54 (1.49-1.59) |
| >9mo | Major adverse cardiovascular events | 14065 (1.7%) | 221027 (1.9%) | <0.001 | 0.9 (0.88-0.91) |
| | Cardiovascular disease | 17599 (2.2%) | 277180 (2.4%) | <0.001 | 0.92 (0.91-0.94) |
| | Ischemic heart disease | 9374 (1.1%) | 144237 (1.2%) | <0.001 | 0.9 (0.88-0.92) |
| | Myocardial angina | 1388 (0.1%) | 17045 (0.1%) | 0.006 | 1.08 (1.02-1.14) |
| | Acute coronary disease | 1205 (0.1%) | 11663 (0.1%) | <0.001 | 1.37 (1.29-1.45) |
| | Myocardial infarction | 3323 (0.4%) | 40152 (0.3%) | <0.001 | 1.11 (1.07-1.15) |
| | Ischemic cardiomyopathy | 983 (0.1%) | 13841 (0.1%) | 0.062 | 0.94 (0.88-1) |
| | Arrythmias | 23914 (3%) | 310775 (2.7%) | <0.001 | 1.14 (1.12-1.15) |
| | Atrial fibrillation and flutter | 4846 (0.5%) | 79539 (0.6%) | <0.001 | 0.83 (0.8-0.85) |
| | Paroxysmal tachycardia | 2823 (0.3%) | 33364 (0.3%) | <0.001 | 1.13 (1.09-1.17) |
| | Other cardiac arrythmias | 7831 (0.9%) | 94494 (0.8%) | <0.001 | 1.13 (1.11-1.16) |
| | Inflammatory heart disease | 1949 (0.2%) | 20705 (0.2%) | <0.001 | 1.25 (1.2-1.31) |
| | Pericarditis | 1275 (0.1%) | 13154 (0.1%) | <0.001 | 1.28 (1.21-1.36) |

| | | | | | |
|---|---|---|---|---|---|
| | Myocarditis | 127 (0%) | 1015 (0%) | <0.001 | 1.65 (1.37-1.98) |
| | Endocarditis | 719 (0.1%) | 7613 (0.1%) | <0.001 | 1.25 (1.16-1.35) |
| | Hypertensive heart disease | 27983 (4.1%) | 472662 (4.5%) | <0.001 | 0.9 (0.89-0.91) |
| | Pulmonary heart diseases | 3387 (0.4%) | 40376 (0.3%) | <0.001 | 1.13 (1.09-1.17) |
| | Heart Failure | 5371 (0.6%) | 76587 (0.6%) | <0.001 | 0.95 (0.93-0.98) |
| | Cardiomyopathy | 2448 (0.3%) | 35274 (0.3%) | <0.001 | 0.93 (0.89-0.96) |
| | Cardiomegaly | 4900 (0.5%) | 54452 (0.4%) | <0.001 | 1.21 (1.18-1.25) |
| | Cardiogenic shock | 791 (0.1%) | 8051 (0.1%) | <0.001 | 1.3 (1.21-1.4) |
| | Cardiac arrest | 2173 (0.2%) | 20768 (0.2%) | <0.001 | 1.39 (1.33-1.45) |
| | Thrombotic disorders | 4410 (0.5%) | 50647 (0.4%) | <0.001 | 1.17 (1.14-1.21) |
| | Pulmonary embolism | 1570 (0.2%) | 18064 (0.1%) | <0.001 | 1.16 (1.1-1.22) |
| | Deep and superficial thrombosis | 3672 (0.4%) | 40599 (0.3%) | <0.001 | 1.21 (1.17-1.25) |
| | Thrombosis with Thrombocytopenia | 0 (0%) | 13 (0%) | 0.32 | NA |
| | Elevated Troponin I | 6176 (0.7%) | 54967 (0.4%) | <0.001 | 1.51 (1.47-1.55) |
| | Elevated Natriuretic peptide B | 6156 (0.7%) | 84548 (0.7%) | 0.934 | 1 (0.98-1.03) |
| | Dyslipidemia | 44346 (7.8%) | 690579 (7.2%) | <0.001 | 1.08 (1.07-1.09) |
| | Triglyceride ≥150 mg/dl | 25611 (3.2%) | 287954 (2.5%) | <0.001 | 1.29 (1.28-1.31) |
| | Total cholesterol ≥200 mg/dl | 24812 (3%) | 283101 (2.4%) | <0.001 | 1.25 (1.24-1.27) |
| | LDL-cholesterol ≥130 mg/dl | 22463 (2.7%) | 247832 (2.1%) | <0.001 | 1.28 (1.26-1.3) |
| | HDL-cholesterol <50 mg/dl | 37975 (5.5%) | 486361 (4.5%) | <0.001 | 1.22 (1.21-1.23) |
| | Cerebrovascular disease | 6715 (0.7%) | 96625 (0.8%) | <0.001 | 0.94 (0.92-0.97) |
| | Transient ischemic attacks | 1850 (0.2%) | 21085 (0.2%) | <0.001 | 1.16 (1.11-1.22) |
| | Cerebral infarction | 2677 (0.3%) | 37571 (0.3%) | 0.01 | 0.95 (0.91-0.99) |
| | Cerebral occlusion without infarction | 2383 (0.3%) | 33350 (0.3%) | 0.017 | 0.95 (0.91-0.99) |
| | Intracerebral hemorrhage | 608 (0.1%) | 8323 (0.1%) | 0.383 | 0.96 (0.89-1.05) |
| | Subarachnoid hemorrhage | 429 (0%) | 5882 (0%) | 0.434 | 0.96 (0.87-1.06) |
| | Hospital admission | 32255 (4.9%) | 457429 (4.6%) | <0.001 | 1.06 (1.05-1.07) |
| | Emergency visits | 43510 (11.4%) | 774690 (10.1%) | <0.001 | 1.13 (1.12-1.14) |
| | Outpatient encounters | 39487 (8.9%) | 722110 (10.2%) | <0.001 | 0.87 (0.86-0.88) |
| | Intensive care unit admission | 1747 (0.2%) | 37950 (0.3%) | <0.001 | 0.62 (0.59-0.65) |
| | Mechanical ventilation | 4219 (0.4%) | 46896 (0.4%) | <0.001 | 1.21 (1.18-1.25) |
| | Referral to Cardio services | 0 (0%) | 56 (0%) | 0.039 | NA |
| | Referral to Neuro services | 56 (0%) | 1714 (0%) | <0.001 | 0.43 (0.33-0.56) |
| | ECMO | 44 (0%) | 486 (0%) | 0.262 | 1.19 (0.88-1.62) |
| | ECMO-VV | 14 (0%) | 159 (0%) | 0.594 | 1.16 (0.67-2) |
| | ECMO-VA | 35 (0%) | 360 (0%) | 0.161 | 1.28 (0.91-1.81) |
| | Mortality | 18316 (1.9%) | 216878 (1.7%) | <0.001 | 1.11 (1.1-1.13) |

For outcomes where event counts were fewer than 10, TriNetX platform suppressed exact numbers and reported '10' for de-identification purposes; thus, relative risks may be overestimated or not statistically meaningful in these cases. When zero events were observed, this is indicated as '0'. Caution is warranted in the interpretation of RRs where numerators are at or below this privacy threshold. ECMO: Extracorporeal membrane oxygenation.

**Table S4.** Outcomes in female infected, unvaccinated individuals compared to naive controls (Group 2 vs Group 1) across four post-exposure time windows.

| Time | Outcome | Group 2 (n=1,200,737) | Group 1 (n=15,366,686) | *p*-value | RR (95%CI) |
|---|---|---|---|---|---|
| | **Follow-up (months)** | **24 ± 16** | **25.1 ± 18** | | |
| 0-3 mo | Major adverse cardiovascular events | 8728 (0.7%) | 104195 (0.7%) | <0.001 | 1.07 (1.05 -1.1) |
| | Cardiovascular disease | 13908 (1.2%) | 147000 (1%) | <0.001 | 1.25 (1.23 -1.27) |
| | Ischemic heart disease | 5353 (0.4%) | 56961 (0.4%) | <0.001 | 1.18 (1.15 -1.21) |
| | Myocardial angina | 517 (0%) | 7045 (0%) | 0.017 | 0.9 (0.82 -0.98) |
| | Acute coronary disease | 962 (0.1%) | 4007 (0%) | <0.001 | 2.93 (2.73 -3.15) |
| | Myocardial infarction | 2377 (0.2%) | 13799 (0.1%) | <0.001 | 2.12 (2.03 -2.21) |
| | Ischemic cardiomyopathy | 401 (0%) | 3218 (0%) | <0.001 | 1.52 (1.37 -1.69) |
| | Arrythmias | 18986 (1.7%) | 160229 (1.1%) | <0.001 | 1.63 (1.6 -1.65) |
| | Atrial fibrillation and flutter | 3540 (0.3%) | 34738 (0.2%) | <0.001 | 1.26 (1.22 -1.3) |
| | Paroxysmal tachycardia | 2226 (0.2%) | 16640 (0.1%) | <0.001 | 1.64 (1.57 -1.72) |
| | Other cardiac arrythmias | 6315 (0.5%) | 47653 (0.3%) | <0.001 | 1.67 (1.62 -1.71) |
| | Inflammatory heart disease | 1916 (0.1%) | 11089 (0.1%) | <0.001 | 2.12 (2.02 -2.22) |
| | Pericarditis | 1362 (0.1%) | 7775 (0%) | <0.001 | 2.14 (2.02 -2.27) |
| | Myocarditis | 199 (0%) | 434 (0%) | <0.001 | 5.59 (4.72 -6.61) |
| | Endocarditis | 511 (0%) | 3321 (0%) | <0.001 | 1.88 (1.71 -2.06) |
| | Hypertensive heart disease | 13577 (1.4%) | 261625 (1.9%) | <0.001 | 0.73 (0.71 -0.74) |
| | Pulmonary heart diseases | 4287 (0.3%) | 22981 (0.1%) | <0.001 | 2.3 (2.23 -2.38) |
| | Heart Failure | 4313 (0.3%) | 35823 (0.2%) | <0.001 | 1.5 (1.45 -1.54) |
| | Cardiomyopathy | 1466 (0.1%) | 12811 (0.1%) | <0.001 | 1.4 (1.33 -1.48) |
| | Cardiomegaly | 4713 (0.4%) | 24776 (0.2%) | <0.001 | 2.35 (2.28 -2.43) |
| | Cardiogenic shock | 474 (0%) | 2453 (0%) | <0.001 | 2.36 (2.13 -2.6) |
| | Cardiac arrest | 1270 (0.1%) | 5670 (0%) | <0.001 | 2.73 (2.57 -2.9) |
| | Thrombotic disorders | 5047 (0.4%) | 25393 (0.2%) | <0.001 | 2.46 (2.39 -2.53) |
| | Pulmonary embolism | 2645 (0.2%) | 10314 (0.1%) | <0.001 | 3.15 (3.01 -3.28) |
| | Deep and superficial thrombosis | 3372 (0.3%) | 18928 (0.1%) | <0.001 | 2.19 (2.12 -2.28) |
| | Thrombosis with Thrombocytopenia | 10 (0%) | 10 (0%) | <0.001 | 12.18 (5.1 -29.2) |
| | Elevated Troponin I | 3806 (0.3%) | 16355 (0.1%) | <0.001 | 2.86 (2.76 -2.96) |
| | Elevated Natriuretic peptide B | 9123 (0.7%) | 40134 (0.3%) | <0.001 | 2.84 (2.78 -2.9) |
| | Dyslipidemia | 22612 (2.7%) | 386766 (2.9%) | <0.001 | 0.92 (0.91 -0.94) |
| | Triglyceride ≥150 mg/dl | 11164 (1%) | 95561 (0.6%) | <0.001 | 1.54 (1.51 -1.57) |
| | Total cholesterol ≥200 mg/dl | 10593 (1%) | 149307 (1%) | 0.019 | 0.98 (0.96 -1) |
| | LDL-cholesterol ≥130 mg/dl | 8378 (0.7%) | 108323 (0.7%) | 0.009 | 1.03 (1.01 -1.05) |
| | HDL-cholesterol <50 mg/dl | 15215 (1.4%) | 170639 (1.1%) | <0.001 | 1.23 (1.21 -1.25) |
| | Cerebrovascular disease | 4353 (0.3%) | 50891 (0.3%) | <0.001 | 1.07 (1.03 -1.1) |
| | Transient ischemic attacks | 890 (0.1%) | 8973 (0.1%) | <0.001 | 1.22 (1.14 -1.3) |
| | Cerebral infarction | 2040 (0.2%) | 20526 (0.1%) | <0.001 | 1.22 (1.17 -1.28) |
| | Cerebral occlusion without infarction | 1357 (0.1%) | 15583 (0.1%) | 0.019 | 1.07 (1.01 -1.13) |
| | Intracerebral hemorrhage | 418 (0%) | 4499 (0%) | 0.015 | 1.13 (1.02 -1.25) |
| | Subarachnoid hemorrhage | 269 (0%) | 3278 (0%) | 0.999 | 1 (0.88 -1.13) |
| | Hospital admission | 27691 (3.2%) | 448931 (3.5%) | <0.001 | 0.91 (0.9 -0.92) |
| | Emergency visits | 23033 (4%) | 340741 (3.1%) | <0.001 | 1.28 (1.27 -1.3) |
| | Outpatient encounters | 25651 (4.2%) | 992889 (9.6%) | <0.001 | 0.44 (0.43 -0.44) |
| | Intensive care unit admission | 1948 (0.2%) | 39889 (0.2%) | <0.001 | 0.61 (0.58 -0.64) |
| | Mechanical ventilation | 7376 (0.6%) | 26248 (0.2%) | <0.001 | 3.45 (3.36 -3.54) |
| | Referral to Cardio services | 10 (0%) | 41 (0%) | 0.001 | 2.97 (1.49 -5.93) |
| | Referral to Neuro services | 33 (0%) | 276 (0%) | 0.04 | 1.46 (1.02 -2.09) |
| | ECMO | 151 (0%) | 313 (0%) | <0.001 | 5.87 (4.84 -7.13) |
| | ECMO-VV | 124 (0%) | 130 (0%) | <0.001 | 11.61 (9.1 -14.8) |
| | ECMO-VA | 38 (0%) | 210 (0%) | <0.001 | 2.2 (1.56 -3.11) |
| | Mortality | 20157 (1.5%) | 60692 (0.4%) | <0.001 | 4.04 (3.98 -4.11) |
| 3-6mo | Major adverse cardiovascular events | 3862 (0.3%) | 40961 (0.3%) | <0.001 | 1.21 (1.17 -1.25) |

| | | | | | |
|---|---|---|---|---|---|
| | Cardiovascular disease | 5506 (0.5%) | 57871 (0.4%) | <0.001 | 1.26 (1.22 -1.29) |
| | Ischemic heart disease | 2347 (0.2%) | 22820 (0.1%) | <0.001 | 1.29 (1.24 -1.35) |
| | Myocardial angina | 346 (0%) | 2863 (0%) | <0.001 | 1.48 (1.32 -1.65) |
| | Acute coronary disease | 366 (0%) | 1965 (0%) | <0.001 | 2.28 (2.04 -2.55) |
| | Myocardial infarction | 850 (0.1%) | 5699 (0%) | <0.001 | 1.83 (1.71 -1.97) |
| | Ischemic cardiomyopathy | 173 (0%) | 1288 (0%) | <0.001 | 1.64 (1.4 -1.92) |
| | Arrythmias | 8580 (0.8%) | 73028 (0.5%) | <0.001 | 1.63 (1.59 -1.66) |
| | Atrial fibrillation and flutter | 1321 (0.1%) | 13319 (0.1%) | <0.001 | 1.23 (1.16 -1.3) |
| | Paroxysmal tachycardia | 1064 (0.1%) | 7579 (0%) | <0.001 | 1.73 (1.62 -1.84) |
| | Other cardiac arrythmias | 2678 (0.2%) | 20441 (0.1%) | <0.001 | 1.65 (1.59 -1.72) |
| | Inflammatory heart disease | 693 (0.1%) | 4448 (0%) | <0.001 | 1.91 (1.76 -2.07) |
| | Pericarditis | 489 (0%) | 3170 (0%) | <0.001 | 1.89 (1.72 -2.08) |
| | Myocarditis | 45 (0%) | 137 (0%) | <0.001 | 4 (2.86 -5.6) |
| | Endocarditis | 216 (0%) | 1310 (0%) | <0.001 | 2.01 (1.74 -2.32) |
| | Hypertensive heart disease | 6769 (0.7%) | 101774 (0.7%) | <0.001 | 0.93 (0.9 -0.95) |
| | Pulmonary heart diseases | 1281 (0.1%) | 9686 (0.1%) | <0.001 | 1.64 (1.54 -1.73) |
| | Heart Failure | 1700 (0.1%) | 14675 (0.1%) | <0.001 | 1.44 (1.37 -1.51) |
| | Cardiomyopathy | 604 (0%) | 5095 (0%) | <0.001 | 1.45 (1.34 -1.58) |
| | Cardiomegaly | 1415 (0.1%) | 9372 (0.1%) | <0.001 | 1.87 (1.77 -1.98) |
| | Cardiogenic shock | 151 (0%) | 938 (0%) | <0.001 | 1.96 (1.65 -2.33) |
| | Cardiac arrest | 426 (0%) | 2464 (0%) | <0.001 | 2.11 (1.9 -2.34) |
| | Thrombotic disorders | 1419 (0.1%) | 10669 (0.1%) | <0.001 | 1.65 (1.56 -1.74) |
| | Pulmonary embolism | 569 (0%) | 4179 (0%) | <0.001 | 1.67 (1.53 -1.83) |
| | Deep and superficial thrombosis | 1144 (0.1%) | 8140 (0.1%) | <0.001 | 1.73 (1.63 -1.84) |
| | Thrombosis with Thrombocytopenia | 0 (0%) | 0 (0%) | NA | NA |
| | Elevated Troponin I | 1212 (0.1%) | 6191 (0%) | <0.001 | 2.41 (2.27 -2.57) |
| | Elevated Natriuretic peptide B | 1869 (0.1%) | 15270 (0.1%) | <0.001 | 1.54 (1.46 -1.61) |
| | Dyslipidemia | 12128 (1.5%) | 142899 (1.1%) | <0.001 | 1.34 (1.31 -1.36) |
| | Triglyceride ≥150 mg/dl | 5276 (0.5%) | 39623 (0.3%) | <0.001 | 1.77 (1.72 -1.82) |
| | Total cholesterol ≥200 mg/dl | 7592 (0.7%) | 58523 (0.4%) | <0.001 | 1.79 (1.74 -1.83) |
| | LDL-cholesterol ≥130 mg/dl | 5889 (0.5%) | 42992 (0.3%) | <0.001 | 1.83 (1.78 -1.88) |
| | HDL-cholesterol <50 mg/dl | 7380 (0.7%) | 61714 (0.4%) | <0.001 | 1.65 (1.62 -1.69) |
| | Cerebrovascular disease | 2165 (0.2%) | 20495 (0.1%) | <0.001 | 1.32 (1.26 -1.38) |
| | Transient ischemic attacks | 552 (0%) | 4244 (0%) | <0.001 | 1.59 (1.46 -1.74) |
| | Cerebral infarction | 884 (0.1%) | 8020 (0.1%) | <0.001 | 1.35 (1.26 -1.45) |
| | Cerebral occlusion without infarction | 737 (0.1%) | 6181 (0%) | <0.001 | 1.46 (1.36 -1.58) |
| | Intracerebral hemorrhage | 202 (0%) | 1367 (0%) | <0.001 | 1.8 (1.55 -2.09) |
| | Subarachnoid hemorrhage | 114 (0%) | 1006 (0%) | 0.001 | 1.38 (1.14 -1.68) |
| | Hospital admission | 11382 (1.3%) | 169673 (1.4%) | 0.049 | 0.98 (0.96 -1) |
| | Emergency visits | 11419 (2.1%) | 176828 (1.7%) | <0.001 | 1.24 (1.22 -1.26) |
| | Outpatient encounters | 12376 (2.1%) | 286297 (3.1%) | <0.001 | 0.69 (0.68 -0.71) |
| | Intensive care unit admission | 790 (0.1%) | 20620 (0.1%) | <0.001 | 0.48 (0.44 -0.51) |
| | Mechanical ventilation | 1090 (0.1%) | 8270 (0.1%) | <0.001 | 1.63 (1.53 -1.73) |
| | Referral to Cardio services | 10 (0%) | 16 (0%) | <0.001 | 7.61 (3.45 -16.7) |
| | Referral to Neuro services | 22 (0%) | 199 (0%) | 0.184 | 1.35 (0.87 -2.09) |
| | ECMO | 10 (0%) | 69 (0%) | 0.089 | 1.77 (0.91 -3.43) |
| | ECMO-VV | 10 (0%) | 28 (0%) | <0.001 | 4.35 (2.11 -8.95) |
| | ECMO-VA | 10 (0%) | 46 (0%) | 0.004 | 2.65 (1.34 -5.25) |
| | Mortality | 4970 (0.4%) | 32324 (0.2%) | <0.001 | 1.87 (1.82 -1.93) |
| 6-9mo | Major adverse cardiovascular events | 3160 (0.3%) | 33069 (0.2%) | <0.001 | 1.22 (1.18 -1.27) |
| | Cardiovascular disease | 4511 (0.4%) | 46206 (0.3%) | <0.001 | 1.29 (1.25 -1.33) |
| | Ischemic heart disease | 1883 (0.2%) | 18338 (0.1%) | <0.001 | 1.29 (1.23 -1.35) |
| | Myocardial angina | 321 (0%) | 2403 (0%) | <0.001 | 1.63 (1.45 -1.84) |
| | Acute coronary disease | 227 (0%) | 1643 (0%) | <0.001 | 1.69 (1.47 -1.94) |
| | Myocardial infarction | 671 (0.1%) | 4942 (0%) | <0.001 | 1.67 (1.54 -1.81) |
| | Ischemic cardiomyopathy | 130 (0%) | 1019 (0%) | <0.001 | 1.56 (1.3 -1.87) |

| | | | | | |
|---|---|---|---|---|---|
| | Arrythmias | 6860 (0.6%) | 60884 (0.4%) | <0.001 | 1.56 (1.52 -1.6) |
| | Atrial fibrillation and flutter | 943 (0.1%) | 10780 (0.1%) | 0.02 | 1.08 (1.01 -1.16) |
| | Paroxysmal tachycardia | 820 (0.1%) | 5977 (0%) | <0.001 | 1.69 (1.57 -1.82) |
| | Other cardiac arrythmias | 2128 (0.2%) | 16542 (0.1%) | <0.001 | 1.62 (1.55 -1.7) |
| | Inflammatory heart disease | 495 (0%) | 3535 (0%) | <0.001 | 1.72 (1.56 -1.89) |
| | Pericarditis | 349 (0%) | 2478 (0%) | <0.001 | 1.72 (1.54 -1.93) |
| | Myocarditis | 26 (0%) | 147 (0%) | <0.001 | 2.16 (1.42 -3.27) |
| | Endocarditis | 149 (0%) | 1035 (0%) | <0.001 | 1.76 (1.48 -2.09) |
| | Hypertensive heart disease | 5599 (0.6%) | 80125 (0.6%) | 0.036 | 0.97 (0.95 -1) |
| | Pulmonary heart diseases | 982 (0.1%) | 7603 (0%) | <0.001 | 1.6 (1.5 -1.71) |
| | Heart Failure | 1215 (0.1%) | 11589 (0.1%) | <0.001 | 1.3 (1.23 -1.38) |
| | Cardiomyopathy | 476 (0%) | 4052 (0%) | <0.001 | 1.44 (1.31 -1.58) |
| | Cardiomegaly | 1196 (0.1%) | 7971 (0.1%) | <0.001 | 1.86 (1.75 -1.98) |
| | Cardiogenic shock | 124 (0%) | 768 (0%) | <0.001 | 1.97 (1.63 -2.38) |
| | Cardiac arrest | 298 (0%) | 1920 (0%) | <0.001 | 1.89 (1.68 -2.14) |
| | Thrombotic disorders | 1139 (0.1%) | 8432 (0.1%) | <0.001 | 1.68 (1.58 -1.78) |
| | Pulmonary embolism | 460 (0%) | 3283 (0%) | <0.001 | 1.72 (1.56 -1.9) |
| | Deep and superficial thrombosis | 873 (0.1%) | 6401 (0%) | <0.001 | 1.68 (1.57 -1.81) |
| | Thrombosis with Thrombocytopenia | 10 (0%) | 10 (0%) | <0.001 | 12.18 (5.1 -29.2) |
| | Elevated Troponin I | 1101 (0.1%) | 5637 (0%) | <0.001 | 2.41 (2.26 -2.57) |
| | Elevated Natriuretic peptide B | 1469 (0.1%) | 12779 (0.1%) | <0.001 | 1.44 (1.37 -1.52) |
| | Dyslipidemia | 10235 (1.3%) | 114886 (0.9%) | <0.001 | 1.41 (1.38 -1.44) |
| | Triglyceride ≥150 mg/dl | 4642 (0.4%) | 34882 (0.2%) | <0.001 | 1.77 (1.71 -1.82) |
| | Total cholesterol ≥200 mg/dl | 6473 (0.6%) | 51271 (0.4%) | <0.001 | 1.74 (1.7 -1.79) |
| | LDL-cholesterol ≥130 mg/dl | 5219 (0.5%) | 38191 (0.3%) | <0.001 | 1.83 (1.77 -1.88) |
| | HDL-cholesterol <50 mg/dl | 6576 (0.6%) | 52154 (0.4%) | <0.001 | 1.75 (1.71 -1.79) |
| | Cerebrovascular disease | 1831 (0.1%) | 16855 (0.1%) | <0.001 | 1.36 (1.29 -1.42) |
| | Transient ischemic attacks | 488 (0%) | 3680 (0%) | <0.001 | 1.63 (1.48 -1.79) |
| | Cerebral infarction | 727 (0.1%) | 6503 (0%) | <0.001 | 1.37 (1.27 -1.48) |
| | Cerebral occlusion without infarction | 591 (0%) | 5074 (0%) | <0.001 | 1.43 (1.31 -1.56) |
| | Intracerebral hemorrhage | 139 (0%) | 1118 (0%) | <0.001 | 1.52 (1.27 -1.81) |
| | Subarachnoid hemorrhage | 105 (0%) | 821 (0%) | <0.001 | 1.56 (1.27 -1.91) |
| | Hospital admission | 8724 (1%) | 142580 (1.2%) | <0.001 | 0.9 (0.88 -0.91) |
| | Emergency visits | 10084 (1.9%) | 151187 (1.4%) | <0.001 | 1.28 (1.26 -1.31) |
| | Outpatient encounters | 9357 (1.6%) | 188386 (2.1%) | <0.001 | 0.79 (0.77 -0.8) |
| | Intensive care unit admission | 674 (0.1%) | 15716 (0.1%) | <0.001 | 0.53 (0.49 -0.57) |
| | Mechanical ventilation | 809 (0.1%) | 6368 (0%) | <0.001 | 1.57 (1.46 -1.69) |
| | Referral to Cardio services | 0 (0%) | 10 (0%) | 0.365 | NA |
| | Referral to Neuro services | 14 (0%) | 207 (0%) | 0.481 | 0.82 (0.48 -1.42) |
| | ECMO | 10 (0%) | 39 (0%) | 0.001 | 3.12 (1.56 -6.26) |
| | ECMO-VV | 10 (0%) | 12 (0%) | <0.001 | 10.15 (4.4 -23.5) |
| | ECMO-VA | 10 (0%) | 25 (0%) | <0.001 | 4.87 (2.34 -10.1) |
| | Mortality | 3680 (0.3%) | 28578 (0.2%) | <0.001 | 1.57 (1.52 -1.62) |
| >9mo | Major adverse cardiovascular events | 15786 (1.3%) | 220036 (1.4%) | <0.001 | 0.92 (0.91 -0.93) |
| | Cardiovascular disease | 22329 (2%) | 301987 (2%) | 0.002 | 0.98 (0.97 -0.99) |
| | Ischemic heart disease | 9303 (0.7%) | 123490 (0.8%) | <0.001 | 0.95 (0.93 -0.97) |
| | Myocardial angina | 1498 (0.1%) | 16055 (0.1%) | <0.001 | 1.14 (1.08 -1.2) |
| | Acute coronary disease | 1158 (0.1%) | 10753 (0.1%) | <0.001 | 1.32 (1.24 -1.4) |
| | Myocardial infarction | 3195 (0.2%) | 34519 (0.2%) | <0.001 | 1.14 (1.1 -1.18) |
| | Ischemic cardiomyopathy | 475 (0%) | 5596 (0%) | 0.468 | 1.04 (0.94 -1.14) |
| | Arrythmias | 39202 (3.7%) | 429856 (2.9%) | <0.001 | 1.27 (1.26 -1.28) |
| | Atrial fibrillation and flutter | 4228 (0.3%) | 65831 (0.4%) | <0.001 | 0.79 (0.77 -0.82) |
| | Paroxysmal tachycardia | 3777 (0.3%) | 40203 (0.3%) | <0.001 | 1.16 (1.12 -1.2) |
| | Other cardiac arrythmias | 10238 (0.8%) | 108121 (0.7%) | <0.001 | 1.19 (1.17 -1.22) |
| | Inflammatory heart disease | 2397 (0.2%) | 23161 (0.1%) | <0.001 | 1.27 (1.22 -1.32) |
| | Pericarditis | 1624 (0.1%) | 15685 (0.1%) | <0.001 | 1.27 (1.2 -1.33) |

| | | | | | |
|---|---|---|---|---|---|
| | Myocarditis | 96 (0%) | 754 (0%) | <0.001 | 1.55 (1.25 -1.92) |
| | Endocarditis | 801 (0.1%) | 7578 (0%) | <0.001 | 1.29 (1.2 -1.39) |
| | Hypertensive heart disease | 35216 (3.6%) | 555908 (4.1%) | <0.001 | 0.88 (0.87 -0.89) |
| | Pulmonary heart diseases | 4176 (0.3%) | 48210 (0.3%) | <0.001 | 1.07 (1.04 -1.11) |
| | Heart Failure | 5316 (0.4%) | 73227 (0.5%) | <0.001 | 0.9 (0.88 -0.93) |
| | Cardiomyopathy | 1868 (0.1%) | 23791 (0.1%) | 0.107 | 0.96 (0.92 -1.01) |
| | Cardiomegaly | 5753 (0.4%) | 56505 (0.4%) | <0.001 | 1.26 (1.23 -1.3) |
| | Cardiogenic shock | 605 (0%) | 5435 (0%) | <0.001 | 1.36 (1.25 -1.48) |
| | Cardiac arrest | 1484 (0.1%) | 14206 (0.1%) | <0.001 | 1.28 (1.21 -1.35) |
| | Thrombotic disorders | 5534 (0.4%) | 58189 (0.4%) | <0.001 | 1.18 (1.15 -1.21) |
| | Pulmonary embolism | 1912 (0.1%) | 20681 (0.1%) | <0.001 | 1.14 (1.08 -1.19) |
| | Deep and superficial thrombosis | 4572 (0.4%) | 45781 (0.3%) | <0.001 | 1.23 (1.2 -1.27) |
| | Thrombosis with Thrombocytopenia | 10 (0%) | 12 (0%) | <0.001 | 10.15 (4.4 -23.4) |
| | Elevated Troponin I | 7036 (0.5%) | 55166 (0.3%) | <0.001 | 1.57 (1.53 -1.61) |
| | Elevated Natriuretic peptide B | 7707 (0.6%) | 94741 (0.6%) | 0.072 | 1.02 (1 -1.05) |
| | Dyslipidemia | 66670 (8.4%) | 919473 (7.3%) | <0.001 | 1.15 (1.14 -1.16) |
| | Triglyceride ≥150 mg/dl | 32118 (2.8%) | 306738 (2%) | <0.001 | 1.39 (1.38 -1.41) |
| | Total cholesterol ≥200 mg/dl | 44690 (4.2%) | 475316 (3.3%) | <0.001 | 1.3 (1.29 -1.31) |
| | LDL-cholesterol ≥130 mg/dl | 36524 (3.3%) | 361503 (2.4%) | <0.001 | 1.35 (1.34 -1.37) |
| | HDL-cholesterol <50 mg/dl | 43758 (4.2%) | 454929 (3.1%) | <0.001 | 1.34 (1.33 -1.35) |
| | Cerebrovascular disease | 9151 (0.7%) | 116412 (0.7%) | 0.077 | 0.98 (0.96 -1) |
| | Transient ischemic attacks | 2861 (0.2%) | 29378 (0.2%) | <0.001 | 1.19 (1.15 -1.24) |
| | Cerebral infarction | 3409 (0.3%) | 42517 (0.3%) | 0.414 | 0.99 (0.95 -1.02) |
| | Cerebral occlusion without infarction | 2872 (0.2%) | 35135 (0.2%) | 0.863 | 1 (0.97 -1.04) |
| | Intracerebral hemorrhage | 650 (0%) | 7934 (0%) | 0.973 | 1 (0.92 -1.08) |
| | Subarachnoid hemorrhage | 485 (0%) | 6129 (0%) | 0.44 | 0.96 (0.88 -1.06) |
| | Hospital admission | 48472 (5.9%) | 650596 (5.4%) | <0.001 | 1.09 (1.08 -1.1) |
| | Emergency visits | 59536 (11.2%) | 998522 (9.7%) | <0.001 | 1.15 (1.14 -1.16) |
| | Outpatient encounters | 51349 (9.2%) | 957650 (10.8%) | <0.001 | 0.85 (0.84 -0.85) |
| | Intensive care unit admission | 2609 (0.2%) | 50492 (0.3%) | <0.001 | 0.64 (0.62 -0.67) |
| | Mechanical ventilation | 4045 (0.3%) | 42625 (0.3%) | <0.001 | 1.17 (1.13 -1.21) |
| | Referral to Cardio services | 10 (0%) | 56 (0%) | 0.02 | 2.17 (1.11 -4.26) |
| | Referral to Neuro services | 68 (0%) | 2534 (0%) | <0.001 | 0.33 (0.26 -0.42) |
| | ECMO | 28 (0%) | 304 (0%) | 0.561 | 1.12 (0.76 -1.65) |
| | ECMO-VV | 12 (0%) | 100 (0%) | 0.212 | 1.46 (0.8 -2.66) |
| | ECMO-VA | 19 (0%) | 218 (0%) | 0.804 | 1.06 (0.66 -1.7) |
| | Mortality | 18156 (1.4%) | 198881 (1.2%) | <0.001 | 1.11 (1.1 -1.13) |

For outcomes where event counts were fewer than 10, TriNetX platform suppressed exact numbers and reported '10' for de-identification purposes; thus, relative risks may be overestimated or not statistically meaningful in these cases. When zero events were observed, this is indicated as '0'. Caution is warranted in the interpretation of RRs where numerators are at or below this privacy threshold. ECMO: Extracorporeal membrane oxygenation.

**Table S5.** Outcomes in male in vaccinated individuals without prior infection compared to naive controls (Group 3 vs Group 1) across four post-exposure time windows.

| Time | Outcome | Group 3 (n=613,395) | Group 1 (n=11,250,469) | *p*-value | RR (95%CI) |
|---|---|---|---|---|---|
| | **Follow-up (months)** | **22.9 ± 19.3** | **22.4 ± 17.5** | | |
| 0-3 mo | Major adverse cardiovascular events | 1753 (0.3%) | 134603 (1.2%) | <0.001 | 0.24 (0.23 -0.25) |
| | Cardiovascular disease | 2258 (0.4%) | 171975 (1.6%) | <0.001 | 0.24 (0.23 -0.25) |
| | Ischemic heart disease | 1249 (0.2%) | 89703 (0.8%) | <0.001 | 0.26 (0.24 -0.27) |
| | Myocardial angina | 194 (0%) | 9660 (0.1%) | <0.001 | 0.37 (0.32 -0.43) |
| | Acute coronary disease | 127 (0%) | 5209 (0%) | <0.001 | 0.45 (0.38 -0.53) |
| | Myocardial infarction | 337 (0.1%) | 20489 (0.2%) | <0.001 | 0.3 (0.27 -0.34) |
| | Ischemic cardiomyopathy | 166 (0%) | 9354 (0.1%) | <0.001 | 0.33 (0.28 -0.38) |
| | Arrythmias | 2521 (0.4%) | 151372 (1.4%) | <0.001 | 0.31 (0.3 -0.32) |
| | Atrial fibrillation and flutter | 686 (0.1%) | 48900 (0.4%) | <0.001 | 0.26 (0.24 -0.28) |
| | Paroxysmal tachycardia | 415 (0.1%) | 17571 (0.1%) | <0.001 | 0.43 (0.39 -0.48) |
| | Other cardiac arrythmias | 992 (0.2%) | 50267 (0.4%) | <0.001 | 0.37 (0.34 -0.39) |
| | Inflammatory heart disease | 248 (0%) | 12113 (0.1%) | <0.001 | 0.38 (0.33 -0.43) |
| | Pericarditis | 166 (0%) | 7811 (0.1%) | <0.001 | 0.39 (0.33 -0.45) |
| | Myocarditis | 13 (0%) | 778 (0%) | <0.001 | 0.31 (0.18 -0.53) |
| | Endocarditis | 81 (0%) | 4208 (0%) | <0.001 | 0.35 (0.28 -0.44) |
| | Hypertensive heart disease | 3293 (0.6%) | 276893 (2.7%) | <0.001 | 0.23 (0.22 -0.24) |
| | Pulmonary heart diseases | 444 (0.1%) | 21963 (0.2%) | <0.001 | 0.37 (0.34 -0.41) |
| | Heart Failure | 677 (0.1%) | 44174 (0.4%) | <0.001 | 0.28 (0.26 -0.3) |
| | Cardiomyopathy | 371 (0.1%) | 23244 (0.2%) | <0.001 | 0.29 (0.26 -0.33) |
| | Cardiomegaly | 568 (0.1%) | 30736 (0.3%) | <0.001 | 0.34 (0.31 -0.37) |
| | Cardiogenic shock | 72 (0%) | 4281 (0%) | <0.001 | 0.31 (0.24 -0.39) |
| | Cardiac arrest | 165 (0%) | 9148 (0.1%) | <0.001 | 0.33 (0.28 -0.39) |
| | Thrombotic disorders | 409 (0.1%) | 26073 (0.2%) | <0.001 | 0.29 (0.26 -0.32) |
| | Pulmonary embolism | 169 (0%) | 10153 (0.1%) | <0.001 | 0.31 (0.26 -0.36) |
| | Deep and superficial thrombosis | 325 (0%) | 20155 (0.2%) | <0.001 | 0.3 (0.27 -0.33) |
| | Thrombosis with Thrombocytopenia | 0 (0%) | 10 (0%) | 0.46 | NA |
| | Elevated Troponin I | 266 (0%) | 23229 (0.2%) | <0.001 | 0.21 (0.19 -0.24) |
| | Elevated Natriuretic peptide B | 785 (0.1%) | 43562 (0.4%) | <0.001 | 0.33 (0.31 -0.36) |
| | Dyslipidemia | 6045 (1.3%) | 414525 (4.3%) | <0.001 | 0.31 (0.3 -0.31) |
| | Triglyceride ≥150 mg/dl | 2937 (0.5%) | 129324 (1.1%) | <0.001 | 0.44 (0.43 -0.46) |
| | Total cholesterol ≥200 mg/dl | 2791 (0.5%) | 124493 (1.1%) | <0.001 | 0.44 (0.42 -0.45) |
| | LDL-cholesterol ≥130 mg/dl | 2426 (0.4%) | 107147 (0.9%) | <0.001 | 0.44 (0.42 -0.45) |
| | HDL-cholesterol <50 mg/dl | 4947 (0.9%) | 269080 (2.5%) | <0.001 | 0.37 (0.36 -0.38) |
| | Cerebrovascular disease | 827 (0.1%) | 51839 (0.4%) | <0.001 | 0.29 (0.27 -0.31) |
| | Transient ischemic attacks | 178 (0%) | 7631 (0.1%) | <0.001 | 0.43 (0.37 -0.5) |
| | Cerebral infarction | 331 (0.1%) | 20920 (0.2%) | <0.001 | 0.29 (0.26 -0.32) |
| | Cerebral occlusion without infarction | 298 (0%) | 18508 (0.2%) | <0.001 | 0.3 (0.26 -0.33) |
| | Intracerebral hemorrhage | 85 (0%) | 5758 (0%) | <0.001 | 0.27 (0.22 -0.34) |
| | Subarachnoid hemorrhage | 43 (0%) | 3969 (0%) | <0.001 | 0.2 (0.15 -0.27) |
| | Hospital admission | 2486 (0.4%) | 362735 (3.6%) | <0.001 | 0.12 (0.12 -0.13) |
| | Emergency visits | 5103 (1.1%) | 276792 (3.5%) | <0.001 | 0.31 (0.3 -0.32) |
| | Outpatient encounters | 7662 (2%) | 861852 (11.2%) | <0.001 | 0.18 (0.18 -0.18) |
| | Intensive care unit admission | 142 (0%) | 37166 (0.3%) | <0.001 | 0.07 (0.06 -0.08) |
| | Mechanical ventilation | 356 (0.1%) | 36596 (0.3%) | <0.001 | 0.18 (0.16 -0.2) |
| | Referral to Cardio services | 0 (0%) | 22 (0%) | 0.273 | NA |
| | Referral to Neuro services | 10 (0%) | 247 (0%) | 0.353 | 0.74 (0.39 -1.4) |
| | ECMO | 10 (0%) | 621 (0%) | <0.001 | 0.3 (0.16 -0.55) |
| | ECMO-VV | 10 (0%) | 227 (0%) | 0.506 | 0.81 (0.43 -1.52) |
| | ECMO-VA | 10 (0%) | 435 (0%) | 0.005 | 0.42 (0.23 -0.79) |
| | Mortality | 1349 (0.2%) | 72284 (0.6%) | <0.001 | 0.34 (0.32 -0.36) |
| 3-6mo | Major adverse cardiovascular events | 1467 (0.2%) | 45287 (0.4%) | <0.001 | 0.59 (0.56 -0.62) |

| | | | | | |
|---|---|---|---|---|---|
| | Cardiovascular disease | 1811 (0.3%) | 57141 (0.5%) | <0.001 | 0.58 (0.55 -0.6) |
| | Ischemic heart disease | 1024 (0.2%) | 30230 (0.3%) | <0.001 | 0.62 (0.58 -0.66) |
| | Myocardial angina | 156 (0%) | 3280 (0%) | 0.097 | 0.87 (0.74 -1.03) |
| | Acute coronary disease | 120 (0%) | 2173 (0%) | 0.9 | 1.01 (0.84 -1.22) |
| | Myocardial infarction | 308 (0%) | 7077 (0.1%) | <0.001 | 0.8 (0.71 -0.89) |
| | Ischemic cardiomyopathy | 126 (0%) | 3307 (0%) | <0.001 | 0.7 (0.59 -0.83) |
| | Arrythmias | 2133 (0.4%) | 56594 (0.5%) | <0.001 | 0.69 (0.66 -0.72) |
| | Atrial fibrillation and flutter | 562 (0.1%) | 16987 (0.1%) | <0.001 | 0.6 (0.56 -0.66) |
| | Paroxysmal tachycardia | 347 (0.1%) | 6750 (0.1%) | 0.288 | 0.94 (0.85 -1.05) |
| | Other cardiac arrythmias | 821 (0.1%) | 18096 (0.2%) | <0.001 | 0.84 (0.78 -0.9) |
| | Inflammatory heart disease | 215 (0%) | 4204 (0%) | 0.355 | 0.94 (0.82 -1.08) |
| | Pericarditis | 137 (0%) | 2763 (0%) | 0.273 | 0.91 (0.77 -1.08) |
| | Myocarditis | 10 (0%) | 204 (0%) | 0.74 | 0.9 (0.48 -1.69) |
| | Endocarditis | 78 (0%) | 1453 (0%) | 0.889 | 0.98 (0.78 -1.24) |
| | Hypertensive heart disease | 2478 (0.5%) | 93614 (0.9%) | <0.001 | 0.5 (0.48 -0.52) |
| | Pulmonary heart diseases | 405 (0.1%) | 8295 (0.1%) | 0.03 | 0.9 (0.81 -0.99) |
| | Heart Failure | 571 (0.1%) | 16032 (0.1%) | <0.001 | 0.65 (0.6 -0.71) |
| | Cardiomyopathy | 305 (0%) | 8091 (0.1%) | <0.001 | 0.69 (0.62 -0.78) |
| | Cardiomegaly | 430 (0.1%) | 9442 (0.1%) | <0.001 | 0.84 (0.76 -0.92) |
| | Cardiogenic shock | 81 (0%) | 1434 (0%) | 0.766 | 1.04 (0.83 -1.29) |
| | Cardiac arrest | 149 (0%) | 3372 (0%) | 0.011 | 0.81 (0.69 -0.95) |
| | Thrombotic disorders | 370 (0.1%) | 9739 (0.1%) | <0.001 | 0.7 (0.63 -0.77) |
| | Pulmonary embolism | 156 (0%) | 3756 (0%) | 0.001 | 0.76 (0.65 -0.89) |
| | Deep and superficial thrombosis | 277 (0%) | 7489 (0.1%) | <0.001 | 0.68 (0.6 -0.76) |
| | Thrombosis with Thrombocytopenia | 0 (0%) | 10 (0%) | 0.46 | NA |
| | Elevated Troponin I | 274 (0%) | 7357 (0.1%) | <0.001 | 0.68 (0.6 -0.77) |
| | Elevated Natriuretic peptide B | 704 (0.1%) | 14607 (0.1%) | 0.001 | 0.88 (0.82 -0.95) |
| | Dyslipidemia | 4201 (0.9%) | 122429 (1.3%) | <0.001 | 0.7 (0.68 -0.72) |
| | Triglyceride ≥150 mg/dl | 2137 (0.4%) | 40756 (0.4%) | 0.387 | 1.02 (0.98 -1.07) |
| | Total cholesterol ≥200 mg/dl | 1961 (0.3%) | 36956 (0.3%) | 0.257 | 1.03 (0.98 -1.07) |
| | LDL-cholesterol ≥130 mg/dl | 1705 (0.3%) | 31328 (0.3%) | 0.087 | 1.04 (0.99 -1.1) |
| | HDL-cholesterol <50 mg/dl | 3372 (0.6%) | 75695 (0.7%) | <0.001 | 0.89 (0.86 -0.92) |
| | Cerebrovascular disease | 691 (0.1%) | 18177 (0.2%) | <0.001 | 0.7 (0.64 -0.75) |
| | Transient ischemic attacks | 163 (0%) | 3062 (0%) | 0.77 | 0.98 (0.83 -1.14) |
| | Cerebral infarction | 278 (0%) | 7472 (0.1%) | <0.001 | 0.68 (0.6 -0.77) |
| | Cerebral occlusion without infarction | 247 (0%) | 6340 (0.1%) | <0.001 | 0.71 (0.63 -0.81) |
| | Intracerebral hemorrhage | 68 (0%) | 1547 (0%) | 0.079 | 0.81 (0.63 -1.03) |
| | Subarachnoid hemorrhage | 51 (0%) | 1005 (0%) | 0.609 | 0.93 (0.7 -1.23) |
| | Hospital admission | 2051 (0.4%) | 99835 (1%) | <0.001 | 0.35 (0.34 -0.37) |
| | Emergency visits | 4893 (1.1%) | 132112 (1.7%) | <0.001 | 0.61 (0.59 -0.63) |
| | Outpatient encounters | 5810 (1.6%) | 210242 (3.1%) | <0.001 | 0.51 (0.49 -0.52) |
| | Intensive care unit admission | 81 (0%) | 14309 (0.1%) | <0.001 | 0.1 (0.08 -0.13) |
| | Mechanical ventilation | 309 (0%) | 9565 (0.1%) | <0.001 | 0.59 (0.53 -0.66) |
| | Referral to Cardio services | 0 (0%) | 10 (0%) | 0.46 | NA |
| | Referral to Neuro services | 10 (0%) | 155 (0%) | 0.608 | 1.18 (0.62 -2.24) |
| | ECMO | 10 (0%) | 115 (0%) | 0.154 | 1.59 (0.84 -3.04) |
| | ECMO-VV | 10 (0%) | 38 (0%) | <0.001 | 4.82 (2.4 -9.68) |
| | ECMO-VA | 10 (0%) | 86 (0%) | 0.02 | 2.13 (1.11 -4.1) |
| | Mortality | 1523 (0.2%) | 36742 (0.3%) | <0.001 | 0.76 (0.72 -0.8) |
| 6-9mo | Major adverse cardiovascular events | 1348 (0.2%) | 34489 (0.3%) | <0.001 | 0.71 (0.67 -0.75) |
| | Cardiovascular disease | 1669 (0.3%) | 43075 (0.4%) | <0.001 | 0.7 (0.67 -0.74) |
| | Ischemic heart disease | 948 (0.2%) | 22875 (0.2%) | <0.001 | 0.76 (0.71 -0.81) |
| | Myocardial angina | 160 (0%) | 2591 (0%) | 0.124 | 1.13 (0.97 -1.33) |
| | Acute coronary disease | 115 (0%) | 1792 (0%) | 0.092 | 1.18 (0.97 -1.42) |
| | Myocardial infarction | 313 (0%) | 6028 (0.1%) | 0.376 | 0.95 (0.85 -1.06) |
| | Ischemic cardiomyopathy | 132 (0%) | 2275 (0%) | 0.492 | 1.06 (0.89 -1.27) |

| | | | | | |
|---|---|---|---|---|---|
| | Arrythmias | 1947 (0.3%) | 44652 (0.4%) | <0.001 | 0.8 (0.76 -0.84) |
| | Atrial fibrillation and flutter | 576 (0.1%) | 12806 (0.1%) | <0.001 | 0.82 (0.76 -0.89) |
| | Paroxysmal tachycardia | 300 (0%) | 5001 (0%) | 0.107 | 1.1 (0.98 -1.24) |
| | Other cardiac arrythmias | 747 (0.1%) | 13892 (0.1%) | 0.811 | 0.99 (0.92 -1.07) |
| | Inflammatory heart disease | 171 (0%) | 3295 (0%) | 0.523 | 0.95 (0.82 -1.11) |
| | Pericarditis | 124 (0%) | 2152 (0%) | 0.556 | 1.06 (0.88 -1.27) |
| | Myocarditis | 10 (0%) | 179 (0%) | 0.943 | 1.02 (0.54 -1.94) |
| | Endocarditis | 50 (0%) | 1139 (0%) | 0.132 | 0.81 (0.61 -1.07) |
| | Hypertensive heart disease | 2302 (0.4%) | 68707 (0.7%) | <0.001 | 0.62 (0.6 -0.65) |
| | Pulmonary heart diseases | 357 (0.1%) | 6207 (0.1%) | 0.324 | 1.06 (0.95 -1.17) |
| | Heart Failure | 548 (0.1%) | 11792 (0.1%) | <0.001 | 0.85 (0.78 -0.93) |
| | Cardiomyopathy | 303 (0%) | 5804 (0%) | 0.452 | 0.96 (0.85 -1.07) |
| | Cardiomegaly | 404 (0.1%) | 7477 (0.1%) | 0.881 | 0.99 (0.9 -1.1) |
| | Cardiogenic shock | 94 (0%) | 1138 (0%) | <0.001 | 1.51 (1.23 -1.87) |
| | Cardiac arrest | 176 (0%) | 2819 (0%) | 0.086 | 1.14 (0.98 -1.33) |
| | Thrombotic disorders | 361 (0.1%) | 7428 (0.1%) | 0.031 | 0.89 (0.8 -0.99) |
| | Pulmonary embolism | 135 (0%) | 2902 (0%) | 0.07 | 0.85 (0.72 -1.01) |
| | Deep and superficial thrombosis | 283 (0%) | 5765 (0%) | 0.082 | 0.9 (0.8 -1.01) |
| | Thrombosis with Thrombocytopenia | 0 (0%) | 0 (0%) | NA | NA |
| | Elevated Troponin I | 260 (0%) | 6438 (0.1%) | <0.001 | 0.74 (0.65 -0.84) |
| | Elevated Natriuretic peptide B | 629 (0.1%) | 11549 (0.1%) | 0.941 | 1 (0.92 -1.08) |
| | Dyslipidemia | 3596 (0.8%) | 92080 (1%) | <0.001 | 0.79 (0.76 -0.82) |
| | Triglyceride ≥150 mg/dl | 1938 (0.3%) | 34156 (0.3%) | <0.001 | 1.1 (1.05 -1.16) |
| | Total cholesterol ≥200 mg/dl | 1860 (0.3%) | 30904 (0.3%) | <0.001 | 1.16 (1.11 -1.22) |
| | LDL-cholesterol ≥130 mg/dl | 1721 (0.3%) | 26624 (0.2%) | <0.001 | 1.24 (1.18 -1.3) |
| | HDL-cholesterol <50 mg/dl | 2922 (0.6%) | 60684 (0.6%) | 0.042 | 0.96 (0.93 -1) |
| | Cerebrovascular disease | 656 (0.1%) | 14239 (0.1%) | <0.001 | 0.84 (0.78 -0.91) |
| | Transient ischemic attacks | 171 (0%) | 2603 (0%) | 0.018 | 1.21 (1.03 -1.41) |
| | Cerebral infarction | 304 (0%) | 5745 (0%) | 0.582 | 0.97 (0.86 -1.09) |
| | Cerebral occlusion without infarction | 252 (0%) | 5058 (0%) | 0.161 | 0.91 (0.81 -1.04) |
| | Intracerebral hemorrhage | 77 (0%) | 1181 (0%) | 0.132 | 1.19 (0.95 -1.5) |
| | Subarachnoid hemorrhage | 39 (0%) | 857 (0%) | 0.265 | 0.83 (0.61 -1.15) |
| | Hospital admission | 1915 (0.3%) | 66439 (0.7%) | <0.001 | 0.49 (0.47 -0.51) |
| | Emergency visits | 4375 (1%) | 109837 (1.5%) | <0.001 | 0.65 (0.63 -0.67) |
| | Outpatient encounters | 4890 (1.3%) | 132293 (2%) | <0.001 | 0.67 (0.65 -0.69) |
| | Intensive care unit admission | 70 (0%) | 10503 (0.1%) | <0.001 | 0.12 (0.1 -0.15) |
| | Mechanical ventilation | 329 (0.1%) | 7163 (0.1%) | 0.002 | 0.84 (0.75 -0.94) |
| | Referral to Cardio services | 0 (0%) | 10 (0%) | 0.46 | NA |
| | Referral to Neuro services | 10 (0%) | 143 (0%) | 0.447 | 1.28 (0.68 -2.43) |
| | ECMO | 10 (0%) | 94 (0%) | 0.041 | 1.95 (1.02 -3.74) |
| | ECMO-VV | 10 (0%) | 34 (0%) | <0.001 | 5.39 (2.66 -10.91) |
| | ECMO-VA | 10 (0%) | 60 (0%) | 0.001 | 3.05 (1.56 -5.96) |
| | Mortality | 1631 (0.2%) | 31950 (0.3%) | 0.008 | 0.94 (0.89 -0.98) |
| >9mo | Major adverse cardiovascular events | 10387 (1.7%) | 202392 (1.8%) | <0.001 | 0.93 (0.91 -0.95) |
| | Cardiovascular disease | 13706 (2.3%) | 253843 (2.4%) | 0.014 | 0.98 (0.96 -1) |
| | Ischemic heart disease | 7082 (1.1%) | 131991 (1.2%) | 0.074 | 0.98 (0.96 -1) |
| | Myocardial angina | 964 (0.1%) | 15932 (0.1%) | 0.002 | 1.11 (1.04 -1.19) |
| | Acute coronary disease | 787 (0.1%) | 10297 (0.1%) | <0.001 | 1.4 (1.3 -1.51) |
| | Myocardial infarction | 2173 (0.3%) | 35788 (0.3%) | <0.001 | 1.11 (1.06 -1.16) |
| | Ischemic cardiomyopathy | 766 (0.1%) | 12340 (0.1%) | 0.001 | 1.14 (1.06 -1.22) |
| | Arrythmias | 15712 (2.6%) | 279594 (2.6%) | 0.001 | 1.03 (1.01 -1.04) |
| | Atrial fibrillation and flutter | 3952 (0.6%) | 73101 (0.6%) | 0.397 | 0.99 (0.96 -1.02) |
| | Paroxysmal tachycardia | 2249 (0.3%) | 29493 (0.2%) | <0.001 | 1.4 (1.34 -1.46) |
| | Other cardiac arrythmias | 5717 (0.9%) | 85656 (0.7%) | <0.001 | 1.23 (1.2 -1.26) |
| | Inflammatory heart disease | 1244 (0.2%) | 18565 (0.2%) | <0.001 | 1.23 (1.16 -1.3) |

| | | | | | |
|---|---|---|---|---|---|
| | Pericarditis | 783 (0.1%) | 11604 (0.1%) | <0.001 | 1.24 (1.15 -1.33) |
| | Myocarditis | 39 (0%) | 927 (0%) | 0.11 | 0.77 (0.56 -1.06) |
| | Endocarditis | 488 (0.1%) | 6990 (0.1%) | <0.001 | 1.28 (1.17 -1.4) |
| | Hypertensive heart disease | 22029 (4.2%) | 431149 (4.4%) | <0.001 | 0.95 (0.94 -0.96) |
| | Pulmonary heart diseases | 2467 (0.4%) | 35833 (0.3%) | <0.001 | 1.26 (1.21 -1.32) |
| | Heart Failure | 3908 (0.6%) | 69294 (0.6%) | 0.072 | 1.03 (1 -1.06) |
| | Cardiomyopathy | 1917 (0.3%) | 31567 (0.3%) | <0.001 | 1.11 (1.06 -1.17) |
| | Cardiomegaly | 2757 (0.4%) | 47843 (0.4%) | 0.004 | 1.06 (1.02 -1.1) |
| | Cardiogenic shock | 570 (0.1%) | 7069 (0.1%) | <0.001 | 1.48 (1.36 -1.61) |
| | Cardiac arrest | 1291 (0.2%) | 18904 (0.2%) | <0.001 | 1.25 (1.18 -1.32) |
| | Thrombotic disorders | 2669 (0.4%) | 44998 (0.4%) | <0.001 | 1.09 (1.05 -1.13) |
| | Pulmonary embolism | 1018 (0.2%) | 16069 (0.1%) | <0.001 | 1.16 (1.09 -1.24) |
| | Deep and superficial thrombosis | 2139 (0.3%) | 35893 (0.3%) | <0.001 | 1.09 (1.05 -1.14) |
| | Thrombosis with Thrombocytopenia | 10 (0%) | 11 (0%) | <0.001 | 16.66 (7.07 - 39.22) |
| | Elevated Troponin I | 3128 (0.5%) | 54866 (0.5%) | 0.028 | 1.04 (1 -1.08) |
| | Elevated Natriuretic peptide B | 4574 (0.7%) | 76989 (0.7%) | <0.001 | 1.09 (1.06 -1.12) |
| | Dyslipidemia | 34094 (7.6%) | 633544 (7%) | <0.001 | 1.09 (1.07 -1.1) |
| | Triglyceride ≥150 mg/dl | 18281 (3.2%) | 270247 (2.4%) | <0.001 | 1.32 (1.3 -1.34) |
| | Total cholesterol ≥200 mg/dl | 17805 (3.1%) | 264757 (2.4%) | <0.001 | 1.3 (1.28 -1.32) |
| | LDL-cholesterol ≥130 mg/dl | 16079 (2.7%) | 231973 (2.1%) | <0.001 | 1.33 (1.31 -1.35) |
| | HDL-cholesterol <50 mg/dl | 26432 (5.2%) | 451315 (4.4%) | <0.001 | 1.17 (1.16 -1.18) |
| | Cerebrovascular disease | 4844 (0.8%) | 88233 (0.8%) | 0.827 | 1 (0.98 -1.03) |
| | Transient ischemic attacks | 1233 (0.2%) | 18935 (0.2%) | <0.001 | 1.2 (1.13 -1.27) |
| | Cerebral infarction | 1884 (0.3%) | 33793 (0.3%) | 0.402 | 1.02 (0.97 -1.07) |
| | Cerebral occlusion without infarction | 1687 (0.3%) | 30475 (0.3%) | 0.554 | 1.02 (0.97 -1.07) |
| | Intracerebral hemorrhage | 472 (0.1%) | 7572 (0.1%) | 0.005 | 1.14 (1.04 -1.25) |
| | Subarachnoid hemorrhage | 360 (0.1%) | 5313 (0%) | <0.001 | 1.24 (1.12 -1.38) |
| | Hospital admission | 16714 (3%) | 423689 (4.4%) | <0.001 | 0.66 (0.65 -0.68) |
| | Emergency visits | 38600 (8.5%) | 691031 (9.4%) | <0.001 | 0.91 (0.9 -0.91) |
| | Outpatient encounters | 35855 (10%) | 626610 (9.7%) | <0.001 | 1.03 (1.02 -1.04) |
| | Intensive care unit admission | 319 (0%) | 37811 (0.3%) | <0.001 | 0.15 (0.14 -0.17) |
| | Mechanical ventilation | 2498 (0.4%) | 44042 (0.4%) | 0.107 | 1.03 (0.99 -1.08) |
| | Referral to Cardio services | 0 (0%) | 59 (0%) | 0.073 | NA |
| | Referral to Neuro services | 25 (0%) | 1685 (0%) | <0.001 | 0.27 (0.18 -0.4) |
| | ECMO | 19 (0%) | 470 (0%) | 0.198 | 0.74 (0.47 -1.17) |
| | ECMO-VV | 10 (0%) | 140 (0%) | 0.41 | 1.31 (0.69 -2.49) |
| | ECMO-VA | 16 (0%) | 346 (0%) | 0.516 | 0.85 (0.51 -1.4) |
| | Mortality | 13876 (2.1%) | 207665 (1.7%) | <0.001 | 1.22 (1.2 -1.25) |

For outcomes where event counts were fewer than 10, TriNetX platform suppressed exact numbers and reported '10' for de-identification purposes; thus, relative risks may be overestimated or not statistically meaningful in these cases. When zero events were observed, this is indicated as '0'. Caution is warranted in the interpretation of RRs where numerators are at or below this privacy threshold. ECMO: Extracorporeal membrane oxygenation.

**Table S6.** Outcomes in female vaccinated individuals without prior infection compared to naive controls (Group 3 vs Group 1) across four post-exposure time windows.

| Time | Outcome | Group 3 (n=742,409) | Group 1 (n=15,528,569 ) | *p*-value | RR (95%CI) |
|---|---|---|---|---|---|
| | **Follow-up (months)** | **26.5 ± 18.9** | **25.1 ± 18** | | |
| 0-3 mo | Major adverse cardiovascular events | 1775 (0.2%) | 106280 (0.7%) | <0.001 | 0.35 (0.33 -0.37) |
| | Cardiovascular disease | 2452 (0.3%) | 149709 (1%) | <0.001 | 0.35 (0.33 -0.36) |
| | Ischemic heart disease | 1065 (0.1%) | 58318 (0.4%) | <0.001 | 0.38 (0.36 -0.4) |
| | Myocardial angina | 165 (0%) | 7134 (0%) | <0.001 | 0.48 (0.41 -0.56) |
| | Acute coronary disease | 125 (0%) | 4036 (0%) | <0.001 | 0.64 (0.54 -0.77) |
| | Myocardial infarction | 305 (0%) | 13959 (0.1%) | <0.001 | 0.45 (0.4 -0.51) |
| | Ischemic cardiomyopathy | 69 (0%) | 3292 (0%) | <0.001 | 0.43 (0.34 -0.55) |
| | Arrythmias | 3405 (0.5%) | 162389 (1.1%) | <0.001 | 0.45 (0.43 -0.46) |
| | Atrial fibrillation and flutter | 526 (0.1%) | 35500 (0.2%) | <0.001 | 0.31 (0.28 -0.34) |
| | Paroxysmal tachycardia | 477 (0.1%) | 16926 (0.1%) | <0.001 | 0.59 (0.53 -0.64) |
| | Other cardiac arrythmias | 1067 (0.1%) | 48228 (0.3%) | <0.001 | 0.46 (0.44 -0.49) |
| | Inflammatory heart disease | 248 (0%) | 11138 (0.1%) | <0.001 | 0.46 (0.41 -0.52) |
| | Pericarditis | 181 (0%) | 7791 (0%) | <0.001 | 0.48 (0.42 -0.56) |
| | Myocarditis | 10 (0%) | 439 (0%) | 0.016 | 0.47 (0.25 -0.88) |
| | Endocarditis | 74 (0%) | 3359 (0%) | <0.001 | 0.46 (0.36 -0.58) |
| | Hypertensive heart disease | 3422 (0.5%) | 267918 (1.9%) | <0.001 | 0.28 (0.27 -0.29) |
| | Pulmonary heart diseases | 530 (0.1%) | 23395 (0.1%) | <0.001 | 0.47 (0.43 -0.51) |
| | Heart Failure | 654 (0.1%) | 36449 (0.2%) | <0.001 | 0.37 (0.35 -0.4) |
| | Cardiomyopathy | 265 (0%) | 13112 (0.1%) | <0.001 | 0.42 (0.37 -0.47) |
| | Cardiomegaly | 529 (0.1%) | 24586 (0.2%) | <0.001 | 0.45 (0.41 -0.49) |
| | Cardiogenic shock | 57 (0%) | 2505 (0%) | <0.001 | 0.47 (0.36 -0.61) |
| | Cardiac arrest | 105 (0%) | 5801 (0%) | <0.001 | 0.38 (0.31 -0.46) |
| | Thrombotic disorders | 510 (0.1%) | 25713 (0.2%) | <0.001 | 0.41 (0.38 -0.45) |
| | Pulmonary embolism | 216 (0%) | 10428 (0.1%) | <0.001 | 0.43 (0.38 -0.49) |
| | Deep and superficial thrombosis | 391 (0.1%) | 19195 (0.1%) | <0.001 | 0.42 (0.38 -0.47) |
| | Thrombosis with Thrombocytopenia | 0 (0%) | 10 (0%) | 0.487 | NA |
| | Elevated Troponin I | 265 (0%) | 16797 (0.1%) | <0.001 | 0.33 (0.29 -0.37) |
| | Elevated Natriuretic peptide B | 793 (0.1%) | 40498 (0.3%) | <0.001 | 0.41 (0.38 -0.44) |
| | Dyslipidemia | 6567 (1.2%) | 393063 (2.9%) | <0.001 | 0.4 (0.39 -0.41) |
| | Triglyceride ≥150 mg/dl | 2553 (0.4%) | 95999 (0.6%) | <0.001 | 0.58 (0.56 -0.61) |
| | Total cholesterol ≥200 mg/dl | 3841 (0.6%) | 151207 (1%) | <0.001 | 0.57 (0.55 -0.59) |
| | LDL-cholesterol ≥130 mg/dl | 2966 (0.4%) | 109919 (0.7%) | <0.001 | 0.59 (0.57 -0.62) |
| | HDL-cholesterol <50 mg/dl | 3619 (0.5%) | 171734 (1.1%) | <0.001 | 0.47 (0.45 -0.48) |
| | Cerebrovascular disease | 983 (0.1%) | 51751 (0.3%) | <0.001 | 0.4 (0.37 -0.42) |
| | Transient ischemic attacks | 265 (0%) | 9097 (0.1%) | <0.001 | 0.61 (0.54 -0.68) |
| | Cerebral infarction | 367 (0%) | 20763 (0.1%) | <0.001 | 0.37 (0.33 -0.41) |
| | Cerebral occlusion without infarction | 332 (0%) | 15825 (0.1%) | <0.001 | 0.44 (0.39 -0.49) |
| | Intracerebral hemorrhage | 71 (0%) | 4563 (0%) | <0.001 | 0.32 (0.26 -0.41) |
| | Subarachnoid hemorrhage | 54 (0%) | 3332 (0%) | <0.001 | 0.34 (0.26 -0.44) |
| | Hospital admission | 3766 (0.6%) | 438054 (3.4%) | <0.001 | 0.17 (0.17 -0.18) |
| | Emergency visits | 6456 (1.2%) | 345713 (3.1%) | <0.001 | 0.37 (0.37 -0.38) |
| | Outpatient encounters | 9382 (2.2%) | 1003386 (9.7%) | <0.001 | 0.23 (0.22 -0.23) |
| | Intensive care unit admission | 143 (0%) | 39866 (0.2%) | <0.001 | 0.07 (0.06 -0.09) |
| | Mechanical ventilation | 269 (0%) | 26577 (0.2%) | <0.001 | 0.21 (0.19 -0.24) |
| | Referral to Cardio services | 0 (0%) | 41 (0%) | 0.159 | NA |
| | Referral to Neuro services | 10 (0%) | 276 (0%) | 0.371 | 0.75 (0.4 -1.41) |
| | ECMO | 10 (0%) | 317 (0%) | 0.182 | 0.65 (0.35 -1.23) |
| | ECMO-VV | 10 (0%) | 138 (0%) | 0.212 | 1.5 (0.79 -2.85) |
| | ECMO-VA | 10 (0%) | 213 (0%) | 0.931 | 0.97 (0.52 -1.83) |
| | Mortality | 1143 (0.1%) | 61041 (0.4%) | <0.001 | 0.39 (0.37 -0.41) |
| 3-6mo | Major adverse cardiovascular events | 1518 (0.2%) | 41758 (0.3%) | <0.001 | 0.76 (0.72 -0.8) |

| | | | | | |
|---|---|---|---|---|---|
| | Cardiovascular disease | 2054 (0.3%) | 58998 (0.4%) | <0.001 | 0.73 (0.7 -0.76) |
| | Ischemic heart disease | 899 (0.1%) | 23365 (0.1%) | <0.001 | 0.8 (0.75 -0.86) |
| | Myocardial angina | 146 (0%) | 2906 (0%) | 0.624 | 1.04 (0.88 -1.23) |
| | Acute coronary disease | 115 (0%) | 1974 (0%) | 0.05 | 1.21 (1 -1.46) |
| | Myocardial infarction | 263 (0%) | 5763 (0%) | 0.373 | 0.95 (0.84 -1.07) |
| | Ischemic cardiomyopathy | 57 (0%) | 1328 (0%) | 0.386 | 0.89 (0.68 -1.16) |
| | Arrythmias | 2877 (0.4%) | 73989 (0.5%) | <0.001 | 0.82 (0.79 -0.85) |
| | Atrial fibrillation and flutter | 508 (0.1%) | 13610 (0.1%) | <0.001 | 0.77 (0.71 -0.84) |
| | Paroxysmal tachycardia | 408 (0.1%) | 7721 (0%) | 0.069 | 1.1 (0.99 -1.21) |
| | Other cardiac arrythmias | 951 (0.1%) | 20730 (0.1%) | 0.211 | 0.96 (0.9 -1.02) |
| | Inflammatory heart disease | 204 (0%) | 4479 (0%) | 0.425 | 0.95 (0.82 -1.09) |
| | Pericarditis | 149 (0%) | 3187 (0%) | 0.707 | 0.97 (0.82 -1.14) |
| | Myocarditis | 10 (0%) | 138 (0%) | 0.212 | 1.5 (0.79 -2.85) |
| | Endocarditis | 58 (0%) | 1320 (0%) | 0.485 | 0.91 (0.7 -1.18) |
| | Hypertensive heart disease | 2767 (0.4%) | 105008 (0.8%) | <0.001 | 0.57 (0.55 -0.59) |
| | Pulmonary heart diseases | 447 (0.1%) | 9903 (0.1%) | 0.177 | 0.94 (0.85 -1.03) |
| | Heart Failure | 585 (0.1%) | 14970 (0.1%) | <0.001 | 0.81 (0.75 -0.88) |
| | Cardiomyopathy | 244 (0%) | 5245 (0%) | 0.585 | 0.97 (0.85 -1.1) |
| | Cardiomegaly | 447 (0.1%) | 9317 (0.1%) | 0.949 | 1 (0.91 -1.1) |
| | Cardiogenic shock | 46 (0%) | 956 (0%) | 0.982 | 1 (0.74 -1.34) |
| | Cardiac arrest | 101 (0%) | 2522 (0%) | 0.064 | 0.83 (0.68 -1.01) |
| | Thrombotic disorders | 421 (0.1%) | 10851 (0.1%) | <0.001 | 0.81 (0.73 -0.89) |
| | Pulmonary embolism | 198 (0%) | 4242 (0%) | 0.647 | 0.97 (0.84 -1.12) |
| | Deep and superficial thrombosis | 315 (0%) | 8272 (0.1%) | <0.001 | 0.79 (0.71 -0.88) |
| | Thrombosis with Thrombocytopenia | 0 (0%) | 0 (0%) | NA | NA |
| | Elevated Troponin I | 272 (0%) | 6348 (0%) | 0.052 | 0.89 (0.79 -1) |
| | Elevated Natriuretic peptide B | 741 (0.1%) | 15480 (0.1%) | 0.85 | 0.99 (0.92 -1.07) |
| | Dyslipidemia | 4775 (0.9%) | 146414 (1.1%) | <0.001 | 0.77 (0.74 -0.79) |
| | Triglyceride ≥150 mg/dl | 2034 (0.3%) | 40284 (0.3%) | <0.001 | 1.1 (1.05 -1.15) |
| | Total cholesterol ≥200 mg/dl | 2914 (0.4%) | 59974 (0.4%) | <0.001 | 1.09 (1.05 -1.13) |
| | LDL-cholesterol ≥130 mg/dl | 2286 (0.3%) | 44052 (0.3%) | <0.001 | 1.14 (1.09 -1.19) |
| | HDL-cholesterol <50 mg/dl | 2624 (0.4%) | 62967 (0.4%) | <0.001 | 0.92 (0.88 -0.95) |
| | Cerebrovascular disease | 844 (0.1%) | 20834 (0.1%) | <0.001 | 0.84 (0.79 -0.9) |
| | Transient ischemic attacks | 210 (0%) | 4312 (0%) | 0.877 | 1.01 (0.88 -1.16) |
| | Cerebral infarction | 315 (0%) | 8119 (0.1%) | <0.001 | 0.8 (0.72 -0.9) |
| | Cerebral occlusion without infarction | 270 (0%) | 6296 (0%) | 0.06 | 0.89 (0.79 -1.01) |
| | Intracerebral hemorrhage | 65 (0%) | 1394 (0%) | 0.782 | 0.97 (0.75 -1.24) |
| | Subarachnoid hemorrhage | 43 (0%) | 1015 (0%) | 0.4 | 0.88 (0.65 -1.19) |
| | Hospital admission | 3124 (0.5%) | 168270 (1.3%) | <0.001 | 0.37 (0.35 -0.38) |
| | Emergency visits | 5912 (1.1%) | 180608 (1.7%) | <0.001 | 0.64 (0.63 -0.66) |
| | Outpatient encounters | 6738 (1.6%) | 293344 (3.1%) | <0.001 | 0.52 (0.51 -0.53) |
| | Intensive care unit admission | 124 (0%) | 20602 (0.1%) | <0.001 | 0.12 (0.1 -0.15) |
| | Mechanical ventilation | 255 (0%) | 8428 (0.1%) | <0.001 | 0.63 (0.55 -0.71) |
| | Referral to Cardio services | 0 (0%) | 16 (0%) | 0.379 | NA |
| | Referral to Neuro services | 10 (0%) | 198 (0%) | 0.889 | 1.05 (0.55 -1.97) |
| | ECMO | 10 (0%) | 72 (0%) | 0.001 | 2.88 (1.49 -5.57) |
| | ECMO-VV | 10 (0%) | 30 (0%) | <0.001 | 6.9 (3.38 -14.12) |
| | ECMO-VA | 10 (0%) | 49 (0%) | <0.001 | 4.23 (2.14 -8.34) |
| | Mortality | 1372 (0.2%) | 32756 (0.2%) | <0.001 | 0.87 (0.82 -0.92) |
| 6-9mo | Major adverse cardiovascular events | 1443 (0.2%) | 33702 (0.2%) | <0.001 | 0.89 (0.85 -0.94) |
| | Cardiovascular disease | 1911 (0.3%) | 47035 (0.3%) | <0.001 | 0.85 (0.81 -0.89) |
| | Ischemic heart disease | 831 (0.1%) | 18723 (0.1%) | 0.023 | 0.92 (0.86 -0.99) |
| | Myocardial angina | 110 (0%) | 2436 (0%) | 0.504 | 0.94 (0.77 -1.13) |
| | Acute coronary disease | 95 (0%) | 1669 (0%) | 0.118 | 1.18 (0.96 -1.45) |
| | Myocardial infarction | 257 (0%) | 4995 (0%) | 0.319 | 1.07 (0.94 -1.21) |
| | Ischemic cardiomyopathy | 57 (0%) | 1037 (0%) | 0.339 | 1.14 (0.87 -1.49) |

| | | | | | |
|---|---|---|---|---|---|
| | Arrythmias | 2672 (0.4%) | 61731 (0.4%) | <0.001 | 0.91 (0.88 -0.95) |
| | Atrial fibrillation and flutter | 489 (0.1%) | 11010 (0.1%) | 0.069 | 0.92 (0.84 -1.01) |
| | Paroxysmal tachycardia | 353 (0%) | 6061 (0%) | 0.001 | 1.21 (1.09 -1.35) |
| | Other cardiac arrythmias | 844 (0.1%) | 16775 (0.1%) | 0.149 | 1.05 (0.98 -1.13) |
| | Inflammatory heart disease | 178 (0%) | 3564 (0%) | 0.648 | 1.04 (0.89 -1.2) |
| | Pericarditis | 135 (0%) | 2501 (0%) | 0.204 | 1.12 (0.94 -1.33) |
| | Myocarditis | 10 (0%) | 145 (0%) | 0.273 | 1.43 (0.75 -2.71) |
| | Endocarditis | 48 (0%) | 1045 (0%) | 0.738 | 0.95 (0.71 -1.27) |
| | Hypertensive heart disease | 2522 (0.4%) | 82432 (0.6%) | <0.001 | 0.66 (0.64 -0.69) |
| | Pulmonary heart diseases | 387 (0%) | 7743 (0%) | 0.482 | 1.04 (0.94 -1.15) |
| | Heart Failure | 569 (0.1%) | 11829 (0.1%) | 0.937 | 1 (0.92 -1.08) |
| | Cardiomyopathy | 226 (0%) | 4151 (0%) | 0.075 | 1.13 (0.99 -1.29) |
| | Cardiomegaly | 399 (0.1%) | 7935 (0%) | 0.392 | 1.05 (0.95 -1.16) |
| | Cardiogenic shock | 43 (0%) | 784 (0%) | 0.416 | 1.14 (0.84 -1.54) |
| | Cardiac arrest | 97 (0%) | 1955 (0%) | 0.797 | 1.03 (0.84 -1.26) |
| | Thrombotic disorders | 359 (0%) | 8539 (0.1%) | 0.011 | 0.87 (0.79 -0.97) |
| | Pulmonary embolism | 163 (0%) | 3318 (0%) | 0.824 | 1.02 (0.87 -1.19) |
| | Deep and superficial thrombosis | 259 (0%) | 6488 (0%) | 0.003 | 0.83 (0.73 -0.94) |
| | Thrombosis with Thrombocytopenia | 0 (0%) | 10 (0%) | 0.487 | NA |
| | Elevated Troponin I | 251 (0%) | 5798 (0%) | 0.087 | 0.9 (0.79 -1.02) |
| | Elevated Natriuretic peptide B | 696 (0.1%) | 12976 (0.1%) | 0.006 | 1.11 (1.03 -1.2) |
| | Dyslipidemia | 4298 (0.8%) | 117730 (0.9%) | <0.001 | 0.85 (0.83 -0.88) |
| | Triglyceride ≥150 mg/dl | 1966 (0.3%) | 35634 (0.2%) | <0.001 | 1.2 (1.15 -1.26) |
| | Total cholesterol ≥200 mg/dl | 2773 (0.4%) | 52594 (0.4%) | <0.001 | 1.18 (1.14 -1.23) |
| | LDL-cholesterol ≥130 mg/dl | 2232 (0.3%) | 39269 (0.3%) | <0.001 | 1.25 (1.19 -1.3) |
| | HDL-cholesterol <50 mg/dl | 2432 (0.4%) | 53480 (0.4%) | 0.927 | 1 (0.96 -1.04) |
| | Cerebrovascular disease | 785 (0.1%) | 17166 (0.1%) | 0.161 | 0.95 (0.89 -1.02) |
| | Transient ischemic attacks | 208 (0%) | 3737 (0%) | 0.042 | 1.16 (1.01 -1.33) |
| | Cerebral infarction | 322 (0%) | 6614 (0%) | 0.883 | 1.01 (0.9 -1.13) |
| | Cerebral occlusion without infarction | 266 (0%) | 5185 (0%) | 0.32 | 1.07 (0.94 -1.2) |
| | Intracerebral hemorrhage | 69 (0%) | 1143 (0%) | 0.071 | 1.25 (0.98 -1.59) |
| | Subarachnoid hemorrhage | 43 (0%) | 842 (0%) | 0.72 | 1.06 (0.78 -1.44) |
| | Hospital admission | 2578 (0.4%) | 142052 (1.1%) | <0.001 | 0.35 (0.34 -0.37) |
| | Emergency visits | 5372 (1%) | 154301 (1.5%) | <0.001 | 0.68 (0.66 -0.7) |
| | Outpatient encounters | 5866 (1.4%) | 193404 (2.1%) | <0.001 | 0.67 (0.66 -0.69) |
| | Intensive care unit admission | 79 (0%) | 15706 (0.1%) | <0.001 | 0.1 (0.08 -0.13) |
| | Mechanical ventilation | 248 (0%) | 6477 (0%) | <0.001 | 0.79 (0.7 -0.9) |
| | Referral to Cardio services | 0 (0%) | 10 (0%) | 0.487 | NA |
| | Referral to Neuro services | 10 (0%) | 206 (0%) | 0.987 | 1.01 (0.53 -1.9) |
| | ECMO | 0 (0%) | 42 (0%) | 0.154 | NA |
| | ECMO-VV | 0 (0%) | 15 (0%) | 0.395 | NA |
| | ECMO-VA | 0 (0%) | 27 (0%) | 0.254 | NA |
| | Mortality | 1441 (0.2%) | 28948 (0.2%) | 0.258 | 1.03 (0.98 -1.09) |
| >9mo | Major adverse cardiovascular events | 11344 (1.5%) | 222559 (1.5%) | <0.001 | 1.06 (1.04 -1.08) |
| | Cardiovascular disease | 15909 (2.2%) | 305195 (2%) | <0.001 | 1.09 (1.07 -1.11) |
| | Ischemic heart disease | 6671 (0.9%) | 125099 (0.8%) | <0.001 | 1.11 (1.08 -1.14) |
| | Myocardial angina | 957 (0.1%) | 16170 (0.1%) | <0.001 | 1.23 (1.15 -1.31) |
| | Acute coronary disease | 791 (0.1%) | 10834 (0.1%) | <0.001 | 1.51 (1.41 -1.63) |
| | Myocardial infarction | 1888 (0.2%) | 34809 (0.2%) | <0.001 | 1.12 (1.07 -1.18) |
| | Ischemic cardiomyopathy | 304 (0%) | 5701 (0%) | 0.09 | 1.11 (0.98 -1.24) |
| | Arrythmias | 22335 (3.2%) | 433163 (2.9%) | <0.001 | 1.09 (1.07 -1.1) |
| | Atrial fibrillation and flutter | 3432 (0.4%) | 66729 (0.4%) | <0.001 | 1.07 (1.03 -1.1) |
| | Paroxysmal tachycardia | 2751 (0.4%) | 40612 (0.3%) | <0.001 | 1.41 (1.35 -1.46) |
| | Other cardiac arrythmias | 6640 (0.9%) | 108798 (0.7%) | <0.001 | 1.28 (1.25 -1.31) |
| | Inflammatory heart disease | 1534 (0.2%) | 23197 (0.1%) | <0.001 | 1.37 (1.3 -1.44) |
| | Pericarditis | 1009 (0.1%) | 15709 (0.1%) | <0.001 | 1.33 (1.25 -1.42) |

| | | | | | |
|---|---|---|---|---|---|
| | Myocarditis | 33 (0%) | 760 (0%) | 0.55 | 0.9 (0.64 -1.27) |
| | Endocarditis | 554 (0.1%) | 7586 (0%) | <0.001 | 1.51 (1.39 -1.65) |
| | Hypertensive heart disease | 25063 (3.9%) | 565850 (4.1%) | <0.001 | 0.96 (0.94 -0.97) |
| | Pulmonary heart diseases | 2820 (0.4%) | 48801 (0.3%) | <0.001 | 1.2 (1.16 -1.25) |
| | Heart Failure | 3942 (0.5%) | 74030 (0.5%) | <0.001 | 1.1 (1.07 -1.14) |
| | Cardiomyopathy | 1412 (0.2%) | 24094 (0.1%) | <0.001 | 1.22 (1.15 -1.28) |
| | Cardiomegaly | 2853 (0.4%) | 56299 (0.4%) | 0.007 | 1.05 (1.01 -1.09) |
| | Cardiogenic shock | 413 (0.1%) | 5524 (0%) | <0.001 | 1.55 (1.4 -1.71) |
| | Cardiac arrest | 861 (0.1%) | 14441 (0.1%) | <0.001 | 1.23 (1.15 -1.32) |
| | Thrombotic disorders | 3158 (0.4%) | 58649 (0.4%) | <0.001 | 1.12 (1.08 -1.16) |
| | Pulmonary embolism | 1140 (0.1%) | 20835 (0.1%) | <0.001 | 1.13 (1.07 -1.2) |
| | Deep and superficial thrombosis | 2540 (0.3%) | 46156 (0.3%) | <0.001 | 1.14 (1.1 -1.19) |
| | Thrombosis with Thrombocytopenia | 0 (0%) | 12 (0%) | 0.447 | NA |
| | Elevated Troponin I | 3482 (0.4%) | 56040 (0.3%) | <0.001 | 1.29 (1.24 -1.33) |
| | Elevated Natriuretic peptide B | 5212 (0.7%) | 95448 (0.6%) | <0.001 | 1.13 (1.1 -1.17) |
| | Dyslipidemia | 43433 (7.9%) | 932017 (7.3%) | <0.001 | 1.09 (1.08 -1.1) |
| | Triglyceride ≥150 mg/dl | 18594 (2.7%) | 309702 (2%) | <0.001 | 1.31 (1.29 -1.33) |
| | Total cholesterol ≥200 mg/dl | 26429 (4%) | 481858 (3.3%) | <0.001 | 1.23 (1.22 -1.25) |
| | LDL-cholesterol ≥130 mg/dl | 21914 (3.2%) | 367603 (2.4%) | <0.001 | 1.31 (1.29 -1.33) |
| | HDL-cholesterol <50 mg/dl | 22507 (3.3%) | 460654 (3.1%) | <0.001 | 1.07 (1.06 -1.09) |
| | Cerebrovascular disease | 6356 (0.8%) | 117698 (0.7%) | <0.001 | 1.12 (1.09 -1.15) |
| | Transient ischemic attacks | 1800 (0.2%) | 29717 (0.2%) | <0.001 | 1.26 (1.2 -1.32) |
| | Cerebral infarction | 2273 (0.3%) | 42891 (0.3%) | <0.001 | 1.1 (1.05 -1.15) |
| | Cerebral occlusion without infarction | 1946 (0.3%) | 35478 (0.2%) | <0.001 | 1.14 (1.09 -1.19) |
| | Intracerebral hemorrhage | 501 (0.1%) | 8001 (0%) | <0.001 | 1.3 (1.19 -1.42) |
| | Subarachnoid hemorrhage | 369 (0%) | 6152 (0%) | <0.001 | 1.24 (1.12 -1.38) |
| | Hospital admission | 22730 (3.6%) | 650921 (5.3%) | <0.001 | 0.68 (0.67 -0.69) |
| | Emergency visits | 47540 (8.9%) | 1017137 (9.8%) | <0.001 | 0.91 (0.9 -0.92) |
| | Outpatient encounters | 43557 (10.8%) | 977358 (11%) | <0.001 | 0.98 (0.97 -0.99) |
| | Intensive care unit admission | 381 (0%) | 50446 (0.3%) | <0.001 | 0.16 (0.14 -0.17) |
| | Mechanical ventilation | 1985 (0.3%) | 43187 (0.3%) | 0.023 | 0.95 (0.91 -0.99) |
| | Referral to Cardio services | 0 (0%) | 56 (0%) | 0.1 | NA |
| | Referral to Neuro services | 34 (0%) | 2531 (0%) | <0.001 | 0.28 (0.2 -0.39) |
| | ECMO | 17 (0%) | 310 (0%) | 0.609 | 1.14 (0.7 -1.85) |
| | ECMO-VV | 10 (0%) | 103 (0%) | 0.031 | 2.01 (1.05 -3.85) |
| | ECMO-VA | 11 (0%) | 224 (0%) | 0.956 | 1.02 (0.56 -1.86) |
| | Mortality | 13581 (1.7%) | 201545 (1.2%) | <0.001 | 1.4 (1.37 -1.42) |

For outcomes where event counts were fewer than 10, TriNetX platform suppressed exact numbers and reported '10' for de-identification purposes; thus, relative risks may be overestimated or not statistically meaningful in these cases. When zero events were observed, this is indicated as '0'. Caution is warranted in the interpretation of RRs where numerators are at or below this privacy threshold. ECMO: Extracorporeal membrane oxygenation.

**Table S7.** Outcomes in male in infected, vaccinated individuals compared to infected, unvaccinated individuals (Group 4 vs Group 2) across four post-exposure time windows.

| Time | Outcome | Group 4 (n=61,264) | Group 2 (n=858,409) | *p*-value | RR (95%CI) |
|---|---|---|---|---|---|
| | **Follow-up (months)** | **22.7 ± 18.8** | **22 ± 16.2** | | |
| 0-3 mo | Major adverse cardiovascular events | 541 (0.8%) | 9734 (1.2%) | <0.001 | 0.64 (0.59 -0.7) |
| | Cardiovascular disease | 659 (1%) | 14276 (1.8%) | <0.001 | 0.54 (0.5 -0.58) |
| | Ischemic heart disease | 432 (0.6%) | 6930 (0.8%) | <0.001 | 0.71 (0.65 -0.79) |
| | Myocardial angina | 63 (0.1%) | 657 (0.1%) | 0.565 | 1.08 (0.83 -1.4) |
| | Acute coronary disease | 136 (0.2%) | 1272 (0.1%) | 0.046 | 1.2 (0.99 -1.43) |
| | Myocardial infarction | 212 (0.3%) | 3145 (0.3%) | <0.001 | 0.76 (0.66 -0.87) |
| | Ischemic cardiomyopathy | 65 (0.1%) | 903 (0.1%) | 0.093 | 0.81 (0.63 -1.04) |
| | Arrythmias | 851 (1.3%) | 16595 (2.1%) | <0.001 | 0.6 (0.56 -0.65) |
| | Atrial fibrillation and flutter | 254 (0.3%) | 4718 (0.5%) | <0.001 | 0.6 (0.53 -0.69) |
| | Paroxysmal tachycardia | 194 (0.2%) | 2462 (0.3%) | 0.106 | 0.89 (0.77 -1.03) |
| | Other cardiac arrythmias | 489 (0.6%) | 7309 (0.8%) | <0.001 | 0.77 (0.71 -0.85) |
| | Inflammatory heart disease | 138 (0.2%) | 2224 (0.2%) | <0.001 | 0.7 (0.59 -0.83) |
| | Pericarditis | 109 (0.1%) | 1454 (0.2%) | 0.079 | 0.84 (0.69 -1.02) |
| | Myocarditis | 11 (0%) | 269 (0%) | 0.009 | 0.46 (0.25 -0.83) |
| | Endocarditis | 29 (0%) | 695 (0.1%) | <0.001 | 0.47 (0.32 -0.68) |
| | Hypertensive heart disease | 539 (1%) | 12317 (1.8%) | <0.001 | 0.56 (0.52 -0.61) |
| | Pulmonary heart diseases | 249 (0.3%) | 4598 (0.5%) | <0.001 | 0.61 (0.54 -0.69) |
| | Heart Failure | 310 (0.4%) | 4685 (0.5%) | <0.001 | 0.74 (0.66 -0.83) |
| | Cardiomyopathy | 151 (0.2%) | 2273 (0.2%) | <0.001 | 0.75 (0.63 -0.88) |
| | Cardiomegaly | 360 (0.4%) | 5365 (0.6%) | <0.001 | 0.76 (0.68 -0.85) |
| | Cardiogenic shock | 65 (0.1%) | 745 (0.1%) | 0.839 | 0.97 (0.76 -1.26) |
| | Cardiac arrest | 109 (0.1%) | 1783 (0.2%) | <0.001 | 0.68 (0.56 -0.83) |
| | Thrombotic disorders | 259 (0.3%) | 5658 (0.6%) | <0.001 | 0.52 (0.46 -0.58) |
| | Pulmonary embolism | 123 (0.1%) | 3101 (0.3%) | <0.001 | 0.44 (0.37 -0.53) |
| | Deep and superficial thrombosis | 192 (0.2%) | 3685 (0.4%) | <0.001 | 0.59 (0.51 -0.68) |
| | Thrombosis with Thrombocytopenia | 0 (0%) | 0 (0%) | NA | NA |
| | Elevated Troponin I | 221 (0.3%) | 4753 (0.5%) | <0.001 | 0.52 (0.45 -0.59) |
| | Elevated Natriuretic peptide B | 471 (0.6%) | 10154 (1.1%) | <0.001 | 0.52 (0.48 -0.57) |
| | Dyslipidemia | 1108 (3.1%) | 21833 (3.7%) | <0.001 | 0.85 (0.8 -0.9) |
| | Triglyceride ≥150 mg/dl | 756 (1.2%) | 12731 (1.6%) | <0.001 | 0.78 (0.73 -0.84) |
| | Total cholesterol ≥200 mg/dl | 604 (1%) | 6539 (0.8%) | <0.001 | 1.19 (1.1 -1.3) |
| | LDL-cholesterol ≥130 mg/dl | 553 (0.8%) | 5817 (0.7%) | <0.001 | 1.2 (1.1 -1.31) |
| | HDL-cholesterol <50 mg/dl | 1125 (2.4%) | 19147 (2.7%) | <0.001 | 0.87 (0.82 -0.92) |
| | Cerebrovascular disease | 259 (0.3%) | 4220 (0.5%) | <0.001 | 0.7 (0.61 -0.79) |
| | Transient ischemic attacks | 72 (0.1%) | 827 (0.1%) | 0.865 | 0.98 (0.77 -1.25) |
| | Cerebral infarction | 149 (0.2%) | 2121 (0.2%) | 0.005 | 0.79 (0.67 -0.93) |
| | Cerebral occlusion without infarction | 94 (0.1%) | 1589 (0.2%) | <0.001 | 0.67 (0.54 -0.82) |
| | Intracerebral hemorrhage | 24 (0%) | 563 (0.1%) | <0.001 | 0.48 (0.32 -0.72) |
| | Subarachnoid hemorrhage | 22 (0%) | 313 (0%) | 0.271 | 0.79 (0.51 -1.21) |
| | Hospital admission | 719 (1.2%) | 19795 (2.9%) | <0.001 | 0.41 (0.38 -0.44) |
| | Emergency visits | 936 (2.3%) | 17873 (4.4%) | <0.001 | 0.53 (0.5 -0.57) |
| | Outpatient encounters | 694 (2.4%) | 22256 (4.8%) | <0.001 | 0.5 (0.47 -0.54) |
| | Intensive care unit admission | 23 (0%) | 1748 (0.2%) | <0.001 | 0.14 (0.1 -0.22) |
| | Mechanical ventilation | 517 (0.6%) | 11014 (1.2%) | <0.001 | 0.52 (0.48 -0.57) |
| | Referral to Cardio services | 0 (0%) | 10 (0%) | 0.344 | NA |
| | Referral to Neuro services | 0 (0%) | 28 (0%) | 0.113 | NA |
| | ECMO | 10 (0%) | 298 (0%) | 0.001 | 0.38 (0.2 -0.7) |
| | ECMO-VV | 10 (0%) | 252 (0%) | 0.009 | 0.44 (0.24 -0.83) |
| | ECMO-VA | 10 (0%) | 115 (0%) | 0.928 | 0.97 (0.51 -1.85) |
| | Mortality | 1375 (1.6%) | 25690 (2.7%) | <0.001 | 0.6 (0.57 -0.63) |
| 3-6mo | Major adverse cardiovascular events | 340 (0.5%) | 3359 (0.4%) | 0.01 | 1.16 (1.04 -1.29) |

| | | | | | |
|---|---|---|---|---|---|
| | Cardiovascular disease | 393 (0.6%) | 4056 (0.5%) | 0.029 | 1.12 (1.01 -1.24) |
| | Ischemic heart disease | 233 (0.3%) | 2374 (0.3%) | 0.096 | 1.12 (0.98 -1.28) |
| | Myocardial angina | 46 (0.1%) | 353 (0%) | 0.014 | 1.47 (1.08 -1.99) |
| | Acute coronary disease | 45 (0.1%) | 421 (0%) | 0.252 | 1.2 (0.88 -1.63) |
| | Myocardial infarction | 94 (0.1%) | 950 (0.1%) | 0.336 | 1.11 (0.9 -1.37) |
| | Ischemic cardiomyopathy | 37 (0%) | 324 (0%) | 0.155 | 1.28 (0.91 -1.8) |
| | Arrythmias | 482 (0.7%) | 5145 (0.7%) | 0.059 | 1.09 (0.99 -1.2) |
| | Atrial fibrillation and flutter | 128 (0.2%) | 1349 (0.2%) | 0.516 | 1.06 (0.89 -1.27) |
| | Paroxysmal tachycardia | 116 (0.1%) | 780 (0.1%) | <0.001 | 1.67 (1.38 -2.03) |
| | Other cardiac arrythmias | 248 (0.3%) | 2004 (0.2%) | <0.001 | 1.43 (1.25 -1.63) |
| | Inflammatory heart disease | 64 (0.1%) | 616 (0.1%) | 0.244 | 1.17 (0.9 -1.51) |
| | Pericarditis | 46 (0.1%) | 410 (0%) | 0.14 | 1.26 (0.93 -1.71) |
| | Myocarditis | 10 (0%) | 42 (0%) | 0.004 | 2.66 (1.33 -5.3) |
| | Endocarditis | 18 (0%) | 216 (0%) | 0.772 | 0.93 (0.58 -1.51) |
| | Hypertensive heart disease | 393 (0.7%) | 4962 (0.7%) | 0.825 | 1.01 (0.91 -1.12) |
| | Pulmonary heart diseases | 99 (0.1%) | 1031 (0.1%) | 0.468 | 1.08 (0.88 -1.33) |
| | Heart Failure | 164 (0.2%) | 1531 (0.2%) | 0.025 | 1.2 (1.02 -1.41) |
| | Cardiomyopathy | 73 (0.1%) | 708 (0.1%) | 0.232 | 1.16 (0.91 -1.47) |
| | Cardiomegaly | 168 (0.2%) | 1235 (0.1%) | <0.001 | 1.54 (1.31 -1.81) |
| | Cardiogenic shock | 34 (0%) | 237 (0%) | 0.01 | 1.6 (1.12 -2.29) |
| | Cardiac arrest | 60 (0.1%) | 545 (0.1%) | 0.133 | 1.23 (0.94 -1.6) |
| | Thrombotic disorders | 104 (0.1%) | 1068 (0.1%) | 0.39 | 1.09 (0.89 -1.34) |
| | Pulmonary embolism | 35 (0%) | 453 (0%) | 0.4 | 0.86 (0.61 -1.22) |
| | Deep and superficial thrombosis | 94 (0.1%) | 855 (0.1%) | 0.053 | 1.23 (1 -1.53) |
| | Thrombosis with Thrombocytopenia | 0 (0%) | 0 (0%) | NA | NA |
| | Elevated Troponin I | 81 (0.1%) | 1161 (0.1%) | 0.026 | 0.77 (0.62 -0.97) |
| | Elevated Natriuretic peptide B | 183 (0.2%) | 1619 (0.2%) | 0.002 | 1.27 (1.09 -1.48) |
| | Dyslipidemia | 659 (1.9%) | 8106 (1.4%) | <0.001 | 1.35 (1.25 -1.46) |
| | Triglyceride ≥150 mg/dl | 447 (0.7%) | 4438 (0.6%) | <0.001 | 1.32 (1.2 -1.45) |
| | Total cholesterol ≥200 mg/dl | 450 (0.7%) | 4041 (0.5%) | <0.001 | 1.44 (1.31 -1.59) |
| | LDL-cholesterol ≥130 mg/dl | 404 (0.6%) | 3583 (0.4%) | <0.001 | 1.42 (1.28 -1.58) |
| | HDL-cholesterol <50 mg/dl | 663 (1.4%) | 6865 (1%) | <0.001 | 1.42 (1.32 -1.54) |
| | Cerebrovascular disease | 184 (0.2%) | 1646 (0.2%) | 0.003 | 1.26 (1.09 -1.47) |
| | Transient ischemic attacks | 55 (0.1%) | 402 (0%) | 0.003 | 1.54 (1.16 -2.04) |
| | Cerebral infarction | 56 (0.1%) | 719 (0.1%) | 0.324 | 0.87 (0.67 -1.15) |
| | Cerebral occlusion without infarction | 78 (0.1%) | 571 (0.1%) | <0.001 | 1.54 (1.22 -1.95) |
| | Intracerebral hemorrhage | 17 (0%) | 179 (0%) | 0.819 | 1.06 (0.65 -1.74) |
| | Subarachnoid hemorrhage | 22 (0%) | 103 (0%) | <0.001 | 2.39 (1.51 -3.78) |
| | Hospital admission | 418 (0.7%) | 5377 (0.8%) | 0.003 | 0.86 (0.78 -0.95) |
| | Emergency visits | 683 (1.7%) | 7898 (2%) | <0.001 | 0.86 (0.8 -0.93) |
| | Outpatient encounters | 343 (1.2%) | 8296 (1.9%) | <0.001 | 0.65 (0.58 -0.72) |
| | Intensive care unit admission | 12 (0%) | 514 (0.1%) | <0.001 | 0.26 (0.14 -0.45) |
| | Mechanical ventilation | 140 (0.2%) | 1100 (0.1%) | <0.001 | 1.41 (1.18 -1.68) |
| | Referral to Cardio services | 0 (0%) | 0 (0%) | NA | NA |
| | Referral to Neuro services | 0 (0%) | 10 (0%) | 0.344 | NA |
| | ECMO | 10 (0%) | 10 (0%) | <0.001 | 11.16 (4.65 - 26.82) |
| | ECMO-VV | 10 (0%) | 10 (0%) | <0.001 | 11.16 (4.65 - 26.82) |
| | ECMO-VA | 0 (0%) | 10 (0%) | 0.344 | NA |
| | Mortality | 569 (0.7%) | 5145 (0.5%) | <0.001 | 1.24 (1.13 -1.35) |
| 6-9mo | Major adverse cardiovascular events | 263 (0.4%) | 2638 (0.3%) | 0.041 | 1.14 (1.01 -1.29) |
| | Cardiovascular disease | 315 (0.5%) | 3276 (0.4%) | 0.066 | 1.11 (0.99 -1.25) |
| | Ischemic heart disease | 190 (0.3%) | 1812 (0.2%) | 0.018 | 1.2 (1.03 -1.39) |
| | Myocardial angina | 36 (0%) | 287 (0%) | 0.05 | 1.41 (1 -2) |
| | Acute coronary disease | 50 (0.1%) | 313 (0%) | <0.001 | 1.79 (1.33 -2.41) |

| | | | | | |
|---|---|---|---|---|---|
| | Myocardial infarction | 92 (0.1%) | 660 (0.1%) | <0.001 | 1.56 (1.26 -1.94) |
| | Ischemic cardiomyopathy | 32 (0%) | 229 (0%) | 0.017 | 1.57 (1.08 -2.27) |
| | Arrythmias | 414 (0.6%) | 4269 (0.6%) | 0.015 | 1.13 (1.03 -1.25) |
| | Atrial fibrillation and flutter | 105 (0.1%) | 1024 (0.1%) | 0.179 | 1.15 (0.94 -1.4) |
| | Paroxysmal tachycardia | 83 (0.1%) | 617 (0.1%) | <0.001 | 1.51 (1.2 -1.9) |
| | Other cardiac arrythmias | 182 (0.2%) | 1601 (0.2%) | 0.001 | 1.31 (1.12 -1.53) |
| | Inflammatory heart disease | 53 (0.1%) | 388 (0%) | 0.003 | 1.53 (1.15 -2.04) |
| | Pericarditis | 42 (0.1%) | 251 (0%) | <0.001 | 1.88 (1.35 -2.6) |
| | Myocarditis | 10 (0%) | 30 (0%) | <0.001 | 3.72 (1.82 -7.61) |
| | Endocarditis | 14 (0%) | 144 (0%) | 0.767 | 1.09 (0.63 -1.88) |
| | Hypertensive heart disease | 330 (0.6%) | 4079 (0.6%) | 0.564 | 1.03 (0.92 -1.16) |
| | Pulmonary heart diseases | 88 (0.1%) | 732 (0.1%) | 0.007 | 1.35 (1.08 -1.69) |
| | Heart Failure | 123 (0.2%) | 1156 (0.1%) | 0.061 | 1.19 (0.99 -1.44) |
| | Cardiomyopathy | 73 (0.1%) | 543 (0.1%) | 0.001 | 1.51 (1.18 -1.93) |
| | Cardiomegaly | 124 (0.2%) | 905 (0.1%) | <0.001 | 1.55 (1.29 -1.87) |
| | Cardiogenic shock | 27 (0%) | 185 (0%) | 0.017 | 1.63 (1.09 -2.44) |
| | Cardiac arrest | 55 (0.1%) | 341 (0%) | <0.001 | 1.8 (1.35 -2.39) |
| | Thrombotic disorders | 93 (0.1%) | 807 (0.1%) | 0.019 | 1.29 (1.04 -1.6) |
| | Pulmonary embolism | 33 (0%) | 350 (0%) | 0.776 | 1.05 (0.74 -1.51) |
| | Deep and superficial thrombosis | 82 (0.1%) | 638 (0.1%) | 0.002 | 1.44 (1.15 -1.82) |
| | Thrombosis with Thrombocytopenia | 0 (0%) | 0 (0%) | NA | NA |
| | Elevated Troponin I | 78 (0.1%) | 969 (0.1%) | 0.336 | 0.89 (0.71 -1.13) |
| | Elevated Natriuretic peptide B | 155 (0.2%) | 1245 (0.1%) | <0.001 | 1.4 (1.18 -1.65) |
| | Dyslipidemia | 602 (1.8%) | 6756 (1.2%) | <0.001 | 1.48 (1.37 -1.61) |
| | Triglyceride ≥150 mg/dl | 458 (0.8%) | 3767 (0.5%) | <0.001 | 1.6 (1.45 -1.76) |
| | Total cholesterol ≥200 mg/dl | 414 (0.7%) | 3648 (0.5%) | <0.001 | 1.47 (1.33 -1.63) |
| | LDL-cholesterol ≥130 mg/dl | 394 (0.6%) | 3116 (0.4%) | <0.001 | 1.6 (1.44 -1.77) |
| | HDL-cholesterol <50 mg/dl | 599 (1.3%) | 5852 (0.9%) | <0.001 | 1.52 (1.39 -1.65) |
| | Cerebrovascular disease | 169 (0.2%) | 1372 (0.2%) | <0.001 | 1.39 (1.19 -1.64) |
| | Transient ischemic attacks | 51 (0.1%) | 357 (0%) | 0.001 | 1.61 (1.2 -2.16) |
| | Cerebral infarction | 79 (0.1%) | 610 (0.1%) | 0.002 | 1.45 (1.15 -1.83) |
| | Cerebral occlusion without infarction | 69 (0.1%) | 518 (0.1%) | 0.001 | 1.5 (1.17 -1.93) |
| | Intracerebral hemorrhage | 21 (0%) | 141 (0%) | 0.028 | 1.66 (1.05 -2.63) |
| | Subarachnoid hemorrhage | 16 (0%) | 91 (0%) | 0.011 | 1.96 (1.15 -3.34) |
| | Hospital admission | 397 (0.7%) | 4630 (0.7%) | 0.317 | 0.95 (0.86 -1.05) |
| | Emergency visits | 663 (1.7%) | 6926 (1.8%) | 0.229 | 0.95 (0.88 -1.03) |
| | Outpatient encounters | 321 (1.2%) | 6480 (1.5%) | <0.001 | 0.77 (0.69 -0.86) |
| | Intensive care unit admission | 18 (0%) | 420 (0%) | 0.001 | 0.47 (0.29 -0.75) |
| | Mechanical ventilation | 117 (0.1%) | 827 (0.1%) | <0.001 | 1.57 (1.29 -1.9) |
| | Referral to Cardio services | 0 (0%) | 0 (0%) | NA | NA |
| | Referral to Neuro services | 10 (0%) | 50 (0%) | 0.017 | 2.23 (1.13 -4.4) |
| | ECMO | 10 (0%) | 45 (0%) | 0.007 | 2.48 (1.25 -4.92) |
| | ECMO-VV | 10 (0%) | 15 (0%) | <0.001 | 7.44 (3.34 -16.57) |
| | ECMO-VA | 10 (0%) | 37 (0%) | 0.001 | 3.02 (1.5 -6.07) |
| | Mortality | 466 (0.6%) | 3737 (0.4%) | <0.001 | 1.39 (1.27 -1.53) |
| >9mo | Major adverse cardiovascular events | 1292 (1.8%) | 13468 (1.7%) | 0.001 | 1.1 (1.04 -1.16) |
| | Cardiovascular disease | 1632 (2.5%) | 16782 (2.2%) | <0.001 | 1.13 (1.07 -1.19) |
| | Ischemic heart disease | 992 (1.3%) | 9031 (1.1%) | <0.001 | 1.26 (1.18 -1.34) |
| | Myocardial angina | 164 (0.2%) | 1374 (0.1%) | <0.001 | 1.34 (1.14 -1.58) |
| | Acute coronary disease | 188 (0.2%) | 1270 (0.1%) | <0.001 | 1.66 (1.42 -1.93) |
| | Myocardial infarction | 350 (0.4%) | 3152 (0.3%) | <0.001 | 1.25 (1.12 -1.39) |
| | Ischemic cardiomyopathy | 92 (0.1%) | 913 (0.1%) | 0.268 | 1.13 (0.91 -1.4) |
| | Arrythmias | 2163 (3.3%) | 22281 (2.9%) | <0.001 | 1.14 (1.09 -1.19) |
| | Atrial fibrillation and flutter | 460 (0.6%) | 4616 (0.5%) | 0.025 | 1.12 (1.01 -1.23) |
| | Paroxysmal tachycardia | 362 (0.4%) | 2589 (0.3%) | <0.001 | 1.57 (1.41 -1.76) |

| | | | | | |
|---|---|---|---|---|---|
| | Other cardiac arrythmias | 929 (1.2%) | 7456 (0.8%) | <0.001 | 1.44 (1.34 -1.54) |
| | Inflammatory heart disease | 270 (0.3%) | 1820 (0.2%) | <0.001 | 1.66 (1.47 -1.89) |
| | Pericarditis | 191 (0.2%) | 1189 (0.1%) | <0.001 | 1.8 (1.55 -2.1) |
| | Myocarditis | 12 (0%) | 110 (0%) | 0.516 | 1.22 (0.67 -2.21) |
| | Endocarditis | 91 (0.1%) | 680 (0.1%) | <0.001 | 1.5 (1.2 -1.86) |
| | Hypertensive heart disease | 2410 (4.6%) | 26569 (4%) | <0.001 | 1.16 (1.11 -1.21) |
| | Pulmonary heart diseases | 389 (0.5%) | 3132 (0.3%) | <0.001 | 1.4 (1.26 -1.55) |
| | Heart Failure | 533 (0.7%) | 5113 (0.6%) | 0.001 | 1.17 (1.07 -1.28) |
| | Cardiomyopathy | 261 (0.3%) | 2237 (0.2%) | <0.001 | 1.31 (1.15 -1.49) |
| | Cardiomegaly | 534 (0.7%) | 4562 (0.5%) | <0.001 | 1.33 (1.21 -1.45) |
| | Cardiogenic shock | 112 (0.1%) | 724 (0.1%) | <0.001 | 1.73 (1.42 -2.11) |
| | Cardiac arrest | 239 (0.3%) | 2071 (0.2%) | <0.001 | 1.29 (1.13 -1.47) |
| | Thrombotic disorders | 478 (0.6%) | 4097 (0.5%) | <0.001 | 1.31 (1.19 -1.44) |
| | Pulmonary embolism | 188 (0.2%) | 1444 (0.2%) | <0.001 | 1.45 (1.25 -1.69) |
| | Deep and superficial thrombosis | 390 (0.5%) | 3401 (0.4%) | <0.001 | 1.29 (1.16 -1.43) |
| | Thrombosis with Thrombocytopenia | 10 (0%) | 0 (0%) | <0.001 | NA |
| | Elevated Troponin I | 452 (0.5%) | 6196 (0.7%) | <0.001 | 0.81 (0.74 -0.89) |
| | Elevated Natriuretic peptide B | 697 (0.9%) | 6022 (0.7%) | <0.001 | 1.3 (1.2 -1.41) |
| | Dyslipidemia | 3787 (11.4%) | 42458 (7.6%) | <0.001 | 1.49 (1.45 -1.54) |
| | Triglyceride ≥150 mg/dl | 2895 (4.9%) | 24890 (3.2%) | <0.001 | 1.53 (1.47 -1.59) |
| | Total cholesterol ≥200 mg/dl | 2751 (4.5%) | 24015 (3%) | <0.001 | 1.49 (1.43 -1.55) |
| | LDL-cholesterol ≥130 mg/dl | 2541 (3.9%) | 21815 (2.7%) | <0.001 | 1.47 (1.42 -1.54) |
| | HDL-cholesterol <50 mg/dl | 3679 (8.1%) | 36493 (5.4%) | <0.001 | 1.5 (1.45 -1.55) |
| | Cerebrovascular disease | 728 (0.9%) | 6384 (0.7%) | <0.001 | 1.29 (1.2 -1.39) |
| | Transient ischemic attacks | 240 (0.3%) | 1796 (0.2%) | <0.001 | 1.5 (1.32 -1.72) |
| | Cerebral infarction | 264 (0.3%) | 2481 (0.3%) | 0.007 | 1.19 (1.05 -1.35) |
| | Cerebral occlusion without infarction | 296 (0.4%) | 2304 (0.2%) | <0.001 | 1.45 (1.28 -1.63) |
| | Intracerebral hemorrhage | 68 (0.1%) | 547 (0.1%) | 0.011 | 1.39 (1.08 -1.79) |
| | Subarachnoid hemorrhage | 54 (0.1%) | 381 (0%) | 0.001 | 1.58 (1.19 -2.11) |
| | Hospital admission | 2267 (3.8%) | 30805 (4.7%) | <0.001 | 0.81 (0.78 -0.85) |
| | Emergency visits | 4083 (10.8%) | 41491 (11%) | 0.161 | 0.98 (0.95 -1.01) |
| | Outpatient encounters | 1592 (5.8%) | 34896 (8.3%) | <0.001 | 0.71 (0.67 -0.74) |
| | Intensive care unit admission | 47 (0.1%) | 1731 (0.2%) | <0.001 | 0.3 (0.22 -0.4) |
| | Mechanical ventilation | 505 (0.6%) | 4143 (0.5%) | <0.001 | 1.35 (1.23 -1.48) |
| | Referral to Cardio services | 0 (0%) | 0 (0%) | NA | NA |
| | Referral to Neuro services | 10 (0%) | 50 (0%) | 0.017 | 2.23 (1.13 -4.4) |
| | ECMO | 10 (0%) | 45 (0%) | 0.007 | 2.48 (1.25 -4.92) |
| | ECMO-VV | 10 (0%) | 15 (0%) | <0.001 | 7.44 (3.34 - 16.57) |
| | ECMO-VA | 10 (0%) | 37 (0%) | 0.001 | 3.02 (1.5 -6.07) |
| | Mortality | 2287 (2.7%) | 17740 (1.9%) | <0.001 | 1.44 (1.38 -1.5) |

For outcomes where event counts were fewer than 10, TriNetX platform suppressed exact numbers and reported '10' for de-identification purposes; thus, relative risks may be overestimated or not statistically meaningful in these cases. When zero events were observed, this is indicated as '0'. Caution is warranted in the interpretation of RRs where numerators are at or below this privacy threshold. ECMO: Extracorporeal membrane oxygenation.

**Table S8.** Outcomes in female in infected, vaccinated individuals compared to infected, unvaccinated individuals (Group 4 vs Group 2) across four post-exposure time windows.

| Time | Outcome | Group 4 (n=94,451) | Group 2 (n=1,190,308) | *p*-value | RR (95%CI) |
|---|---|---|---|---|---|
| | **Follow-up (months)** | **25.8 ± 19.1** | **24 ± 16** | | |
| 0-3 mo | Major adverse cardiovascular events | 427 (0.5%) | 8731 (0.7%) | <0.001 | 0.62 (0.56 -0.68) |
| | Cardiovascular disease | 673 (0.8%) | 13852 (1.2%) | <0.001 | 0.62 (0.58 -0.67) |
| | Ischemic heart disease | 308 (0.3%) | 5354 (0.4%) | <0.001 | 0.72 (0.64 -0.81) |
| | Myocardial angina | 52 (0.1%) | 507 (0%) | 0.087 | 1.28 (0.96 -1.71) |
| | Acute coronary disease | 59 (0.1%) | 968 (0.1%) | 0.039 | 0.76 (0.58 -0.99) |
| | Myocardial infarction | 121 (0.1%) | 2388 (0.2%) | <0.001 | 0.63 (0.53 -0.76) |
| | Ischemic cardiomyopathy | 26 (0%) | 404 (0%) | 0.273 | 0.8 (0.54 -1.19) |
| | Arrythmias | 1001 (1.2%) | 18875 (1.8%) | <0.001 | 0.7 (0.66 -0.75) |
| | Atrial fibrillation and flutter | 169 (0.2%) | 3540 (0.3%) | <0.001 | 0.59 (0.51 -0.69) |
| | Paroxysmal tachycardia | 143 (0.1%) | 2238 (0.2%) | 0.01 | 0.8 (0.68 -0.95) |
| | Other cardiac arrythmias | 370 (0.4%) | 6297 (0.5%) | <0.001 | 0.75 (0.68 -0.83) |
| | Inflammatory heart disease | 106 (0.1%) | 1900 (0.1%) | <0.001 | 0.7 (0.57 -0.85) |
| | Pericarditis | 88 (0.1%) | 1354 (0.1%) | 0.056 | 0.81 (0.65 -1.01) |
| | Myocarditis | 10 (0%) | 198 (0%) | 0.148 | 0.63 (0.33 -1.19) |
| | Endocarditis | 21 (0%) | 506 (0%) | 0.003 | 0.52 (0.33 -0.8) |
| | Hypertensive heart disease | 636 (0.9%) | 13512 (1.4%) | <0.001 | 0.64 (0.59 -0.69) |
| | Pulmonary heart diseases | 215 (0.2%) | 4313 (0.3%) | <0.001 | 0.62 (0.54 -0.71) |
| | Heart Failure | 217 (0.2%) | 4318 (0.3%) | <0.001 | 0.63 (0.55 -0.72) |
| | Cardiomyopathy | 99 (0.1%) | 1476 (0.1%) | 0.088 | 0.84 (0.68 -1.03) |
| | Cardiomegaly | 217 (0.2%) | 4654 (0.4%) | <0.001 | 0.58 (0.51 -0.67) |
| | Cardiogenic shock | 32 (0%) | 479 (0%) | 0.311 | 0.83 (0.58 -1.19) |
| | Cardiac arrest | 53 (0.1%) | 1291 (0.1%) | <0.001 | 0.51 (0.39 -0.67) |
| | Thrombotic disorders | 212 (0.2%) | 5055 (0.4%) | <0.001 | 0.52 (0.46 -0.6) |
| | Pulmonary embolism | 113 (0.1%) | 2648 (0.2%) | <0.001 | 0.53 (0.44 -0.64) |
| | Deep and superficial thrombosis | 143 (0.1%) | 3388 (0.3%) | <0.001 | 0.53 (0.45 -0.62) |
| | Thrombosis with Thrombocytopenia | 0 (0%) | 10 (0%) | 0.37 | NA |
| | Elevated Troponin I | 167 (0.2%) | 3893 (0.3%) | <0.001 | 0.53 (0.46 -0.62) |
| | Elevated Natriuretic peptide B | 324 (0.3%) | 9197 (0.7%) | <0.001 | 0.44 (0.39 -0.49) |
| | Dyslipidemia | 1192 (2.5%) | 22560 (2.7%) | 0.001 | 0.91 (0.86 -0.96) |
| | Triglyceride ≥150 mg/dl | 745 (0.9%) | 11173 (1%) | 0.079 | 0.94 (0.87 -1.01) |
| | Total cholesterol ≥200 mg/dl | 856 (1.2%) | 10559 (1%) | <0.001 | 1.22 (1.14 -1.3) |
| | LDL-cholesterol ≥130 mg/dl | 725 (0.9%) | 8392 (0.7%) | <0.001 | 1.22 (1.14 -1.32) |
| | HDL-cholesterol <50 mg/dl | 890 (1.2%) | 15201 (1.4%) | <0.001 | 0.84 (0.79 -0.9) |
| | Cerebrovascular disease | 240 (0.2%) | 4386 (0.4%) | <0.001 | 0.69 (0.6 -0.78) |
| | Transient ischemic attacks | 63 (0.1%) | 895 (0.1%) | 0.329 | 0.88 (0.68 -1.14) |
| | Cerebral infarction | 93 (0.1%) | 2050 (0.2%) | <0.001 | 0.57 (0.46 -0.7) |
| | Cerebral occlusion without infarction | 77 (0.1%) | 1357 (0.1%) | 0.003 | 0.71 (0.56 -0.89) |
| | Intracerebral hemorrhage | 26 (0%) | 421 (0%) | 0.191 | 0.77 (0.52 -1.14) |
| | Subarachnoid hemorrhage | 22 (0%) | 275 (0%) | 0.986 | 1 (0.65 -1.54) |
| | Hospital admission | 1091 (1.5%) | 26397 (3%) | <0.001 | 0.5 (0.47 -0.54) |
| | Emergency visits | 1332 (2.8%) | 22786 (4%) | <0.001 | 0.69 (0.65 -0.73) |
| | Outpatient encounters | 667 (1.8%) | 25681 (4.2%) | <0.001 | 0.42 (0.39 -0.45) |
| | Intensive care unit admission | 37 (0%) | 1946 (0.2%) | <0.001 | 0.23 (0.17 -0.32) |
| | Mechanical ventilation | 227 (0.2%) | 7467 (0.6%) | <0.001 | 0.38 (0.33 -0.43) |
| | Referral to Cardio services | 0 (0%) | 10 (0%) | 0.37 | NA |
| | Referral to Neuro services | 10 (0%) | 33 (0%) | <0.001 | 3.77 (1.86 -7.65) |
| | ECMO | 10 (0%) | 151 (0%) | 0.554 | 0.82 (0.44 -1.56) |
| | ECMO-VV | 10 (0%) | 124 (0%) | 0.991 | 1 (0.53 -1.91) |
| | ECMO-VA | 10 (0%) | 38 (0%) | <0.001 | 3.28 (1.63 -6.57) |
| | Mortality | 947 (0.9%) | 20302 (1.6%) | <0.001 | 0.58 (0.54 -0.62) |
| 3-6mo | Major adverse cardiovascular events | 315 (0.3%) | 3869 (0.3%) | 0.696 | 1.02 (0.91 -1.15) |

| | | | | | |
|---|---|---|---|---|---|
| | Cardiovascular disease | 488 (0.6%) | 5510 (0.5%) | 0.008 | 1.13 (1.03 -1.24) |
| | Ischemic heart disease | 191 (0.2%) | 2355 (0.2%) | 0.844 | 1.02 (0.88 -1.18) |
| | Myocardial angina | 25 (0%) | 353 (0%) | 0.556 | 0.89 (0.59 -1.33) |
| | Acute coronary disease | 33 (0%) | 366 (0%) | 0.526 | 1.12 (0.79 -1.6) |
| | Myocardial infarction | 56 (0.1%) | 855 (0.1%) | 0.135 | 0.81 (0.62 -1.07) |
| | Ischemic cardiomyopathy | 14 (0%) | 173 (0%) | 0.978 | 1.01 (0.59 -1.74) |
| | Arrythmias | 663 (0.8%) | 8540 (0.8%) | 0.65 | 1.02 (0.94 -1.1) |
| | Atrial fibrillation and flutter | 81 (0.1%) | 1332 (0.1%) | 0.013 | 0.75 (0.6 -0.94) |
| | Paroxysmal tachycardia | 125 (0.1%) | 1071 (0.1%) | <0.001 | 1.46 (1.22 -1.76) |
| | Other cardiac arrythmias | 254 (0.3%) | 2663 (0.2%) | 0.003 | 1.22 (1.07 -1.38) |
| | Inflammatory heart disease | 64 (0.1%) | 689 (0.1%) | 0.256 | 1.16 (0.9 -1.5) |
| | Pericarditis | 39 (0%) | 483 (0%) | 0.966 | 1.01 (0.73 -1.4) |
| | Myocarditis | 10 (0%) | 45 (0%) | 0.002 | 2.77 (1.39 -5.49) |
| | Endocarditis | 28 (0%) | 217 (0%) | 0.017 | 1.61 (1.09 -2.38) |
| | Hypertensive heart disease | 479 (0.7%) | 6748 (0.7%) | 0.364 | 0.96 (0.87 -1.05) |
| | Pulmonary heart diseases | 92 (0.1%) | 1296 (0.1%) | 0.261 | 0.89 (0.72 -1.09) |
| | Heart Failure | 113 (0.1%) | 1710 (0.1%) | 0.041 | 0.82 (0.68 -0.99) |
| | Cardiomyopathy | 58 (0.1%) | 605 (0%) | 0.189 | 1.2 (0.92 -1.57) |
| | Cardiomegaly | 121 (0.1%) | 1399 (0.1%) | 0.402 | 1.08 (0.9 -1.3) |
| | Cardiogenic shock | 14 (0%) | 152 (0%) | 0.625 | 1.15 (0.66 -1.98) |
| | Cardiac arrest | 31 (0%) | 435 (0%) | 0.514 | 0.89 (0.62 -1.28) |
| | Thrombotic disorders | 111 (0.1%) | 1423 (0.1%) | 0.782 | 0.97 (0.8 -1.18) |
| | Pulmonary embolism | 45 (0%) | 571 (0%) | 0.9 | 0.98 (0.72 -1.33) |
| | Deep and superficial thrombosis | 93 (0.1%) | 1147 (0.1%) | 0.912 | 1.01 (0.82 -1.25) |
| | Thrombosis with Thrombocytopenia | 0 (0%) | 0 (0%) | NA | NA |
| | Elevated Troponin I | 103 (0.1%) | 1226 (0.1%) | 0.692 | 1.04 (0.85 -1.27) |
| | Elevated Natriuretic peptide B | 141 (0.1%) | 1874 (0.1%) | 0.428 | 0.93 (0.79 -1.11) |
| | Dyslipidemia | 896 (1.9%) | 12049 (1.5%) | <0.001 | 1.28 (1.19 -1.37) |
| | Triglyceride ≥150 mg/dl | 550 (0.7%) | 5227 (0.5%) | <0.001 | 1.48 (1.35 -1.61) |
| | Total cholesterol ≥200 mg/dl | 699 (1%) | 7542 (0.7%) | <0.001 | 1.39 (1.29 -1.51) |
| | LDL-cholesterol ≥130 mg/dl | 610 (0.8%) | 5868 (0.5%) | <0.001 | 1.48 (1.36 -1.6) |
| | HDL-cholesterol <50 mg/dl | 575 (0.8%) | 7330 (0.7%) | 0.006 | 1.13 (1.04 -1.23) |
| | Cerebrovascular disease | 191 (0.2%) | 2187 (0.2%) | 0.222 | 1.1 (0.95 -1.27) |
| | Transient ischemic attacks | 42 (0%) | 560 (0%) | 0.69 | 0.94 (0.69 -1.28) |
| | Cerebral infarction | 87 (0.1%) | 890 (0.1%) | 0.081 | 1.22 (0.98 -1.52) |
| | Cerebral occlusion without infarction | 54 (0.1%) | 733 (0.1%) | 0.559 | 0.92 (0.7 -1.21) |
| | Intracerebral hemorrhage | 23 (0%) | 201 (0%) | 0.107 | 1.42 (0.93 -2.19) |
| | Subarachnoid hemorrhage | 14 (0%) | 115 (0%) | 0.139 | 1.52 (0.87 -2.64) |
| | Hospital admission | 716 (1%) | 11099 (1.3%) | <0.001 | 0.77 (0.72 -0.83) |
| | Emergency visits | 935 (2%) | 11309 (2.1%) | 0.276 | 0.96 (0.9 -1.03) |
| | Outpatient encounters | 384 (1%) | 12395 (2.1%) | <0.001 | 0.49 (0.44 -0.54) |
| | Intensive care unit admission | 26 (0%) | 790 (0.1%) | <0.001 | 0.4 (0.27 -0.59) |
| | Mechanical ventilation | 77 (0.1%) | 1099 (0.1%) | 0.223 | 0.87 (0.69 -1.09) |
| | Referral to Cardio services | 0 (0%) | 10 (0%) | 0.37 | NA |
| | Referral to Neuro services | 0 (0%) | 22 (0%) | 0.184 | NA |
| | ECMO | 10 (0%) | 10 (0%) | <0.001 | 12.45 (5.18 - 29.9) |
| | ECMO-VV | 0 (0%) | 10 (0%) | 0.37 | NA |
| | ECMO-VA | 10 (0%) | 10 (0%) | <0.001 | 12.45 (5.18 - 29.9) |
| | Mortality | 457 (0.4%) | 5023 (0.4%) | 0.011 | 1.13 (1.03 -1.25) |
| 6-9mo | Major adverse cardiovascular events | 269 (0.3%) | 3153 (0.3%) | 0.272 | 1.07 (0.95 -1.21) |
| | Cardiovascular disease | 394 (0.5%) | 4500 (0.4%) | 0.03 | 1.12 (1.01 -1.24) |
| | Ischemic heart disease | 157 (0.2%) | 1886 (0.2%) | 0.623 | 1.04 (0.89 -1.23) |
| | Myocardial angina | 21 (0%) | 316 (0%) | 0.41 | 0.83 (0.53 -1.29) |
| | Acute coronary disease | 25 (0%) | 229 (0%) | 0.144 | 1.36 (0.9 -2.05) |

| | | | | | |
|---|---|---|---|---|---|
| | Myocardial infarction | 60 (0.1%) | 677 (0.1%) | 0.474 | 1.1 (0.85 -1.43) |
| | Ischemic cardiomyopathy | 12 (0%) | 128 (0%) | 0.608 | 1.17 (0.65 -2.11) |
| | Arrythmias | 600 (0.7%) | 6851 (0.7%) | 0.001 | 1.15 (1.06 -1.25) |
| | Atrial fibrillation and flutter | 67 (0.1%) | 944 (0.1%) | 0.311 | 0.88 (0.69 -1.13) |
| | Paroxysmal tachycardia | 81 (0.1%) | 825 (0.1%) | 0.074 | 1.23 (0.98 -1.55) |
| | Other cardiac arrythmias | 194 (0.2%) | 2130 (0.2%) | 0.044 | 1.16 (1 -1.35) |
| | Inflammatory heart disease | 40 (0%) | 492 (0%) | 0.927 | 1.02 (0.74 -1.4) |
| | Pericarditis | 23 (0%) | 344 (0%) | 0.398 | 0.83 (0.55 -1.27) |
| | Myocarditis | 10 (0%) | 27 (0%) | <0.001 | 4.61 (2.23 -9.52) |
| | Endocarditis | 17 (0%) | 150 (0%) | 0.175 | 1.41 (0.86 -2.33) |
| | Hypertensive heart disease | 387 (0.5%) | 5585 (0.6%) | 0.2 | 0.94 (0.84 -1.04) |
| | Pulmonary heart diseases | 79 (0.1%) | 983 (0.1%) | 0.981 | 1 (0.8 -1.26) |
| | Heart Failure | 116 (0.1%) | 1223 (0.1%) | 0.093 | 1.18 (0.97 -1.42) |
| | Cardiomyopathy | 57 (0.1%) | 482 (0%) | 0.005 | 1.48 (1.12 -1.94) |
| | Cardiomegaly | 79 (0.1%) | 1188 (0.1%) | 0.114 | 0.83 (0.66 -1.05) |
| | Cardiogenic shock | 18 (0%) | 128 (0%) | 0.024 | 1.75 (1.07 -2.87) |
| | Cardiac arrest | 27 (0%) | 306 (0%) | 0.645 | 1.1 (0.74 -1.63) |
| | Thrombotic disorders | 88 (0.1%) | 1133 (0.1%) | 0.775 | 0.97 (0.78 -1.2) |
| | Pulmonary embolism | 29 (0%) | 461 (0%) | 0.2 | 0.78 (0.54 -1.14) |
| | Deep and superficial thrombosis | 80 (0.1%) | 870 (0.1%) | 0.238 | 1.15 (0.91 -1.44) |
| | Thrombosis with Thrombocytopenia | 0 (0%) | 10 (0%) | 0.37 | NA |
| | Elevated Troponin I | 85 (0.1%) | 1124 (0.1%) | 0.566 | 0.94 (0.75 -1.17) |
| | Elevated Natriuretic peptide B | 131 (0.1%) | 1466 (0.1%) | 0.26 | 1.11 (0.93 -1.33) |
| | Dyslipidemia | 791 (1.7%) | 10166 (1.3%) | <0.001 | 1.34 (1.25 -1.44) |
| | Triglyceride ≥150 mg/dl | 456 (0.6%) | 4630 (0.4%) | <0.001 | 1.39 (1.26 -1.52) |
| | Total cholesterol ≥200 mg/dl | 610 (0.9%) | 6451 (0.6%) | <0.001 | 1.43 (1.31 -1.55) |
| | LDL-cholesterol ≥130 mg/dl | 522 (0.7%) | 5200 (0.5%) | <0.001 | 1.43 (1.31 -1.56) |
| | HDL-cholesterol <50 mg/dl | 549 (0.8%) | 6501 (0.6%) | <0.001 | 1.21 (1.11 -1.32) |
| | Cerebrovascular disease | 163 (0.2%) | 1835 (0.1%) | 0.181 | 1.12 (0.95 -1.31) |
| | Transient ischemic attacks | 42 (0%) | 491 (0%) | 0.674 | 1.07 (0.78 -1.47) |
| | Cerebral infarction | 55 (0.1%) | 727 (0.1%) | 0.667 | 0.94 (0.72 -1.24) |
| | Cerebral occlusion without infarction | 51 (0%) | 596 (0%) | 0.644 | 1.07 (0.8 -1.42) |
| | Intracerebral hemorrhage | 12 (0%) | 140 (0%) | 0.831 | 1.07 (0.59 -1.92) |
| | Subarachnoid hemorrhage | 10 (0%) | 103 (0%) | 0.566 | 1.21 (0.63 -2.31) |
| | Hospital admission | 552 (0.8%) | 8473 (1%) | <0.001 | 0.78 (0.72 -0.85) |
| | Emergency visits | 846 (1.8%) | 9968 (1.9%) | 0.75 | 0.99 (0.92 -1.06) |
| | Outpatient encounters | 298 (0.8%) | 9375 (1.6%) | <0.001 | 0.5 (0.44 -0.56) |
| | Intensive care unit admission | 23 (0%) | 674 (0.1%) | <0.001 | 0.42 (0.27 -0.63) |
| | Mechanical ventilation | 76 (0.1%) | 813 (0.1%) | 0.226 | 1.16 (0.91 -1.46) |
| | Referral to Cardio services | 0 (0%) | 0 (0%) | NA | NA |
| | Referral to Neuro services | 10 (0%) | 14 (0%) | <0.001 | 8.89 (3.95 - 20.02) |
| | ECMO | 0 (0%) | 10 (0%) | 0.37 | NA |
| | ECMO-VV | 0 (0%) | 10 (0%) | 0.37 | NA |
| | ECMO-VA | 0 (0%) | 10 (0%) | 0.37 | NA |
| | Mortality | 385 (0.4%) | 3708 (0.3%) | <0.001 | 1.29 (1.16 -1.44) |
| >9mo | Major adverse cardiovascular events | 1324 (1.4%) | 15728 (1.3%) | 0.047 | 1.06 (1 -1.12) |
| | Cardiovascular disease | 1983 (2.3%) | 22230 (2%) | <0.001 | 1.14 (1.09 -1.2) |
| | Ischemic heart disease | 826 (0.8%) | 9294 (0.8%) | 0.003 | 1.11 (1.04 -1.19) |
| | Myocardial angina | 158 (0.2%) | 1484 (0.1%) | 0.001 | 1.33 (1.13 -1.57) |
| | Acute coronary disease | 90 (0.1%) | 1169 (0.1%) | 0.696 | 0.96 (0.77 -1.19) |
| | Myocardial infarction | 256 (0.2%) | 3188 (0.2%) | 0.972 | 1 (0.88 -1.13) |
| | Ischemic cardiomyopathy | 29 (0%) | 480 (0%) | 0.135 | 0.75 (0.52 -1.09) |
| | Arrythmias | 3410 (4.3%) | 38900 (3.7%) | <0.001 | 1.15 (1.11 -1.19) |
| | Atrial fibrillation and flutter | 362 (0.4%) | 4236 (0.3%) | 0.292 | 1.06 (0.95 -1.18) |
| | Paroxysmal tachycardia | 388 (0.4%) | 3761 (0.3%) | <0.001 | 1.29 (1.17 -1.44) |

| | | | | | |
|---|---|---|---|---|---|
| | Other cardiac arrythmias | 1036 (1.1%) | 10144 (0.8%) | <0.001 | 1.3 (1.22 -1.39) |
| | Inflammatory heart disease | 228 (0.2%) | 2359 (0.2%) | 0.007 | 1.21 (1.05 -1.38) |
| | Pericarditis | 134 (0.1%) | 1594 (0.1%) | 0.599 | 1.05 (0.88 -1.25) |
| | Myocarditis | 10 (0%) | 94 (0%) | 0.397 | 1.32 (0.69 -2.54) |
| | Endocarditis | 102 (0.1%) | 795 (0.1%) | <0.001 | 1.6 (1.3 -1.97) |
| | Hypertensive heart disease | 2830 (3.9%) | 34979 (3.6%) | <0.001 | 1.09 (1.05 -1.13) |
| | Pulmonary heart diseases | 378 (0.4%) | 4178 (0.3%) | 0.024 | 1.13 (1.02 -1.25) |
| | Heart Failure | 420 (0.4%) | 5316 (0.4%) | 0.7 | 0.98 (0.89 -1.08) |
| | Cardiomyopathy | 169 (0.2%) | 1874 (0.1%) | 0.137 | 1.13 (0.96 -1.32) |
| | Cardiomegaly | 496 (0.5%) | 5669 (0.4%) | 0.052 | 1.1 (1 -1.2) |
| | Cardiogenic shock | 64 (0.1%) | 607 (0%) | 0.038 | 1.31 (1.01 -1.7) |
| | Cardiac arrest | 142 (0.1%) | 1499 (0.1%) | 0.062 | 1.18 (0.99 -1.4) |
| | Thrombotic disorders | 480 (0.5%) | 5529 (0.4%) | 0.093 | 1.08 (0.99 -1.19) |
| | Pulmonary embolism | 161 (0.2%) | 1912 (0.1%) | 0.569 | 1.05 (0.89 -1.23) |
| | Deep and superficial thrombosis | 406 (0.4%) | 4566 (0.4%) | 0.044 | 1.11 (1 -1.23) |
| | Thrombosis with Thrombocytopenia | 10 (0%) | 10 (0%) | <0.001 | 12.45 (5.18 - 29.9) |
| | Elevated Troponin I | 550 (0.5%) | 7108 (0.6%) | 0.345 | 0.96 (0.88 -1.05) |
| | Elevated Natriuretic peptide B | 627 (0.6%) | 7654 (0.6%) | 0.701 | 1.02 (0.94 -1.1) |
| | Dyslipidemia | 5587 (12.3%) | 65958 (8.4%) | <0.001 | 1.47 (1.43 -1.5) |
| | Triglyceride ≥150 mg/dl | 3404 (4.3%) | 31727 (2.8%) | <0.001 | 1.51 (1.46 -1.56) |
| | Total cholesterol ≥200 mg/dl | 4371 (6.3%) | 44282 (4.2%) | <0.001 | 1.49 (1.45 -1.54) |
| | LDL-cholesterol ≥130 mg/dl | 3683 (4.7%) | 36343 (3.3%) | <0.001 | 1.44 (1.4 -1.49) |
| | HDL-cholesterol <50 mg/dl | 3848 (5.3%) | 43297 (4.2%) | <0.001 | 1.28 (1.24 -1.32) |
| | Cerebrovascular disease | 765 (0.8%) | 9146 (0.7%) | 0.19 | 1.05 (0.98 -1.13) |
| | Transient ischemic attacks | 273 (0.3%) | 2852 (0.2%) | 0.004 | 1.2 (1.06 -1.36) |
| | Cerebral infarction | 252 (0.2%) | 3420 (0.3%) | 0.184 | 0.92 (0.81 -1.04) |
| | Cerebral occlusion without infarction | 269 (0.3%) | 2860 (0.2%) | 0.011 | 1.18 (1.04 -1.33) |
| | Intracerebral hemorrhage | 51 (0%) | 651 (0%) | 0.86 | 0.98 (0.73 -1.3) |
| | Subarachnoid hemorrhage | 49 (0%) | 482 (0%) | 0.115 | 1.27 (0.94 -1.7) |
| | Hospital admission | 3310 (4.8%) | 47086 (5.7%) | <0.001 | 0.84 (0.81 -0.87) |
| | Emergency visits | 5123 (11.4%) | 58839 (11.2%) | 0.306 | 1.01 (0.99 -1.04) |
| | Outpatient encounters | 1462 (4%) | 51230 (9.1%) | <0.001 | 0.44 (0.42 -0.47) |
| | Intensive care unit admission | 67 (0.1%) | 2605 (0.2%) | <0.001 | 0.31 (0.25 -0.4) |
| | Mechanical ventilation | 326 (0.3%) | 4069 (0.3%) | 0.872 | 0.99 (0.89 -1.11) |
| | Referral to Cardio services | 0 (0%) | 10 (0%) | 0.37 | NA |
| | Referral to Neuro services | 10 (0%) | 68 (0%) | 0.07 | 1.83 (0.94 -3.56) |
| | ECMO | 10 (0%) | 28 (0%) | <0.001 | 4.45 (2.16 -9.15) |
| | ECMO-VV | 10 (0%) | 12 (0%) | <0.001 | 10.37 (4.48 - 24.01) |
| | ECMO-VA | 10 (0%) | 19 (0%) | <0.001 | 6.55 (3.05 - 14.09) |
| | Mortality | 1900 (1.8%) | 18241 (1.4%) | <0.001 | 1.3 (1.24 -1.36) |

For outcomes where event counts were fewer than 10, TriNetX platform suppressed exact numbers and reported '10' for de-identification purposes; thus, relative risks may be overestimated or not statistically meaningful in these cases. When zero events were observed, this is indicated as '0'. Caution is warranted in the interpretation of RRs where numerators are at or below this privacy threshold. ECMO: Extracorporeal membrane oxygenation.

**Table S9.** Outcomes in male vaccinated individuals without infection compared to infected, unvaccinated individuals (Group 3 vs Group 2) across four post-exposure time windows.

| Time | Outcome | Group 3 (n=625,068) | Group 2 (n=848,176) | *p*-value | RR (95%CI) |
|---|---|---|---|---|---|
| | **Follow-up (months)** | **22.7 ± 18.8** | **22 ± 16.2** | | |
| 0-3 mo | Major adverse cardiovascular events | 1693 (0.3%) | 9663 (1.1%) | <0.001 | 0.24 (0.23 -0.25) |
| | Cardiovascular disease | 2209 (0.4%) | 14202 (1.8%) | <0.001 | 0.21 (0.2 -0.22) |
| | Ischemic heart disease | 1200 (0.2%) | 6710 (0.8%) | <0.001 | 0.25 (0.23 -0.26) |
| | Myocardial angina | 184 (0%) | 610 (0.1%) | <0.001 | 0.44 (0.37 -0.51) |
| | Acute coronary disease | 129 (0%) | 1222 (0.1%) | <0.001 | 0.15 (0.13 -0.18) |
| | Myocardial infarction | 346 (0.1%) | 3121 (0.3%) | <0.001 | 0.16 (0.14 -0.18) |
| | Ischemic cardiomyopathy | 163 (0%) | 926 (0.1%) | <0.001 | 0.25 (0.22 -0.3) |
| | Arrythmias | 2507 (0.4%) | 15915 (2%) | <0.001 | 0.21 (0.2 -0.22) |
| | Atrial fibrillation and flutter | 654 (0.1%) | 4822 (0.5%) | <0.001 | 0.19 (0.18 -0.21) |
| | Paroxysmal tachycardia | 410 (0.1%) | 2557 (0.3%) | <0.001 | 0.23 (0.21 -0.26) |
| | Other cardiac arrythmias | 1001 (0.2%) | 6379 (0.7%) | <0.001 | 0.22 (0.21 -0.24) |
| | Inflammatory heart disease | 256 (0%) | 2327 (0.2%) | <0.001 | 0.16 (0.14 -0.18) |
| | Pericarditis | 171 (0%) | 1495 (0.2%) | <0.001 | 0.17 (0.14 -0.19) |
| | Myocarditis | 14 (0%) | 279 (0%) | <0.001 | 0.07 (0.04 -0.12) |
| | Endocarditis | 85 (0%) | 752 (0.1%) | <0.001 | 0.16 (0.13 -0.21) |
| | Hypertensive heart disease | 3148 (0.6%) | 12479 (1.8%) | <0.001 | 0.33 (0.31 -0.34) |
| | Pulmonary heart diseases | 453 (0.1%) | 5033 (0.5%) | <0.001 | 0.13 (0.12 -0.14) |
| | Heart Failure | 656 (0.1%) | 4924 (0.5%) | <0.001 | 0.19 (0.17 -0.2) |
| | Cardiomyopathy | 365 (0.1%) | 2362 (0.2%) | <0.001 | 0.22 (0.2 -0.25) |
| | Cardiomegaly | 584 (0.1%) | 5574 (0.6%) | <0.001 | 0.15 (0.14 -0.16) |
| | Cardiogenic shock | 73 (0%) | 752 (0.1%) | <0.001 | 0.14 (0.11 -0.18) |
| | Cardiac arrest | 159 (0%) | 1832 (0.2%) | <0.001 | 0.13 (0.11 -0.15) |
| | Thrombotic disorders | 416 (0.1%) | 6092 (0.6%) | <0.001 | 0.1 (0.09 -0.11) |
| | Pulmonary embolism | 176 (0%) | 3384 (0.4%) | <0.001 | 0.08 (0.06 -0.09) |
| | Deep and superficial thrombosis | 328 (0%) | 3921 (0.4%) | <0.001 | 0.12 (0.11 -0.13) |
| | Thrombosis with Thrombocytopenia | 0 (0%) | 0 (0%) | NA | NA |
| | Elevated Troponin I | 258 (0%) | 4359 (0.5%) | <0.001 | 0.09 (0.08 -0.1) |
| | Elevated Natriuretic peptide B | 757 (0.1%) | 10193 (1.1%) | <0.001 | 0.1 (0.1 -0.11) |
| | Dyslipidemia | 5897 (1.2%) | 21348 (3.5%) | <0.001 | 0.35 (0.34 -0.36) |
| | Triglyceride ≥150 mg/dl | 2815 (0.5%) | 12159 (1.5%) | <0.001 | 0.33 (0.31 -0.34) |
| | Total cholesterol ≥200 mg/dl | 2640 (0.4%) | 6476 (0.8%) | <0.001 | 0.58 (0.55 -0.6) |
| | LDL-cholesterol ≥130 mg/dl | 2325 (0.4%) | 5758 (0.7%) | <0.001 | 0.57 (0.55 -0.6) |
| | HDL-cholesterol <50 mg/dl | 4787 (0.9%) | 18373 (2.5%) | <0.001 | 0.35 (0.34 -0.36) |
| | Cerebrovascular disease | 825 (0.1%) | 4289 (0.5%) | <0.001 | 0.27 (0.25 -0.29) |
| | Transient ischemic attacks | 176 (0%) | 771 (0.1%) | <0.001 | 0.33 (0.28 -0.39) |
| | Cerebral infarction | 316 (0%) | 2146 (0.2%) | <0.001 | 0.21 (0.19 -0.24) |
| | Cerebral occlusion without infarction | 305 (0%) | 1543 (0.2%) | <0.001 | 0.29 (0.25 -0.32) |
| | Intracerebral hemorrhage | 81 (0%) | 585 (0.1%) | <0.001 | 0.2 (0.16 -0.25) |
| | Subarachnoid hemorrhage | 46 (0%) | 329 (0%) | <0.001 | 0.2 (0.15 -0.28) |
| | Hospital admission | 2427 (0.4%) | 19728 (2.9%) | <0.001 | 0.15 (0.14 -0.15) |
| | Emergency visits | 5000 (1%) | 18523 (4.5%) | <0.001 | 0.24 (0.23 -0.24) |
| | Outpatient encounters | 7499 (1.9%) | 23366 (4.8%) | <0.001 | 0.4 (0.39 -0.41) |
| | Intensive care unit admission | 142 (0%) | 1744 (0.2%) | <0.001 | 0.12 (0.1 -0.14) |
| | Mechanical ventilation | 339 (0.1%) | 10711 (1.1%) | <0.001 | 0.05 (0.04 -0.05) |
| | Referral to Cardio services | 0 (0%) | 10 (0%) | 0.009 | NA |
| | Referral to Neuro services | 10 (0%) | 26 (0%) | 0.112 | 0.56 (0.27 -1.16) |
| | ECMO | 10 (0%) | 289 (0%) | <0.001 | 0.05 (0.03 -0.09) |
| | ECMO-VV | 10 (0%) | 240 (0%) | <0.001 | 0.06 (0.03 -0.11) |
| | ECMO-VA | 10 (0%) | 98 (0%) | <0.001 | 0.15 (0.08 -0.28) |
| | Mortality | 1380 (0.2%) | 25841 (2.7%) | <0.001 | 0.08 (0.07 -0.08) |
| 3-6mo | Major adverse cardiovascular events | 1418 (0.2%) | 3424 (0.4%) | <0.001 | 0.56 (0.52 -0.59) |

| | | | | | |
|---|---|---|---|---|---|
| | Cardiovascular disease | 1752 (0.3%) | 4189 (0.5%) | <0.001 | 0.55 (0.52 -0.58) |
| | Ischemic heart disease | 996 (0.2%) | 2392 (0.3%) | <0.001 | 0.58 (0.53 -0.62) |
| | Myocardial angina | 153 (0%) | 337 (0%) | <0.001 | 0.66 (0.54 -0.79) |
| | Acute coronary disease | 121 (0%) | 386 (0%) | <0.001 | 0.45 (0.37 -0.55) |
| | Myocardial infarction | 300 (0%) | 961 (0.1%) | <0.001 | 0.45 (0.39 -0.51) |
| | Ischemic cardiomyopathy | 116 (0%) | 323 (0%) | <0.001 | 0.52 (0.42 -0.64) |
| | Arrythmias | 2097 (0.3%) | 5337 (0.7%) | <0.001 | 0.51 (0.49 -0.54) |
| | Atrial fibrillation and flutter | 538 (0.1%) | 1375 (0.1%) | <0.001 | 0.55 (0.5 -0.61) |
| | Paroxysmal tachycardia | 346 (0.1%) | 825 (0.1%) | <0.001 | 0.6 (0.53 -0.68) |
| | Other cardiac arrythmias | 810 (0.1%) | 1995 (0.2%) | <0.001 | 0.57 (0.53 -0.62) |
| | Inflammatory heart disease | 215 (0%) | 632 (0.1%) | <0.001 | 0.49 (0.42 -0.57) |
| | Pericarditis | 142 (0%) | 418 (0%) | <0.001 | 0.49 (0.41 -0.59) |
| | Myocarditis | 10 (0%) | 46 (0%) | <0.001 | 0.32 (0.16 -0.62) |
| | Endocarditis | 73 (0%) | 224 (0%) | <0.001 | 0.47 (0.36 -0.61) |
| | Hypertensive heart disease | 2354 (0.4%) | 5242 (0.8%) | <0.001 | 0.57 (0.55 -0.6) |
| | Pulmonary heart diseases | 402 (0.1%) | 1097 (0.1%) | <0.001 | 0.52 (0.47 -0.59) |
| | Heart Failure | 558 (0.1%) | 1593 (0.2%) | <0.001 | 0.49 (0.45 -0.54) |
| | Cardiomyopathy | 290 (0%) | 750 (0.1%) | <0.001 | 0.56 (0.48 -0.64) |
| | Cardiomegaly | 449 (0.1%) | 1329 (0.1%) | <0.001 | 0.48 (0.43 -0.54) |
| | Cardiogenic shock | 79 (0%) | 236 (0%) | <0.001 | 0.48 (0.38 -0.63) |
| | Cardiac arrest | 136 (0%) | 557 (0.1%) | <0.001 | 0.35 (0.29 -0.43) |
| | Thrombotic disorders | 369 (0.1%) | 1121 (0.1%) | <0.001 | 0.47 (0.42 -0.53) |
| | Pulmonary embolism | 156 (0%) | 489 (0.1%) | <0.001 | 0.46 (0.38 -0.55) |
| | Deep and superficial thrombosis | 279 (0%) | 891 (0.1%) | <0.001 | 0.45 (0.39 -0.51) |
| | Thrombosis with Thrombocytopenia | 0 (0%) | 0 (0%) | NA | NA |
| | Elevated Troponin I | 249 (0%) | 1134 (0.1%) | <0.001 | 0.31 (0.27 -0.36) |
| | Elevated Natriuretic peptide B | 670 (0.1%) | 1627 (0.2%) | <0.001 | 0.57 (0.52 -0.63) |
| | Dyslipidemia | 4036 (0.9%) | 8318 (1.4%) | <0.001 | 0.61 (0.59 -0.63) |
| | Triglyceride ≥150 mg/dl | 2004 (0.3%) | 4393 (0.5%) | <0.001 | 0.64 (0.6 -0.67) |
| | Total cholesterol ≥200 mg/dl | 1847 (0.3%) | 4080 (0.5%) | <0.001 | 0.64 (0.6 -0.67) |
| | LDL-cholesterol ≥130 mg/dl | 1603 (0.3%) | 3591 (0.4%) | <0.001 | 0.63 (0.6 -0.67) |
| | HDL-cholesterol <50 mg/dl | 3199 (0.6%) | 7006 (1%) | <0.001 | 0.61 (0.58 -0.63) |
| | Cerebrovascular disease | 681 (0.1%) | 1711 (0.2%) | <0.001 | 0.56 (0.51 -0.61) |
| | Transient ischemic attacks | 158 (0%) | 414 (0%) | <0.001 | 0.55 (0.46 -0.66) |
| | Cerebral infarction | 275 (0%) | 732 (0.1%) | <0.001 | 0.54 (0.47 -0.62) |
| | Cerebral occlusion without infarction | 247 (0%) | 603 (0.1%) | <0.001 | 0.59 (0.51 -0.68) |
| | Intracerebral hemorrhage | 65 (0%) | 177 (0%) | <0.001 | 0.53 (0.4 -0.71) |
| | Subarachnoid hemorrhage | 47 (0%) | 109 (0%) | 0.007 | 0.63 (0.44 -0.88) |
| | Hospital admission | 1974 (0.3%) | 5600 (0.8%) | <0.001 | 0.41 (0.39 -0.43) |
| | Emergency visits | 4724 (1%) | 8228 (2.1%) | <0.001 | 0.48 (0.47 -0.5) |
| | Outpatient encounters | 5624 (1.5%) | 9303 (2%) | <0.001 | 0.73 (0.71 -0.75) |
| | Intensive care unit admission | 80 (0%) | 516 (0.1%) | <0.001 | 0.22 (0.17 -0.28) |
| | Mechanical ventilation | 296 (0%) | 1084 (0.1%) | <0.001 | 0.39 (0.34 -0.44) |
| | Referral to Cardio services | 0 (0%) | 0 (0%) | NA | NA |
| | Referral to Neuro services | 10 (0%) | 13 (0%) | 0.794 | 1.12 (0.49 -2.55) |
| | ECMO | 10 (0%) | 11 (0%) | 0.526 | 1.32 (0.56 -3.11) |
| | ECMO-VV | 10 (0%) | 10 (0%) | 0.403 | 1.45 (0.6 -3.49) |
| | ECMO-VA | 10 (0%) | 10 (0%) | 0.403 | 1.45 (0.6 -3.49) |
| | Mortality | 1531 (0.2%) | 5285 (0.5%) | <0.001 | 0.42 (0.4 -0.45) |
| 6-9mo | Major adverse cardiovascular events | 1319 (0.2%) | 2768 (0.3%) | <0.001 | 0.64 (0.6 -0.68) |
| | Cardiovascular disease | 1638 (0.3%) | 3432 (0.4%) | <0.001 | 0.62 (0.59 -0.66) |
| | Ischemic heart disease | 924 (0.1%) | 1875 (0.2%) | <0.001 | 0.68 (0.63 -0.74) |
| | Myocardial angina | 157 (0%) | 295 (0%) | 0.008 | 0.77 (0.63 -0.93) |
| | Acute coronary disease | 115 (0%) | 291 (0%) | <0.001 | 0.57 (0.46 -0.71) |
| | Myocardial infarction | 309 (0%) | 687 (0.1%) | <0.001 | 0.64 (0.56 -0.73) |
| | Ischemic cardiomyopathy | 132 (0%) | 249 (0%) | 0.013 | 0.77 (0.62 -0.95) |

| | | | | | |
|---|---|---|---|---|---|
| | Arrythmias | 1959 (0.3%) | 4593 (0.6%) | <0.001 | 0.55 (0.52 -0.58) |
| | Atrial fibrillation and flutter | 561 (0.1%) | 1074 (0.1%) | <0.001 | 0.73 (0.66 -0.81) |
| | Paroxysmal tachycardia | 304 (0%) | 659 (0.1%) | <0.001 | 0.66 (0.58 -0.76) |
| | Other cardiac arrythmias | 739 (0.1%) | 1649 (0.2%) | <0.001 | 0.63 (0.58 -0.69) |
| | Inflammatory heart disease | 185 (0%) | 420 (0%) | <0.001 | 0.63 (0.53 -0.75) |
| | Pericarditis | 131 (0%) | 273 (0%) | 0.001 | 0.69 (0.56 -0.85) |
| | Myocarditis | 10 (0%) | 33 (0%) | 0.019 | 0.44 (0.22 -0.89) |
| | Endocarditis | 56 (0%) | 155 (0%) | <0.001 | 0.52 (0.39 -0.71) |
| | Hypertensive heart disease | 2270 (0.4%) | 4322 (0.6%) | <0.001 | 0.67 (0.64 -0.7) |
| | Pulmonary heart diseases | 367 (0.1%) | 777 (0.1%) | <0.001 | 0.67 (0.59 -0.76) |
| | Heart Failure | 547 (0.1%) | 1223 (0.1%) | <0.001 | 0.63 (0.57 -0.69) |
| | Cardiomyopathy | 301 (0%) | 588 (0.1%) | <0.001 | 0.73 (0.64 -0.84) |
| | Cardiomegaly | 418 (0.1%) | 974 (0.1%) | <0.001 | 0.61 (0.54 -0.68) |
| | Cardiogenic shock | 97 (0%) | 181 (0%) | 0.043 | 0.78 (0.61 -0.99) |
| | Cardiac arrest | 175 (0%) | 346 (0%) | 0.001 | 0.73 (0.61 -0.88) |
| | Thrombotic disorders | 363 (0.1%) | 872 (0.1%) | <0.001 | 0.59 (0.52 -0.67) |
| | Pulmonary embolism | 141 (0%) | 366 (0%) | <0.001 | 0.55 (0.46 -0.67) |
| | Deep and superficial thrombosis | 284 (0%) | 692 (0.1%) | <0.001 | 0.59 (0.51 -0.67) |
| | Thrombosis with Thrombocytopenia | 0 (0%) | 0 (0%) | NA | NA |
| | Elevated Troponin I | 241 (0%) | 952 (0.1%) | <0.001 | 0.36 (0.31 -0.42) |
| | Elevated Natriuretic peptide B | 608 (0.1%) | 1272 (0.1%) | <0.001 | 0.66 (0.6 -0.73) |
| | Dyslipidemia | 3559 (0.8%) | 7003 (1.2%) | <0.001 | 0.63 (0.61 -0.66) |
| | Triglyceride ≥150 mg/dl | 1894 (0.3%) | 3841 (0.5%) | <0.001 | 0.69 (0.65 -0.72) |
| | Total cholesterol ≥200 mg/dl | 1821 (0.3%) | 3670 (0.4%) | <0.001 | 0.7 (0.66 -0.74) |
| | LDL-cholesterol ≥130 mg/dl | 1663 (0.3%) | 3135 (0.4%) | <0.001 | 0.75 (0.71 -0.8) |
| | HDL-cholesterol <50 mg/dl | 2852 (0.5%) | 6017 (0.9%) | <0.001 | 0.63 (0.6 -0.66) |
| | Cerebrovascular disease | 653 (0.1%) | 1440 (0.2%) | <0.001 | 0.64 (0.58 -0.7) |
| | Transient ischemic attacks | 172 (0%) | 373 (0%) | <0.001 | 0.67 (0.56 -0.8) |
| | Cerebral infarction | 312 (0%) | 629 (0.1%) | <0.001 | 0.71 (0.62 -0.81) |
| | Cerebral occlusion without infarction | 252 (0%) | 546 (0.1%) | <0.001 | 0.66 (0.57 -0.77) |
| | Intracerebral hemorrhage | 77 (0%) | 152 (0%) | 0.026 | 0.73 (0.56 -0.96) |
| | Subarachnoid hemorrhage | 40 (0%) | 101 (0%) | 0.003 | 0.57 (0.4 -0.83) |
| | Hospital admission | 1867 (0.3%) | 4900 (0.7%) | <0.001 | 0.44 (0.42 -0.46) |
| | Emergency visits | 4311 (0.9%) | 7267 (1.9%) | <0.001 | 0.49 (0.48 -0.51) |
| | Outpatient encounters | 4810 (1.3%) | 7234 (1.6%) | <0.001 | 0.8 (0.77 -0.83) |
| | Intensive care unit admission | 71 (0%) | 430 (0%) | <0.001 | 0.23 (0.18 -0.3) |
| | Mechanical ventilation | 328 (0%) | 835 (0.1%) | <0.001 | 0.55 (0.49 -0.63) |
| | Referral to Cardio services | 0 (0%) | 0 (0%) | NA | NA |
| | Referral to Neuro services | 10 (0%) | 10 (0%) | 0.402 | 1.45 (0.6 -3.49) |
| | ECMO | 10 (0%) | 10 (0%) | 0.403 | 1.45 (0.6 -3.48) |
| | ECMO-VV | 10 (0%) | 10 (0%) | 0.403 | 1.45 (0.6 -3.49) |
| | ECMO-VA | 10 (0%) | 10 (0%) | 0.403 | 1.45 (0.6 -3.49) |
| | Mortality | 1689 (0.3%) | 3926 (0.4%) | <0.001 | 0.62 (0.59 -0.66) |
| >9mo | Major adverse cardiovascular events | 10036 (1.6%) | 14065 (1.7%) | 0.001 | 0.96 (0.93 -0.98) |
| | Cardiovascular disease | 13397 (2.2%) | 17599 (2.2%) | 0.501 | 0.99 (0.97 -1.02) |
| | Ischemic heart disease | 6787 (1.1%) | 9374 (1.1%) | 0.905 | 1 (0.97 -1.03) |
| | Myocardial angina | 944 (0.1%) | 1388 (0.1%) | 0.676 | 0.98 (0.91 -1.07) |
| | Acute coronary disease | 784 (0.1%) | 1205 (0.1%) | 0.158 | 0.94 (0.86 -1.03) |
| | Myocardial infarction | 2133 (0.3%) | 3323 (0.4%) | 0.001 | 0.92 (0.87 -0.97) |
| | Ischemic cardiomyopathy | 749 (0.1%) | 983 (0.1%) | 0.048 | 1.1 (1 -1.21) |
| | Arrythmias | 15734 (2.6%) | 23914 (3%) | <0.001 | 0.85 (0.83 -0.87) |
| | Atrial fibrillation and flutter | 3746 (0.6%) | 4846 (0.5%) | <0.001 | 1.09 (1.04 -1.13) |
| | Paroxysmal tachycardia | 2194 (0.3%) | 2823 (0.3%) | <0.001 | 1.12 (1.06 -1.18) |
| | Other cardiac arrythmias | 5736 (0.9%) | 7831 (0.9%) | 0.082 | 1.03 (1 -1.07) |
| | Inflammatory heart disease | 1276 (0.2%) | 1949 (0.2%) | 0.095 | 0.94 (0.88 -1.01) |
| | Pericarditis | 815 (0.1%) | 1275 (0.1%) | 0.071 | 0.92 (0.85 -1.01) |

| | | | | | |
|---|---|---|---|---|---|
| | Myocarditis | 40 (0%) | 127 (0%) | <0.001 | 0.46 (0.32 -0.65) |
| | Endocarditis | 491 (0.1%) | 719 (0.1%) | 0.841 | 0.99 (0.88 -1.11) |
| | Hypertensive heart disease | 21618 (4%) | 27983 (4.1%) | 0.04 | 0.98 (0.97 -1) |
| | Pulmonary heart diseases | 2430 (0.4%) | 3387 (0.4%) | 0.435 | 1.02 (0.97 -1.08) |
| | Heart Failure | 3848 (0.6%) | 5371 (0.6%) | 0.805 | 1.01 (0.97 -1.05) |
| | Cardiomyopathy | 1888 (0.3%) | 2448 (0.3%) | 0.001 | 1.11 (1.04 -1.17) |
| | Cardiomegaly | 2859 (0.4%) | 4900 (0.5%) | <0.001 | 0.83 (0.79 -0.87) |
| | Cardiogenic shock | 557 (0.1%) | 791 (0.1%) | 0.734 | 1.02 (0.91 -1.14) |
| | Cardiac arrest | 1285 (0.2%) | 2173 (0.2%) | <0.001 | 0.85 (0.8 -0.91) |
| | Thrombotic disorders | 2714 (0.4%) | 4410 (0.5%) | <0.001 | 0.87 (0.83 -0.92) |
| | Pulmonary embolism | 1033 (0.2%) | 1570 (0.2%) | 0.147 | 0.94 (0.87 -1.02) |
| | Deep and superficial thrombosis | 2180 (0.3%) | 3672 (0.4%) | <0.001 | 0.85 (0.8 -0.89) |
| | Thrombosis with Thrombocytopenia | 10 (0%) | 0 (0%) | <0.001 | NA |
| | Elevated Troponin I | 2953 (0.4%) | 6176 (0.7%) | <0.001 | 0.68 (0.65 -0.71) |
| | Elevated Natriuretic peptide B | 4366 (0.7%) | 6156 (0.7%) | 0.427 | 0.98 (0.95 -1.02) |
| | Dyslipidemia | 33446 (7.3%) | 44346 (7.8%) | <0.001 | 0.94 (0.92 -0.95) |
| | Triglyceride ≥150 mg/dl | 17810 (3.1%) | 25611 (3.2%) | <0.001 | 0.97 (0.95 -0.98) |
| | Total cholesterol ≥200 mg/dl | 17202 (2.9%) | 24812 (3%) | 0.008 | 0.98 (0.96 -0.99) |
| | LDL-cholesterol ≥130 mg/dl | 15393 (2.6%) | 22463 (2.7%) | 0.001 | 0.97 (0.95 -0.99) |
| | HDL-cholesterol <50 mg/dl | 25289 (4.8%) | 37975 (5.5%) | <0.001 | 0.88 (0.87 -0.89) |
| | Cerebrovascular disease | 4759 (0.7%) | 6715 (0.7%) | 0.874 | 1 (0.96 -1.04) |
| | Transient ischemic attacks | 1206 (0.2%) | 1850 (0.2%) | 0.096 | 0.94 (0.88 -1.01) |
| | Cerebral infarction | 1884 (0.3%) | 2677 (0.3%) | 0.817 | 1.01 (0.95 -1.07) |
| | Cerebral occlusion without infarction | 1642 (0.2%) | 2383 (0.3%) | 0.763 | 0.99 (0.93 -1.06) |
| | Intracerebral hemorrhage | 489 (0.1%) | 608 (0.1%) | 0.012 | 1.16 (1.03 -1.31) |
| | Subarachnoid hemorrhage | 368 (0.1%) | 429 (0%) | 0.002 | 1.24 (1.08 -1.43) |
| | Hospital admission | 16173 (2.8%) | 32255 (4.9%) | <0.001 | 0.58 (0.56 -0.59) |
| | Emergency visits | 37742 (8.1%) | 43510 (11.4%) | <0.001 | 0.71 (0.7 -0.72) |
| | Outpatient encounters | 34724 (9.4%) | 39487 (8.9%) | <0.001 | 1.05 (1.04 -1.07) |
| | Intensive care unit admission | 321 (0%) | 1747 (0.2%) | <0.001 | 0.26 (0.23 -0.29) |
| | Mechanical ventilation | 2433 (0.4%) | 4219 (0.4%) | <0.001 | 0.81 (0.77 -0.86) |
| | Referral to Cardio services | 0 (0%) | 0 (0%) | NA | NA |
| | Referral to Neuro services | 27 (0%) | 56 (0%) | 0.125 | 0.7 (0.44 -1.11) |
| | ECMO | 21 (0%) | 44 (0%) | 0.163 | 0.69 (0.41 -1.16) |
| | ECMO-VV | 10 (0%) | 14 (0%) | 0.932 | 1.04 (0.46 -2.33) |
| | ECMO-VA | 16 (0%) | 35 (0%) | 0.171 | 0.66 (0.37 -1.2) |
| | Mortality | 14030 (2.1%) | 18316 (1.9%) | <0.001 | 1.11 (1.09 -1.14) |

For outcomes where event counts were fewer than 10, TriNetX platform suppressed exact numbers and reported '10' for de-identification purposes; thus, relative risks may be overestimated or not statistically meaningful in these cases. When zero events were observed, this is indicated as '0'. Caution is warranted in the interpretation of RRs where numerators are at or below this privacy threshold. ECMO: Extracorporeal membrane oxygenation.

**Table S10.** Outcomes in female vaccinated individuals without infection compared to infected, unvaccinated individuals (Group 3 vs Group 2) across four post-exposure time windows.

| Time | Outcome | Group 3 (n=751,081) | Group 2 (n=1,184,987) | *p*-value | RR (95%CI) |
|---|---|---|---|---|---|
| | **Follow-up (months)** | **25.8 ± 19.1** | **24 ± 16** | | |
| 0-3 mo | Major adverse cardiovascular events | 1760 (0.2%) | 8728 (0.7%) | <0.001 | 0.32 (0.3 -0.34) |
| | Cardiovascular disease | 2451 (0.3%) | 13908 (1.2%) | <0.001 | 0.27 (0.26 -0.29) |
| | Ischemic heart disease | 1064 (0.1%) | 5353 (0.4%) | <0.001 | 0.32 (0.3 -0.34) |
| | Myocardial angina | 164 (0%) | 517 (0%) | <0.001 | 0.52 (0.44 -0.63) |
| | Acute coronary disease | 132 (0%) | 962 (0.1%) | <0.001 | 0.23 (0.19 -0.27) |
| | Myocardial infarction | 313 (0%) | 2377 (0.2%) | <0.001 | 0.22 (0.19 -0.24) |
| | Ischemic cardiomyopathy | 72 (0%) | 401 (0%) | <0.001 | 0.3 (0.23 -0.38) |
| | Arrythmias | 3438 (0.5%) | 18986 (1.7%) | <0.001 | 0.27 (0.26 -0.28) |
| | Atrial fibrillation and flutter | 521 (0.1%) | 3540 (0.3%) | <0.001 | 0.24 (0.22 -0.26) |
| | Paroxysmal tachycardia | 477 (0.1%) | 2226 (0.2%) | <0.001 | 0.35 (0.32 -0.39) |
| | Other cardiac arrythmias | 1079 (0.1%) | 6315 (0.5%) | <0.001 | 0.28 (0.26 -0.3) |
| | Inflammatory heart disease | 256 (0%) | 1916 (0.1%) | <0.001 | 0.22 (0.19 -0.25) |
| | Pericarditis | 184 (0%) | 1362 (0.1%) | <0.001 | 0.22 (0.19 -0.26) |
| | Myocarditis | 10 (0%) | 199 (0%) | <0.001 | 0.08 (0.04 -0.16) |
| | Endocarditis | 79 (0%) | 511 (0%) | <0.001 | 0.26 (0.2 -0.32) |
| | Hypertensive heart disease | 3337 (0.5%) | 13577 (1.4%) | <0.001 | 0.37 (0.36 -0.39) |
| | Pulmonary heart diseases | 534 (0.1%) | 4287 (0.3%) | <0.001 | 0.2 (0.19 -0.22) |
| | Heart Failure | 660 (0.1%) | 4313 (0.3%) | <0.001 | 0.25 (0.23 -0.27) |
| | Cardiomyopathy | 272 (0%) | 1466 (0.1%) | <0.001 | 0.31 (0.27 -0.35) |
| | Cardiomegaly | 570 (0.1%) | 4713 (0.4%) | <0.001 | 0.2 (0.18 -0.22) |
| | Cardiogenic shock | 56 (0%) | 474 (0%) | <0.001 | 0.2 (0.15 -0.26) |
| | Cardiac arrest | 107 (0%) | 1270 (0.1%) | <0.001 | 0.14 (0.11 -0.17) |
| | Thrombotic disorders | 501 (0.1%) | 5047 (0.4%) | <0.001 | 0.16 (0.15 -0.18) |
| | Pulmonary embolism | 211 (0%) | 2645 (0.2%) | <0.001 | 0.13 (0.11 -0.15) |
| | Deep and superficial thrombosis | 384 (0%) | 3372 (0.3%) | <0.001 | 0.19 (0.17 -0.21) |
| | Thrombosis with Thrombocytopenia | 0 (0%) | 10 (0%) | 0.014 | NA |
| | Elevated Troponin I | 262 (0%) | 3806 (0.3%) | <0.001 | 0.11 (0.1 -0.13) |
| | Elevated Natriuretic peptide B | 797 (0.1%) | 9123 (0.7%) | <0.001 | 0.14 (0.13 -0.15) |
| | Dyslipidemia | 6548 (1.1%) | 22612 (2.7%) | <0.001 | 0.42 (0.41 -0.43) |
| | Triglyceride ≥150 mg/dl | 2604 (0.4%) | 11164 (1%) | <0.001 | 0.38 (0.36 -0.39) |
| | Total cholesterol ≥200 mg/dl | 3831 (0.6%) | 10593 (1%) | <0.001 | 0.57 (0.55 -0.59) |
| | LDL-cholesterol ≥130 mg/dl | 2933 (0.4%) | 8378 (0.7%) | <0.001 | 0.56 (0.54 -0.59) |
| | HDL-cholesterol <50 mg/dl | 3682 (0.5%) | 15215 (1.4%) | <0.001 | 0.38 (0.36 -0.39) |
| | Cerebrovascular disease | 964 (0.1%) | 4353 (0.3%) | <0.001 | 0.36 (0.34 -0.39) |
| | Transient ischemic attacks | 257 (0%) | 890 (0.1%) | <0.001 | 0.48 (0.41 -0.55) |
| | Cerebral infarction | 356 (0%) | 2040 (0.2%) | <0.001 | 0.29 (0.26 -0.32) |
| | Cerebral occlusion without infarction | 331 (0%) | 1357 (0.1%) | <0.001 | 0.4 (0.36 -0.45) |
| | Intracerebral hemorrhage | 71 (0%) | 418 (0%) | <0.001 | 0.28 (0.22 -0.36) |
| | Subarachnoid hemorrhage | 57 (0%) | 269 (0%) | <0.001 | 0.35 (0.26 -0.47) |
| | Hospital admission | 4168 (0.6%) | 27691 (3.2%) | <0.001 | 0.2 (0.2 -0.21) |
| | Emergency visits | 6483 (1.1%) | 23033 (4%) | <0.001 | 0.29 (0.28 -0.3) |
| | Outpatient encounters | 9139 (2.1%) | 25651 (4.2%) | <0.001 | 0.49 (0.48 -0.5) |
| | Intensive care unit admission | 143 (0%) | 1948 (0.2%) | <0.001 | 0.12 (0.1 -0.14) |
| | Mechanical ventilation | 271 (0%) | 7376 (0.6%) | <0.001 | 0.06 (0.05 -0.07) |
| | Referral to Cardio services | 0 (0%) | 10 (0%) | 0.014 | NA |
| | Referral to Neuro services | 10 (0%) | 33 (0%) | 0.051 | 0.5 (0.25 -1.02) |
| | ECMO | 10 (0%) | 151 (0%) | <0.001 | 0.11 (0.06 -0.21) |
| | ECMO-VV | 0 (0%) | 124 (0%) | <0.001 | NA |
| | ECMO-VA | 10 (0%) | 38 (0%) | 0.016 | 0.44 (0.22 -0.88) |
| | Mortality | 1157 (0.1%) | 20157 (1.5%) | <0.001 | 0.1 (0.09 -0.1) |
| 3-6mo | Major adverse cardiovascular events | 1507 (0.2%) | 3862 (0.3%) | <0.001 | 0.62 (0.58 -0.66) |

| | | | | | |
|---|---|---|---|---|---|
| | Cardiovascular disease | 2046 (0.3%) | 5506 (0.5%) | <0.001 | 0.57 (0.54 -0.6) |
| | Ischemic heart disease | 902 (0.1%) | 2347 (0.2%) | <0.001 | 0.62 (0.57 -0.67) |
| | Myocardial angina | 153 (0%) | 346 (0%) | 0.001 | 0.73 (0.6 -0.88) |
| | Acute coronary disease | 121 (0%) | 366 (0%) | <0.001 | 0.55 (0.44 -0.67) |
| | Myocardial infarction | 276 (0%) | 850 (0.1%) | <0.001 | 0.53 (0.47 -0.61) |
| | Ischemic cardiomyopathy | 57 (0%) | 173 (0%) | <0.001 | 0.55 (0.4 -0.74) |
| | Arrythmias | 2861 (0.4%) | 8580 (0.8%) | <0.001 | 0.5 (0.47 -0.52) |
| | Atrial fibrillation and flutter | 497 (0.1%) | 1321 (0.1%) | <0.001 | 0.61 (0.55 -0.68) |
| | Paroxysmal tachycardia | 415 (0.1%) | 1064 (0.1%) | <0.001 | 0.64 (0.57 -0.72) |
| | Other cardiac arrythmias | 952 (0.1%) | 2678 (0.2%) | <0.001 | 0.58 (0.53 -0.62) |
| | Inflammatory heart disease | 205 (0%) | 693 (0.1%) | <0.001 | 0.49 (0.42 -0.57) |
| | Pericarditis | 150 (0%) | 489 (0%) | <0.001 | 0.51 (0.42 -0.61) |
| | Myocarditis | 10 (0%) | 45 (0%) | 0.003 | 0.37 (0.19 -0.73) |
| | Endocarditis | 60 (0%) | 216 (0%) | <0.001 | 0.46 (0.35 -0.61) |
| | Hypertensive heart disease | 2695 (0.4%) | 6769 (0.7%) | <0.001 | 0.6 (0.58 -0.63) |
| | Pulmonary heart diseases | 439 (0.1%) | 1281 (0.1%) | <0.001 | 0.56 (0.5 -0.62) |
| | Heart Failure | 587 (0.1%) | 1700 (0.1%) | <0.001 | 0.56 (0.51 -0.62) |
| | Cardiomyopathy | 243 (0%) | 604 (0%) | <0.001 | 0.66 (0.57 -0.77) |
| | Cardiomegaly | 476 (0.1%) | 1415 (0.1%) | <0.001 | 0.55 (0.5 -0.61) |
| | Cardiogenic shock | 45 (0%) | 151 (0%) | <0.001 | 0.49 (0.35 -0.69) |
| | Cardiac arrest | 104 (0%) | 426 (0%) | <0.001 | 0.4 (0.33 -0.5) |
| | Thrombotic disorders | 411 (0.1%) | 1419 (0.1%) | <0.001 | 0.47 (0.42 -0.53) |
| | Pulmonary embolism | 194 (0%) | 569 (0%) | <0.001 | 0.56 (0.48 -0.66) |
| | Deep and superficial thrombosis | 309 (0%) | 1144 (0.1%) | <0.001 | 0.44 (0.39 -0.5) |
| | Thrombosis with Thrombocytopenia | 0 (0%) | 0 (0%) | NA | NA |
| | Elevated Troponin I | 261 (0%) | 1212 (0.1%) | <0.001 | 0.35 (0.31 -0.4) |
| | Elevated Natriuretic peptide B | 744 (0.1%) | 1869 (0.1%) | <0.001 | 0.64 (0.59 -0.7) |
| | Dyslipidemia | 4780 (0.8%) | 12128 (1.5%) | <0.001 | 0.57 (0.55 -0.59) |
| | Triglyceride ≥150 mg/dl | 2059 (0.3%) | 5276 (0.5%) | <0.001 | 0.62 (0.59 -0.66) |
| | Total cholesterol ≥200 mg/dl | 2935 (0.4%) | 7592 (0.7%) | <0.001 | 0.61 (0.58 -0.64) |
| | LDL-cholesterol ≥130 mg/dl | 2278 (0.3%) | 5889 (0.5%) | <0.001 | 0.62 (0.59 -0.65) |
| | HDL-cholesterol <50 mg/dl | 2665 (0.4%) | 7380 (0.7%) | <0.001 | 0.56 (0.53 -0.58) |
| | Cerebrovascular disease | 832 (0.1%) | 2165 (0.2%) | <0.001 | 0.62 (0.58 -0.68) |
| | Transient ischemic attacks | 209 (0%) | 552 (0%) | <0.001 | 0.62 (0.53 -0.73) |
| | Cerebral infarction | 321 (0%) | 884 (0.1%) | <0.001 | 0.6 (0.52 -0.68) |
| | Cerebral occlusion without infarction | 268 (0%) | 737 (0.1%) | <0.001 | 0.6 (0.52 -0.69) |
| | Intracerebral hemorrhage | 62 (0%) | 202 (0%) | <0.001 | 0.51 (0.38 -0.68) |
| | Subarachnoid hemorrhage | 41 (0%) | 114 (0%) | 0.004 | 0.6 (0.42 -0.85) |
| | Hospital admission | 3376 (0.5%) | 11382 (1.3%) | <0.001 | 0.39 (0.38 -0.41) |
| | Emergency visits | 5940 (1.1%) | 11419 (2.1%) | <0.001 | 0.52 (0.5 -0.53) |
| | Outpatient encounters | 6618 (1.5%) | 12376 (2.1%) | <0.001 | 0.72 (0.7 -0.74) |
| | Intensive care unit admission | 124 (0%) | 790 (0.1%) | <0.001 | 0.25 (0.21 -0.31) |
| | Mechanical ventilation | 252 (0%) | 1090 (0.1%) | <0.001 | 0.38 (0.33 -0.43) |
| | Referral to Cardio services | 0 (0%) | 10 (0%) | 0.014 | NA |
| | Referral to Neuro services | 10 (0%) | 22 (0%) | 0.455 | 0.75 (0.36 -1.59) |
| | ECMO | 10 (0%) | 10 (0%) | 0.255 | 1.66 (0.69 -3.98) |
| | ECMO-VV | 10 (0%) | 10 (0%) | 0.254 | 1.66 (0.69 -3.98) |
| | ECMO-VA | 10 (0%) | 10 (0%) | 0.254 | 1.66 (0.69 -3.98) |
| | Mortality | 1376 (0.2%) | 4970 (0.4%) | <0.001 | 0.46 (0.43 -0.49) |
| 6-9mo | Major adverse cardiovascular events | 1424 (0.2%) | 3160 (0.3%) | <0.001 | 0.71 (0.67 -0.76) |
| | Cardiovascular disease | 1897 (0.3%) | 4511 (0.4%) | <0.001 | 0.65 (0.61 -0.68) |
| | Ischemic heart disease | 823 (0.1%) | 1883 (0.2%) | <0.001 | 0.7 (0.65 -0.76) |
| | Myocardial angina | 118 (0%) | 321 (0%) | <0.001 | 0.61 (0.49 -0.75) |
| | Acute coronary disease | 97 (0%) | 227 (0%) | 0.004 | 0.71 (0.56 -0.89) |
| | Myocardial infarction | 260 (0%) | 671 (0.1%) | <0.001 | 0.64 (0.55 -0.73) |
| | Ischemic cardiomyopathy | 56 (0%) | 130 (0%) | 0.033 | 0.71 (0.52 -0.97) |

| | | | | | |
|---|---|---|---|---|---|
| | Arrythmias | 2665 (0.4%) | 6860 (0.6%) | <0.001 | 0.57 (0.55 -0.6) |
| | Atrial fibrillation and flutter | 467 (0.1%) | 943 (0.1%) | <0.001 | 0.81 (0.72 -0.9) |
| | Paroxysmal tachycardia | 343 (0%) | 820 (0.1%) | <0.001 | 0.69 (0.61 -0.78) |
| | Other cardiac arrythmias | 852 (0.1%) | 2128 (0.2%) | <0.001 | 0.65 (0.6 -0.7) |
| | Inflammatory heart disease | 183 (0%) | 495 (0%) | <0.001 | 0.61 (0.51 -0.72) |
| | Pericarditis | 139 (0%) | 349 (0%) | <0.001 | 0.66 (0.54 -0.8) |
| | Myocarditis | 10 (0%) | 26 (0%) | 0.221 | 0.64 (0.31 -1.32) |
| | Endocarditis | 50 (0%) | 149 (0%) | <0.001 | 0.56 (0.4 -0.77) |
| | Hypertensive heart disease | 2507 (0.4%) | 5599 (0.6%) | <0.001 | 0.67 (0.64 -0.71) |
| | Pulmonary heart diseases | 374 (0%) | 982 (0.1%) | <0.001 | 0.62 (0.55 -0.7) |
| | Heart Failure | 564 (0.1%) | 1215 (0.1%) | <0.001 | 0.75 (0.68 -0.83) |
| | Cardiomyopathy | 213 (0%) | 476 (0%) | <0.001 | 0.74 (0.63 -0.87) |
| | Cardiomegaly | 431 (0.1%) | 1196 (0.1%) | <0.001 | 0.59 (0.53 -0.66) |
| | Cardiogenic shock | 37 (0%) | 124 (0%) | <0.001 | 0.49 (0.34 -0.71) |
| | Cardiac arrest | 99 (0%) | 298 (0%) | <0.001 | 0.55 (0.44 -0.69) |
| | Thrombotic disorders | 361 (0%) | 1139 (0.1%) | <0.001 | 0.52 (0.46 -0.58) |
| | Pulmonary embolism | 165 (0%) | 460 (0%) | <0.001 | 0.59 (0.49 -0.7) |
| | Deep and superficial thrombosis | 260 (0%) | 873 (0.1%) | <0.001 | 0.49 (0.42 -0.56) |
| | Thrombosis with Thrombocytopenia | 0 (0%) | 10 (0%) | 0.014 | NA |
| | Elevated Troponin I | 236 (0%) | 1101 (0.1%) | <0.001 | 0.35 (0.3 -0.4) |
| | Elevated Natriuretic peptide B | 702 (0.1%) | 1469 (0.1%) | <0.001 | 0.77 (0.7 -0.84) |
| | Dyslipidemia | 4264 (0.8%) | 10235 (1.3%) | <0.001 | 0.59 (0.57 -0.62) |
| | Triglyceride ≥150 mg/dl | 1980 (0.3%) | 4642 (0.4%) | <0.001 | 0.68 (0.65 -0.72) |
| | Total cholesterol ≥200 mg/dl | 2771 (0.4%) | 6473 (0.6%) | <0.001 | 0.67 (0.64 -0.7) |
| | LDL-cholesterol ≥130 mg/dl | 2191 (0.3%) | 5219 (0.5%) | <0.001 | 0.67 (0.64 -0.7) |
| | HDL-cholesterol <50 mg/dl | 2436 (0.4%) | 6576 (0.6%) | <0.001 | 0.57 (0.54 -0.6) |
| | Cerebrovascular disease | 777 (0.1%) | 1831 (0.1%) | <0.001 | 0.69 (0.63 -0.75) |
| | Transient ischemic attacks | 208 (0%) | 488 (0%) | <0.001 | 0.7 (0.6 -0.83) |
| | Cerebral infarction | 310 (0%) | 727 (0.1%) | <0.001 | 0.7 (0.61 -0.8) |
| | Cerebral occlusion without infarction | 261 (0%) | 591 (0%) | <0.001 | 0.73 (0.63 -0.84) |
| | Intracerebral hemorrhage | 65 (0%) | 139 (0%) | 0.086 | 0.77 (0.58 -1.04) |
| | Subarachnoid hemorrhage | 44 (0%) | 105 (0%) | 0.04 | 0.69 (0.49 -0.99) |
| | Hospital admission | 2771 (0.4%) | 8724 (1%) | <0.001 | 0.42 (0.4 -0.43) |
| | Emergency visits | 5343 (1%) | 10084 (1.9%) | <0.001 | 0.52 (0.5 -0.54) |
| | Outpatient encounters | 5726 (1.3%) | 9357 (1.6%) | <0.001 | 0.82 (0.8 -0.85) |
| | Intensive care unit admission | 79 (0%) | 674 (0.1%) | <0.001 | 0.19 (0.15 -0.24) |
| | Mechanical ventilation | 252 (0%) | 809 (0.1%) | <0.001 | 0.51 (0.44 -0.59) |
| | Referral to Cardio services | 0 (0%) | 0 (0%) | NA | NA |
| | Referral to Neuro services | 10 (0%) | 14 (0%) | 0.685 | 1.18 (0.53 -2.66) |
| | ECMO | 0 (0%) | 10 (0%) | 0.014 | NA |
| | ECMO-VV | 0 (0%) | 10 (0%) | 0.014 | NA |
| | ECMO-VA | 0 (0%) | 10 (0%) | 0.014 | NA |
| | Mortality | 1442 (0.2%) | 3680 (0.3%) | <0.001 | 0.65 (0.61 -0.69) |
| >9mo | Major adverse cardiovascular events | 1424 (0.2%) | 3160 (0.3%) | <0.001 | 0.71 (0.67 -0.76) |
| | Cardiovascular disease | 1897 (0.3%) | 4511 (0.4%) | <0.001 | 0.65 (0.61 -0.68) |
| | Ischemic heart disease | 823 (0.1%) | 1883 (0.2%) | <0.001 | 0.7 (0.65 -0.76) |
| | Myocardial angina | 118 (0%) | 321 (0%) | <0.001 | 0.61 (0.49 -0.75) |
| | Acute coronary disease | 97 (0%) | 227 (0%) | 0.004 | 0.71 (0.56 -0.89) |
| | Myocardial infarction | 260 (0%) | 671 (0.1%) | <0.001 | 0.64 (0.55 -0.73) |
| | Ischemic cardiomyopathy | 56 (0%) | 130 (0%) | 0.033 | 0.71 (0.52 -0.97) |
| | Arrythmias | 2665 (0.4%) | 6860 (0.6%) | <0.001 | 0.57 (0.55 -0.6) |
| | Atrial fibrillation and flutter | 467 (0.1%) | 943 (0.1%) | <0.001 | 0.81 (0.72 -0.9) |
| | Paroxysmal tachycardia | 343 (0%) | 820 (0.1%) | <0.001 | 0.69 (0.61 -0.78) |
| | Other cardiac arrythmias | 852 (0.1%) | 2128 (0.2%) | <0.001 | 0.65 (0.6 -0.7) |
| | Inflammatory heart disease | 183 (0%) | 495 (0%) | <0.001 | 0.61 (0.51 -0.72) |
| | Pericarditis | 139 (0%) | 349 (0%) | <0.001 | 0.66 (0.54 -0.8) |

| | | | | | |
|---|---|---|---|---|---|
| | Myocarditis | 10 (0%) | 26 (0%) | 0.221 | 0.64 (0.31 -1.32) |
| | Endocarditis | 50 (0%) | 149 (0%) | <0.001 | 0.56 (0.4 -0.77) |
| | Hypertensive heart disease | 2507 (0.4%) | 5599 (0.6%) | <0.001 | 0.67 (0.64 -0.71) |
| | Pulmonary heart diseases | 374 (0%) | 982 (0.1%) | <0.001 | 0.62 (0.55 -0.7) |
| | Heart Failure | 564 (0.1%) | 1215 (0.1%) | <0.001 | 0.75 (0.68 -0.83) |
| | Cardiomyopathy | 213 (0%) | 476 (0%) | <0.001 | 0.74 (0.63 -0.87) |
| | Cardiomegaly | 431 (0.1%) | 1196 (0.1%) | <0.001 | 0.59 (0.53 -0.66) |
| | Cardiogenic shock | 37 (0%) | 124 (0%) | <0.001 | 0.49 (0.34 -0.71) |
| | Cardiac arrest | 99 (0%) | 298 (0%) | <0.001 | 0.55 (0.44 -0.69) |
| | Thrombotic disorders | 361 (0%) | 1139 (0.1%) | <0.001 | 0.52 (0.46 -0.58) |
| | Pulmonary embolism | 165 (0%) | 460 (0%) | <0.001 | 0.59 (0.49 -0.7) |
| | Deep and superficial thrombosis | 260 (0%) | 873 (0.1%) | <0.001 | 0.49 (0.42 -0.56) |
| | Thrombosis with Thrombocytopenia | 0 (0%) | 10 (0%) | 0.014 | NA |
| | Elevated Troponin I | 236 (0%) | 1101 (0.1%) | <0.001 | 0.35 (0.3 -0.4) |
| | Elevated Natriuretic peptide B | 702 (0.1%) | 1469 (0.1%) | <0.001 | 0.77 (0.7 -0.84) |
| | Dyslipidemia | 4264 (0.8%) | 10235 (1.3%) | <0.001 | 0.59 (0.57 -0.62) |
| | Triglyceride ≥150 mg/dl | 1980 (0.3%) | 4642 (0.4%) | <0.001 | 0.68 (0.65 -0.72) |
| | Total cholesterol ≥200 mg/dl | 2771 (0.4%) | 6473 (0.6%) | <0.001 | 0.67 (0.64 -0.7) |
| | LDL-cholesterol ≥130 mg/dl | 2191 (0.3%) | 5219 (0.5%) | <0.001 | 0.67 (0.64 -0.7) |
| | HDL-cholesterol <50 mg/dl | 2436 (0.4%) | 6576 (0.6%) | <0.001 | 0.57 (0.54 -0.6) |
| | Cerebrovascular disease | 777 (0.1%) | 1831 (0.1%) | <0.001 | 0.69 (0.63 -0.75) |
| | Transient ischemic attacks | 208 (0%) | 488 (0%) | <0.001 | 0.7 (0.6 -0.83) |
| | Cerebral infarction | 310 (0%) | 727 (0.1%) | <0.001 | 0.7 (0.61 -0.8) |
| | Cerebral occlusion without infarction | 261 (0%) | 591 (0%) | <0.001 | 0.73 (0.63 -0.84) |
| | Intracerebral hemorrhage | 65 (0%) | 139 (0%) | 0.086 | 0.77 (0.58 -1.04) |
| | Subarachnoid hemorrhage | 44 (0%) | 105 (0%) | 0.04 | 0.69 (0.49 -0.99) |
| | Hospital admission | 2771 (0.4%) | 8724 (1%) | <0.001 | 0.42 (0.4 -0.43) |
| | Emergency visits | 5343 (1%) | 10084 (1.9%) | <0.001 | 0.52 (0.5 -0.54) |
| | Outpatient encounters | 5726 (1.3%) | 9357 (1.6%) | <0.001 | 0.82 (0.8 -0.85) |
| | Intensive care unit admission | 79 (0%) | 674 (0.1%) | <0.001 | 0.19 (0.15 -0.24) |
| | Mechanical ventilation | 252 (0%) | 809 (0.1%) | <0.001 | 0.51 (0.44 -0.59) |
| | Referral to Cardio services | 0 (0%) | 0 (0%) | NA | NA |
| | Referral to Neuro services | 10 (0%) | 14 (0%) | 0.685 | 1.18 (0.53 -2.66) |
| | ECMO | 0 (0%) | 10 (0%) | 0.014 | NA |
| | ECMO-VV | 0 (0%) | 10 (0%) | 0.014 | NA |
| | ECMO-VA | 0 (0%) | 10 (0%) | 0.014 | NA |
| | Mortality | 1442 (0.2%) | 3680 (0.3%) | <0.001 | 0.65 (0.61 -0.69) |

For outcomes where event counts were fewer than 10, TriNetX platform suppressed exact numbers and reported '10' for de-identification purposes; thus, relative risks may be overestimated or not statistically meaningful in these cases. When zero events were observed, this is indicated as '0'. Caution is warranted in the interpretation of RRs where numerators are at or below this privacy threshold. ECMO: Extracorporeal membrane oxygenation.

**Table S11.** Dose-dependent protection against cardiovascular events during the acute phase following Pfizer-BioNTech vaccination.

| Outcome | Group A | Roup B | *p*-value | RR (95%CI) |
|---|---|---|---|---|
| **Comparison groups** | **Monovalent Second dose (n=785,847)** | **Monovalent One dose (n=301,327)** | | |
| Major adverse cardiovascular events | 397 (0.05%) | 236 (0.08%) | <0.001 | 0.65 (0.55-0.76) |
| Cardiovascular disease | 520 (0.07%) | 301 (0.1%) | <0.001 | 0.66 (0.57-0.76) |
| Ischemic heart disease | 264 (0.03%) | 152 (0.05%) | <0.001 | 0.67 (0.55-0.81) |
| Myocardial angina | 58 (0.01%) | 16 (0.01%) | 0.244 | 1.39 (0.8-2.41) |
| Acute coronary disease | 34 (0%) | 28 (0.01%) | 0.002 | 0.47 (0.28-0.77) |
| Myocardial infarction | 69 (0.01%) | 56 (0.02%) | <0.001 | 0.47 (0.33-0.67) |
| Ischemic cardiomyopathy | 29 (0%) | 20 (0.01%) | 0.04 | 0.56 (0.31-0.98) |
| Arrythmias | 670 (0.09%) | 410 (0.14%) | <0.001 | 0.62 (0.54-0.7) |
| Atrial fibrillation and flutter | 142 (0.02%) | 84 (0.03%) | 0.002 | 0.65 (0.5-0.85) |
| Paroxysmal tachycardia | 88 (0.01%) | 75 (0.02%) | <0.001 | 0.45 (0.33-0.61) |
| Other cardiac arrythmias | 218 (0.03%) | 157 (0.05%) | <0.001 | 0.53 (0.43-0.65) |
| Inflammatory heart disease | 56 (0.01%) | 44 (0.01%) | <0.001 | 0.49 (0.33-0.72) |
| Pericarditis | 37 (0%) | 34 (0.01%) | <0.001 | 0.42 (0.26-0.66) |
| Myocarditis | 10 (0%) | 10 (0%) | 0.026 | 0.38 (0.16-0.92) |
| Endocarditis | 17 (0%) | 14 (0%) | 0.03 | 0.47 (0.23-0.94) |
| Hypertensive heart disease | 750 (0.11%) | 391 (0.15%) | <0.001 | 0.73 (0.64-0.82) |
| Pulmonary heart diseases | 89 (0.01%) | 74 (0.02%) | <0.001 | 0.46 (0.34-0.63) |
| Heart Failure | 165 (0.02%) | 95 (0.03%) | 0.001 | 0.66 (0.52-0.86) |
| Cardiomyopathy | 77 (0.01%) | 58 (0.02%) | <0.001 | 0.51 (0.36-0.72) |
| Cardiomegaly | 99 (0.01%) | 87 (0.03%) | <0.001 | 0.43 (0.33-0.58) |
| Cardiogenic shock | 14 (0%) | 21 (0.01%) | <0.001 | 0.26 (0.13-0.5) |
| Cardiac arrest | 31 (0%) | 38 (0.01%) | <0.001 | 0.31 (0.19-0.5) |
| Thrombotic disorders | 85 (0.01%) | 60 (0.02%) | <0.001 | 0.54 (0.39-0.75) |
| Pulmonary embolism | 30 (0%) | 28 (0.01%) | <0.001 | 0.41 (0.25-0.69) |
| Deep and superficial thrombosis | 67 (0.01%) | 41 (0.01%) | 0.017 | 0.63 (0.42-0.92) |
| Thrombosis with Thrombocytopenia | 51 (0.01%) | 55 (0.02%) | <0.001 | 0.36 (0.24-0.52) |
| Elevated Troponin I | 200 (0.02%) | 137 (0.04%) | <0.001 | 0.56 (0.45-0.69) |
| Elevated Natriuretic peptide B | 1186 (0.19%) | 1225 (0.53%) | <0.001 | 0.35 (0.32-0.38) |
| Dyslipidemia | 469 (0.06%) | 533 (0.19%) | <0.001 | 0.33 (0.29-0.37) |
| Triglyceride ≥150 mg/dl | 543 (0.07%) | 627 (0.23%) | <0.001 | 0.32 (0.29-0.36) |
| Total cholesterol ≥200 mg/dl | 430 (0.06%) | 520 (0.18%) | <0.001 | 0.31 (0.27-0.35) |
| LDL-cholesterol ≥130 mg/dl | 780 (0.11%) | 978 (0.37%) | <0.001 | 0.29 (0.27-0.32) |
| HDL-cholesterol <50 mg/dl | 190 (0.02%) | 118 (0.04%) | <0.001 | 0.62 (0.49-0.78) |
| Cerebrovascular disease | 51 (0.01%) | 27 (0.01%) | 0.173 | 0.72 (0.45-1.15) |
| Transient ischemic attacks | 79 (0.01%) | 53 (0.02%) | 0.001 | 0.57 (0.4-0.81) |
| Cerebral infarction | 64 (0.01%) | 45 (0.01%) | 0.002 | 0.55 (0.37-0.8) |
| Cerebral occlusion without infarction | 14 (0%) | 11 (0%) | 0.068 | 0.49 (0.22-1.07) |
| Intracerebral hemorrhage | 546 (0.08%) | 653 (0.26%) | <0.001 | 0.3 (0.27-0.34) |
| Subarachnoid hemorrhage | 1241 (0.21%) | 806 (0.36%) | <0.001 | 0.57 (0.52-0.63) |
| Hospital admission | 2151 (0.45%) | 1465 (1%) | <0.001 | 0.45 (0.42-0.48) |
| Emergency visits | 18 (0%) | 35 (0.01%) | <0.001 | 0.2 (0.11-0.35) |
| Outpatient encounters | 67 (0.01%) | 75 (0.02%) | <0.001 | 0.34 (0.25-0.48) |
| Intensive care unit admission | 187 (0.02%) | 306 (0.1%) | <0.001 | 0.23 (0.2-0.28) |
| Mechanical ventilation | 397 (0.05%) | 236 (0.08%) | <0.001 | 0.65 (0.55-0.76) |
| Referral to Cardio services | 520 (0.07%) | 301 (0.1%) | <0.001 | 0.66 (0.57-0.76) |
| Referral to Neuro services | 264 (0.03%) | 152 (0.05%) | <0.001 | 0.67 (0.55-0.81) |
| ECMO | 58 (0.01%) | 16 (0.01%) | 0.244 | 1.39 (0.8-2.41) |

| | | | | |
|---|---|---|---|---|
| ECMO-VV | 34 (0%) | 28 (0.01%) | 0.002 | 0.47 (0.28-0.77) |
| ECMO-VA | 69 (0.01%) | 56 (0.02%) | <0.001 | 0.47 (0.33-0.67) |
| Mortality | 29 (0%) | 20 (0.01%) | 0.04 | 0.56 (0.31-0.98) |
| **Comparison groups** | **Monovalent Booster dose (n=252,933)** | **Monovalent One dose (n=306,413)** | | |
| Major adverse cardiovascular events | 215 (0.09%) | 248 (0.08%) | 0.599 | 1.05 (0.88-1.26) |
| Cardiovascular disease | 278 (0.11%) | 316 (0.11%) | 0.366 | 1.08 (0.92-1.27) |
| Ischemic heart disease | 137 (0.05%) | 159 (0.05%) | 0.774 | 1.03 (0.82-1.3) |
| Myocardial angina | 13 (0%) | 18 (0.01%) | 0.668 | 0.86 (0.42-1.75) |
| Acute coronary disease | 22 (0.01%) | 30 (0.01%) | 0.611 | 0.87 (0.5-1.5) |
| Myocardial infarction | 40 (0.01%) | 58 (0.02%) | 0.324 | 0.82 (0.55-1.22) |
| Ischemic cardiomyopathy | 13 (0%) | 21 (0.01%) | 0.376 | 0.73 (0.37-1.46) |
| Arrythmias | 327 (0.14%) | 431 (0.15%) | 0.32 | 0.93 (0.81-1.07) |
| Atrial fibrillation and flutter | 80 (0.03%) | 89 (0.03%) | 0.661 | 1.07 (0.79-1.45) |
| Paroxysmal tachycardia | 71 (0.03%) | 80 (0.02%) | 0.751 | 1.05 (0.77-1.45) |
| Other cardiac arrythmias | 129 (0.05%) | 165 (0.05%) | 0.576 | 0.94 (0.74-1.18) |
| Inflammatory heart disease | 25 (0.01%) | 47 (0.01%) | 0.059 | 0.63 (0.39-1.02) |
| Pericarditis | 22 (0.01%) | 36 (0.01%) | 0.228 | 0.72 (0.43-1.23) |
| Myocarditis | 0 (0%) | 10 (0%) | 0.004 | NA |
| Endocarditis | 10 (0%) | 15 (0%) | 0.559 | 0.79 (0.35-1.76) |
| Hypertensive heart disease | 335 (0.16%) | 407 (0.15%) | 0.41 | 1.06 (0.92-1.23) |
| Pulmonary heart diseases | 68 (0.03%) | 79 (0.02%) | 0.896 | 1.02 (0.74-1.41) |
| Heart Failure | 90 (0.03%) | 101 (0.03%) | 0.693 | 1.06 (0.8-1.41) |
| Cardiomyopathy | 38 (0.01%) | 63 (0.02%) | 0.102 | 0.72 (0.48-1.07) |
| Cardiomegaly | 62 (0.02%) | 97 (0.03%) | 0.087 | 0.76 (0.55-1.04) |
| Cardiogenic shock | 12 (0%) | 24 (0.01%) | 0.132 | 0.59 (0.3-1.18) |
| Cardiac arrest | 24 (0.01%) | 41 (0.01%) | 0.149 | 0.69 (0.42-1.15) |
| Thrombotic disorders | 48 (0.02%) | 64 (0.02%) | 0.547 | 0.89 (0.61-1.3) |
| Pulmonary embolism | 24 (0.01%) | 31 (0.01%) | 0.749 | 0.92 (0.54-1.56) |
| Deep and superficial thrombosis | 33 (0.01%) | 43 (0.01%) | 0.687 | 0.91 (0.58-1.43) |
| Thrombosis with Thrombocytopenia | 34 (0.01%) | 56 (0.02%) | 0.127 | 0.72 (0.47-1.1) |
| Elevated Troponin I | 96 (0.04%) | 148 (0.05%) | 0.046 | 0.77 (0.6-1) |
| Elevated Natriuretic peptide B | 1012 (0.61%) | 1255 (0.54%) | 0.001 | 1.15 (1.06-1.24) |
| Dyslipidemia | 430 (0.19%) | 541 (0.19%) | 0.948 | 1 (0.88-1.13) |
| Triglyceride ≥150 mg/dl | 558 (0.25%) | 638 (0.23%) | 0.059 | 1.12 (1-1.25) |
| Total cholesterol ≥200 mg/dl | 471 (0.2%) | 534 (0.18%) | 0.138 | 1.1 (0.97-1.24) |
| LDL-cholesterol ≥130 mg/dl | 783 (0.37%) | 1004 (0.37%) | 0.92 | 1.01 (0.92-1.1) |
| HDL-cholesterol <50 mg/dl | 116 (0.04%) | 127 (0.04%) | 0.489 | 1.09 (0.85-1.41) |
| Cerebrovascular disease | 32 (0.01%) | 28 (0.01%) | 0.238 | 1.36 (0.82-2.25) |
| Transient ischemic attacks | 51 (0.02%) | 56 (0.02%) | 0.69 | 1.08 (0.74-1.58) |
| Cerebral infarction | 45 (0.02%) | 47 (0.01%) | 0.539 | 1.14 (0.76-1.71) |
| Cerebral occlusion without infarction | 16 (0.01%) | 14 (0%) | 0.409 | 1.35 (0.66-2.77) |
| Intracerebral hemorrhage | 337 (0.15%) | 683 (0.26%) | <0.001 | 0.58 (0.51-0.66) |
| Subarachnoid hemorrhage | 615 (0.35%) | 842 (0.37%) | 0.232 | 0.94 (0.85-1.04) |
| Hospital admission | 872 (0.66%) | 1482 (0.98%) | <0.001 | 0.68 (0.62-0.74) |
| Emergency visits | 28 (0.01%) | 35 (0.01%) | 0.826 | 0.95 (0.58-1.56) |
| Outpatient encounters | 52 (0.02%) | 80 (0.02%) | 0.138 | 0.77 (0.54-1.09) |
| Intensive care unit admission | 208 (0.08%) | 335 (0.1%) | <0.001 | 0.73 (0.62-0.87) |
| Mechanical ventilation | 215 (0.09%) | 248 (0.08%) | 0.599 | 1.05 (0.88-1.26) |
| Referral to Cardio services | 278 (0.11%) | 316 (0.11%) | 0.366 | 1.08 (0.92-1.27) |
| Referral to Neuro services | 137 (0.05%) | 159 (0.05%) | 0.774 | 1.03 (0.82-1.3) |
| ECMO | 13 (0%) | 18 (0.01%) | 0.668 | 0.86 (0.42-1.75) |

| | | | | |
|---|---|---|---|---|
| ECMO-VV | 22 (0.01%) | 30 (0.01%) | 0.611 | 0.87 (0.5-1.5) |
| ECMO-VA | 40 (0.01%) | 58 (0.02%) | 0.324 | 0.82 (0.55-1.22) |
| Mortality | 13 (0%) | 21 (0.01%) | 0.376 | 0.73 (0.37-1.46) |
| **Comparison groups** | **Bivalent vaccines Booster dose (n=62,794)** | **Bivalent vaccines One dose (n=47,377)** | | |
| Major adverse cardiovascular events | 70 (0.11%) | 36 (0.07%) | 0.063 | 1.46 (0.98-2.18) |
| Cardiovascular disease | 125 (0.2%) | 71 (0.15%) | 0.055 | 1.33 (0.99-1.78) |
| Ischemic heart disease | 50 (0.07%) | 25 (0.05%) | 0.096 | 1.5 (0.93-2.42) |
| Myocardial angina | 10 (0.01%) | 10 (0.02%) | 0.514 | 0.75 (0.31-1.8) |
| Acute coronary disease | 10 (0.01%) | 10 (0.02%) | 0.512 | 0.75 (0.31-1.79) |
| Myocardial infarction | 16 (0.02%) | 12 (0.02%) | 0.993 | 1 (0.47-2.11) |
| Ischemic cardiomyopathy | 10 (0.01%) | 0 (0%) | 0.006 | NA |
| Arrythmias | 138 (0.23%) | 84 (0.18%) | 0.12 | 1.24 (0.95-1.63) |
| Atrial fibrillation and flutter | 29 (0.04%) | 20 (0.04%) | 0.78 | 1.08 (0.61-1.92) |
| Paroxysmal tachycardia | 17 (0.02%) | 10 (0.02%) | 0.548 | 1.27 (0.58-2.77) |
| Other cardiac arrythmias | 65 (0.1%) | 39 (0.08%) | 0.267 | 1.25 (0.84-1.86) |
| Inflammatory heart disease | 10 (0.01%) | 10 (0.02%) | 0.513 | 0.75 (0.31-1.8) |
| Pericarditis | 10 (0.01%) | 10 (0.02%) | 0.512 | 0.75 (0.31-1.79) |
| Myocarditis | 10 (0.01%) | 10 (0.02%) | 0.511 | 0.75 (0.31-1.79) |
| Endocarditis | 10 (0.01%) | 10 (0.02%) | 0.512 | 0.75 (0.31-1.79) |
| Hypertensive heart disease | 143 (0.28%) | 81 (0.21%) | 0.036 | 1.34 (1.02-1.76) |
| Pulmonary heart diseases | 23 (0.03%) | 14 (0.03%) | 0.543 | 1.23 (0.63-2.39) |
| Heart Failure | 27 (0.04%) | 18 (0.03%) | 0.707 | 1.12 (0.62-2.04) |
| Cardiomyopathy | 11 (0.02%) | 10 (0.02%) | 0.651 | 0.82 (0.35-1.93) |
| Cardiomegaly | 34 (0.05%) | 20 (0.04%) | 0.393 | 1.27 (0.73-2.21) |
| Cardiogenic shock | 10 (0.01%) | 10 (0.02%) | 0.511 | 0.75 (0.31-1.79) |
| Cardiac arrest | 10 (0.01%) | 10 (0.02%) | 0.511 | 0.75 (0.31-1.79) |
| Thrombotic disorders | 16 (0.02%) | 10 (0.02%) | 0.656 | 1.2 (0.54-2.64) |
| Pulmonary embolism | 10 (0.01%) | 10 (0.02%) | 0.513 | 0.75 (0.31-1.79) |
| Deep and superficial thrombosis | 12 (0.02%) | 10 (0.02%) | 0.799 | 0.9 (0.39-2.08) |
| Thrombosis with Thrombocytopenia | 11 (0.02%) | 10 (0.02%) | 0.653 | 0.82 (0.35-1.94) |
| Elevated Troponin I | 26 (0.04%) | 12 (0.02%) | 0.163 | 1.62 (0.82-3.21) |
| Elevated Natriuretic peptide B | 722 (2.14%) | 541 (2.07%) | 0.578 | 1.03 (0.92-1.15) |
| Dyslipidemia | 285 (0.52%) | 211 (0.51%) | 0.818 | 1.02 (0.86-1.22) |
| Triglyceride ≥150 mg/dl | 434 (0.86%) | 326 (0.86%) | 0.985 | 1 (0.87-1.15) |
| Total cholesterol ≥200 mg/dl | 374 (0.68%) | 285 (0.69%) | 0.839 | 0.98 (0.84-1.15) |
| LDL-cholesterol ≥130 mg/dl | 587 (1.26%) | 453 (1.27%) | 0.834 | 0.99 (0.87-1.12) |
| HDL-cholesterol <50 mg/dl | 31 (0.05%) | 22 (0.04%) | 0.846 | 1.06 (0.61-1.82) |
| Cerebrovascular disease | 10 (0.01%) | 10 (0.02%) | 0.513 | 0.75 (0.31-1.79) |
| Transient ischemic attacks | 10 (0.01%) | 10 (0.02%) | 0.513 | 0.75 (0.31-1.8) |
| Cerebral infarction | 10 (0.01%) | 10 (0.02%) | 0.514 | 0.75 (0.31-1.8) |
| Cerebral occlusion without infarction | 10 (0.01%) | 10 (0.02%) | 0.512 | 0.75 (0.31-1.79) |
| Intracerebral hemorrhage | 213 (0.38%) | 205 (0.49%) | 0.008 | 0.77 (0.64-0.93) |
| Subarachnoid hemorrhage | 223 (0.5%) | 157 (0.46%) | 0.419 | 1.09 (0.89-1.33) |
| Hospital admission | 247 (1.19%) | 236 (3.48%) | <0.001 | 0.34 (0.29-0.41) |
| Emergency visits | 10 (0.01%) | 10 (0.02%) | 0.515 | 0.75 (0.31-1.8) |
| Outpatient encounters | 14 (0.02%) | 10 (0.02%) | 0.915 | 1.05 (0.46-2.35) |
| Intensive care unit admission | 59 (0.08%) | 35 (0.07%) | 0.281 | 1.26 (0.83-1.91) |
| Mechanical ventilation | 70 (0.11%) | 36 (0.07%) | 0.063 | 1.46 (0.98-2.18) |
| Referral to Cardio services | 125 (0.2%) | 71 (0.15%) | 0.055 | 1.33 (0.99-1.78) |
| Referral to Neuro services | 50 (0.07%) | 25 (0.05%) | 0.096 | 1.5 (0.93-2.42) |
| ECMO | 10 (0.01%) | 10 (0.02%) | 0.514 | 0.75 (0.31-1.8) |

| | | | | |
|---|---|---|---|---|
| ECMO-VV | 10 (0.01%) | 10 (0.02%) | 0.512 | 0.75 (0.31-1.79) |
| ECMO-VA | 16 (0.02%) | 12 (0.02%) | 0.993 | 1 (0.47-2.11) |
| Mortality | 10 (0.01%) | 0 (0%) | 0.006 | NA |

For outcomes where event counts were fewer than 10, TriNetX platform suppressed exact numbers and reported '10' for de-identification purposes; thus, relative risks may be overestimated or not statistically meaningful in these cases. When zero events were observed, this is indicated as '0'. Caution is warranted in the interpretation of RRs where numerators are at or below this privacy threshold. ECMO: Extracorporeal membrane oxygenation.
